# Neurophysiological Alterations during Sensory Processing in Autism - A Meta-Analysis

**DOI:** 10.1101/2025.05.15.25327383

**Authors:** Anjuli Ghosh, Natalia Nasarre-Nacenta, Sarah Baumeister, Nathalie E. Holz, Tobias Banaschewski, Daniel Brandeis, Pascal-M. Aggensteiner, Anna Kaiser

**Affiliations:** Department of Child and Adolescent Psychiatry and Psychotherapy, Central Institute of Mental Health, Medical Faculty Mannheim, Heidelberg University, Mannheim, Germany; German Center for Mental Health (DZPG), partner site Mannheim-Heidelberg-Ulm; Department of Child and Adolescent Psychiatry and Psychotherapy, University Hospital of Psychiatry, University of Zurich, Zurich, Switzerland

## Abstract

**Objective:** While sensory processing alterations in autism are well-documented, the neurophysiological correlates remain unclear. This meta-analysis examined differences in early event-related potentials (ERP) and event-related fields (ERF) between autistic and non-autistic individuals using electroencephalography and magnetoencephalography to identify neurophysiological alterations that may underlie variations in sensory perception, communication, and social interaction in autism.

**Methods:** Following PRISMA guidelines, a database search was conducted for peer-reviewed studies published from January 1980 onwards. Random-effects meta-analyses were performed using the metafor package in R. Standardised mean-group differences in early ERP/ERF latencies and amplitudes were analysed with moderator analyses exploring demographic and methodological factors, including neurophysiological technique, sensory modality, age group, sex, and language impairment.

**Results:** 145 studies (3778 autistic, 3484 non-autistic participants) were included. Autistic individuals exhibited significantly longer latencies in P/M50 (SMD=0.44; SE=0.21; 95% CI 0.03-0.86; p=0.04), P/M100 (SMD=0.18; SE=0.08; 95% CI 0.01-0.36; p=0.03), N170 (SMD=0.33; SE=0.12; 95% CI 0.10-0.56; p=0.01), and P/M200 (SMD=0.30; SE=0.09; 95% CI 0.12-0.48; p=0.00) components. P/M50 showed the greatest latency alteration, with an effect-size nearing medium, especially in individuals with language impairment (Q_B_(2)=7.70, p=0.02), followed by N170 most notable in autistic adolescents and adults (Q_B_ (3)=12.30, p=0.01). No significant amplitude alterations were found, and substantial heterogeneity was observed.

**Conclusion:** Neurophysiological characteristics of sensory processing in autism implicate multiple mechanisms and stages given prolonged P/M50-(sensory filtering challenges) and N170-latency (social perception alterations). These component timings show potential as biomarkers, though heterogeneity and modest effect-sizes limit clinical application, highlighting the need for further research

## 1. Introduction

### 1.1. Sensory alterations in autism and their functional significance

Autism is a lifelong neurodevelopmental condition characterised by alterations in verbal and nonverbal communication, social interaction, patterns of repetitive behaviour and specialised interests (American Psychiatric Association, 2022). Affecting approximately 1% of the global population (Zeidan et al., 2022), autism is among the most prevalent neurodevelopmental conditions worldwide. Sensory alterations are highly prevalent in autism, affecting up to 90% of autistic individuals (Leekam et al., 2007; Tavassoli et al., 2014; Tomchek & Dunn, 2007). Also formally recognised as a diagnostic feature of autism in the Diagnostic and Statistical Manual of Mental Disorders, 5th edition (DSM-5) (American Psychiatric Association, 2013) and its Text Revision (DSM-5-TR) (American Psychiatric Association, 2022), as well as the International Classification of Diseases, 11^th^ edition (ICD-11) (World Health Organization, 2025), sensory alterations have been explored as potential (bio-)markers for autism (Marco et al., 2011; Shan et al., 2023). Although sensory alterations can lead to positive effects, such as improved focus, they often result in negative experiences, causing distraction, anxiety, and limiting daily activities (Jones et al., 2020). Research on sensory processing in autism focuses on auditory, tactile, and visual domains. Delayed auditory responses may contribute to language impairment in autistic children, linking slower processing to communication difficulties (Richards & Goswami, 2018). In visual processing, a focus on local details may enhance task performance but hinder social cue interpretation (Chung & Son, 2020). In tactile processing, altered neural responses to touch may affect (non-)comfort with physical contact and the emotional evaluation of touch (Cascio et al., 2015).

### 1.2. EEG and MEG: exploring brain dynamics through ERPs and ERFs

Several studies have explored neurophysiological mechanisms associated with sensory processing in autism using electroencephalography (EEG) and magnetoencephalography (MEG). These methods record early event-related potentials (ERPs) and magnetic fields (ERFs). EEG and MEG offer high temporal resolution, essential for examining sensory responses, which occur within the first few hundred milliseconds after stimulus presentation (Woodman, 2010). Both capture overlapping information (Ahlfors et al., 2022; Ahlfors & Mody, 2019), but EEG is more widely accessible and cost-effective (Berger, 1929), while MEG provides higher spatial resolution for some cortical sources and is contactless (Cohen, 1972; Fred et al., 2022), benefiting autistic individuals who often exhibit tactile sensitivities (Asmika et al., 2018). Research has identified ERP/ERF differences between autistic and non-autistic individuals, with the N170, associated with face processing, often regarded as the most promising potential neurophysiological marker in autism (Kang et al., 2018; Shan et al., 2023).

### 1.3. Objectives

Despite several important reviews (Jeste & Nelson, 2009; Marco et al., 2011), to our knowledge, no meta-analysis has yet comprehensively and systematically synthesised all early ERP/ERF components related to sensory processing in autism by quantitatively combining results from relevant studies.

Therefore, this meta-analysis summarizes EEG and MEG studies on sensory (auditory, visual, tactile, olfactory, gustatory) processing differences between autistic and non-autistic children, adolescents, and adults for a clearer understanding of the neurophysiology of sensory alterations. By examining ERP/ERF components across time frames, from early sensory registration to later processing stages, it provides a comprehensive view of timing-related sensory differences. We hypothesized smaller ERP/ERF amplitudes and longer ERP/ERF latencies in autistic individuals. Exploring heterogeneity, by potentially relevant demographic, methodological and further study-related moderators were analysed. Finally, the study aimed to identify research gaps for future exploration.

## 2. Methods

### 2.1. Search strategy

The current meta-analysis was registered on PROSPERO (https://www.crd.york.ac.uk/prospero/display_record.php?RecordID=370924).

The literature search was conducted according to the PRISMA 2020 guidelines (Page et al., 2021). An initial search was performed using the databases MEDLINE (via PubMed), PsycINFO, Web of Science, and the Clinical Trials Register on 22^nd^ and 23^rd^ of November 2022 and updated on 23^rd^ of April 2024. Additionally, studies from the reference list of five reviews (Chen et al., 2020; Feroz et al., 2022; Marco et al., 2011; Morimoto et al., 2021; Williams et al., 2021) were included. Authors were contacted to inquire about unpublished findings, clarify graphical data, or resolve uncertainties in result interpretation.

An initial pool of 1753 references was obtained. Figure 1 provides an overview of the screening process and record counts.

### 2.2. Eligibility and selection

Studies were included if they were:

(a) Recording electroencephalography (EEG) or magnetoencephalography (MEG) while participants engaged in tasks about auditory, visual, tactile, olfactory or gustatory perception.
(b) Reporting quantitative data to compare neurophysiological indices of sensory processing between autistic and non-autistic children, adolescents and adults.
(c) The autistic group having a standard clinical diagnosis of autism according to: DSM-III, DSM-III-R, DSM-IV, DSM-IV-TR, DSM-V, ICD-10 or ICD-11 or with above cut-off scores on validated rating scales ADOS or ADI-R.
(e) Published in one of the following languages: English, German, French or Spanish.
(f) Journal articles published in peer-reviewed journals.
(g) Published between January 1980 (DSM-III releasing year) and April 2024
(h) Sufficient information to calculate the effect size.

### 2.3. Data coding and data extraction

All relevant variables were collected in a coding sheet (eMethods in supplement). The first and second author collected, extracted, and coded the data from the studies independently and consulted each other during the process. Disagreements (<5%) were solved in discussion with the senior authors.

### 2.4. Quality Assessment

A combination of the modified Newcastle-Ottawa Scale (NOS) assessing the quality of nonrandomized studies in meta-analyses and a self-constructed scale evaluating the quality of EEG/MEG recordings was used (eMethods in supplement).

### 2.5. Data Synthesis

Data analysis and visualisation were conducted via the *metafor* package (Viechtbauer, 2010); (version 4.4.0) in R Studio (version 2023.09.1+494) (Posit Team, 2023). Standardised mean differences (SMD) in ERP/ERF amplitudes and latencies between autistic and non-autistic individuals were used as effect-size measure (Hedges & Olkin, 1985). For positive ERP/ERF amplitudes, negative SMD indicated smaller amplitudes in the autistic group compared to the non-autistic group. Conversely, for negative ERP/ERF amplitudes, positive SMD indicated smaller amplitudes in the autistic group. For ERP/ERF latencies, positive SMD reflected longer latencies in the autistic group compared to the non-autistic group (and vice versa). Effect-sizes were calculated using exact statistics from the publications or obtained directly from authors. A mean effect-size was then computed for each ERP/ERF component to assess alterations in amplitude and latency. Multilevel models using random-effects assumptions were fitted to estimate mean effect-sizes, allowing for inferences beyond specific studies(Borenstein et al., 2010). These models addressed data dependencies from multiple effect-sizes within the same study (e.g. multiple age groups) (Viechtbauer, 2010). Studies were weighted by heteroscedastic sampling variance, and mixed-effects models were used to explore moderator effects. This included demographic (age groups, language impairment, co-occurring condition, medication, sex, IQ), methodological (MEG/EEG, modality of stimulus presentation, task type) and further study-related (diagnostic criteria, year of publication) moderators. All moderators, except IQ and sex were categorical. Heterogeneity was assessed using Cochrane’s Q-test (Hedges & Olkin, 1985), with Q_W_ indicating residual heterogeneity and Q_B_ testing moderator impact. The REML-estimator measured heterogeneity in effect-sizes. Sensitivity analyses checked robustness, and trim-and-fill analyses explored potential publication bias by identifying and adjusting for asymmetry in the funnel plot.

## 3. Results

### 3.1. Study characteristics

7262 participants across 145 studies were included (19 studies were added during the updated search). 119 studies used EEG and 26 studies MEG. 83 studied children, 24 adolescences, and 38 adults. 20.62% of the participants were female. Characteristics of the included studies can be found in eTable 1 in supplement. A summary of demographic characteristics across all ERP/ERF components is presented in Table 1. The demographic characteristics displayed separately for each component are represented in eTable 2-15 in supplement.

**Table 1:** Demographic information across all ERP/ERF components.

|  | **Autistic** | **Non-autistic** |  |  |  |
| --- | --- | --- | --- | --- | --- |
| **Total n** | **3778** | **3484** | **t** | **df** | **p** |
| Age (years). M (SD) | 12.86 (6.92) | 13.54 (7.37) | 3.99 | 7028 | <0.0001 |
| Male (%). M (SD) | 84.25 (11.52) | 74.23 (17.17) | 27.06 | 6172 | <0.0001 |
| IQ. M (SD) | 95.70 (16.01) | 110.08 (8.55) | 30.47 | 3198 | <0.0001 |
| **Children n (<12.0y)** | **2333** | **2110** | **t** | **df** | **p** |
| Age (years). M (SD) | 8.76 (2.57) | 9.03 (2.45) | 3.58 | 4341 | 0.0003 |
| Male (%). M (SD) | 86.12 (8.56) | 75.25 (15.77) | 26.68 | 3802 | <0.0001 |
| IQ. M (SD) | 91.80 (16.66) | 108.83 (9.37) | 28.63 | 2187 | <0.0001 |
| **Adolescent n**<br>(12.0–<18.0y) | **737** | **633** | **t** | **df** | **p** |
| Age (years). M (SD) | 13.95 (1.57) | 14.33 (1.61) | 4.30 | 1298 | <0.0001 |
| Male (%). M (SD) | 83.39 (5.09) | 70.15 (16.92) | 18.72 | 1176 | <0.0001 |
| IQ. M (SD) | 96.45 (8.52) | 112.24 (4.23) | 13.64 | 223 | <0.0001 |
| **Adults n (≥18.0y)** | **708** | **741** | **t** | **df** | **p** |
| Age (years). M (SD) | 25.31 (5.02) | 25.91 (5.03) | 2.22 | 1385 | 0.03 |
| Male (%). M (SD) | 78.70 (19.29) | 74.84 (20.47) | 3.34 | 1190 | 0.0009 |
| IQ. M (SD) | 106.41<br>(10.52) | 113.15 (5.22) | 11.07 | 784 | <0.0001 |

### 3.2. Overall mean effects

The overall mean estimated SMD from the multilevel models, along with the corresponding heterogeneity estimates, are shown in Table 2.

**Table 2:**
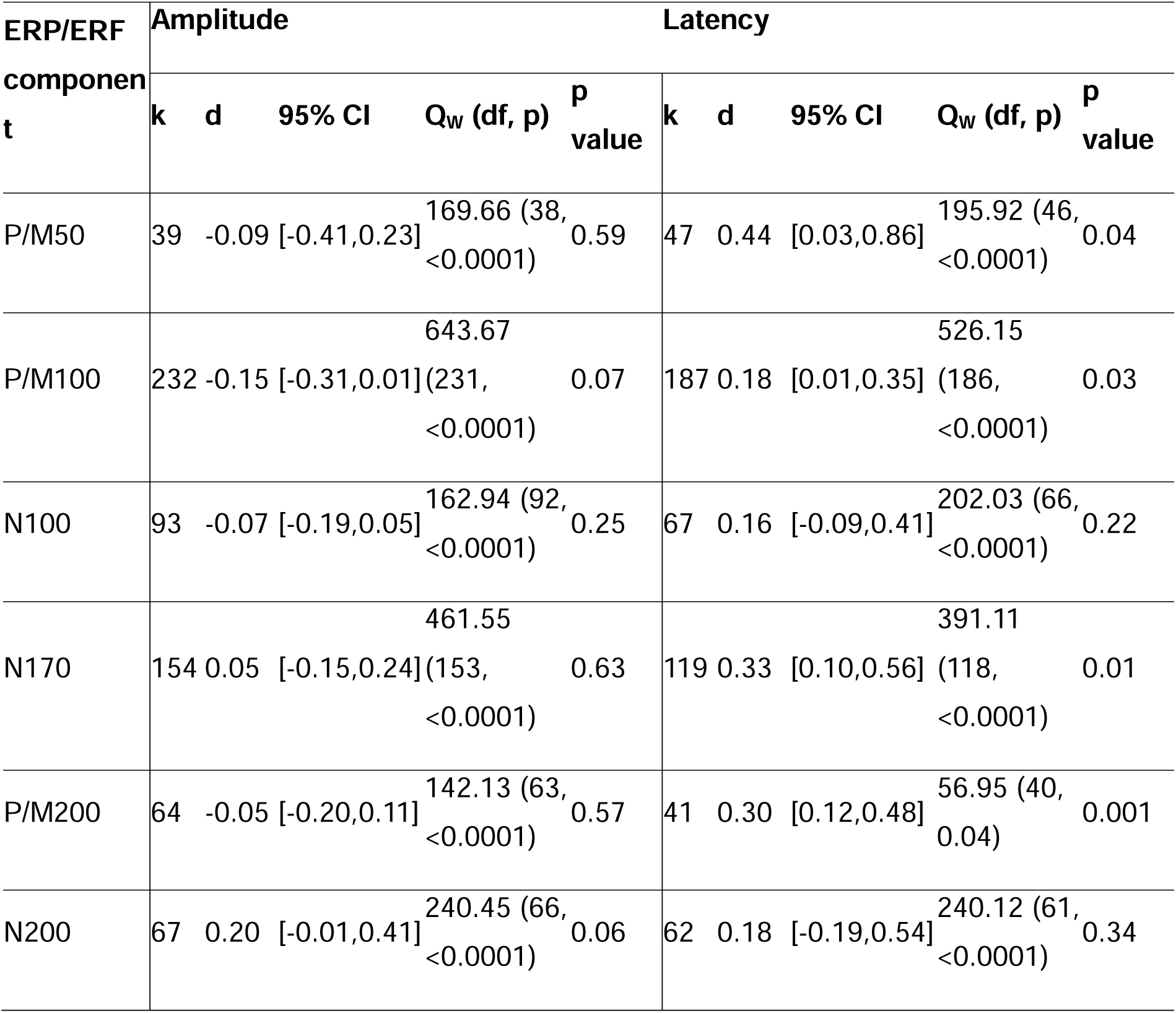
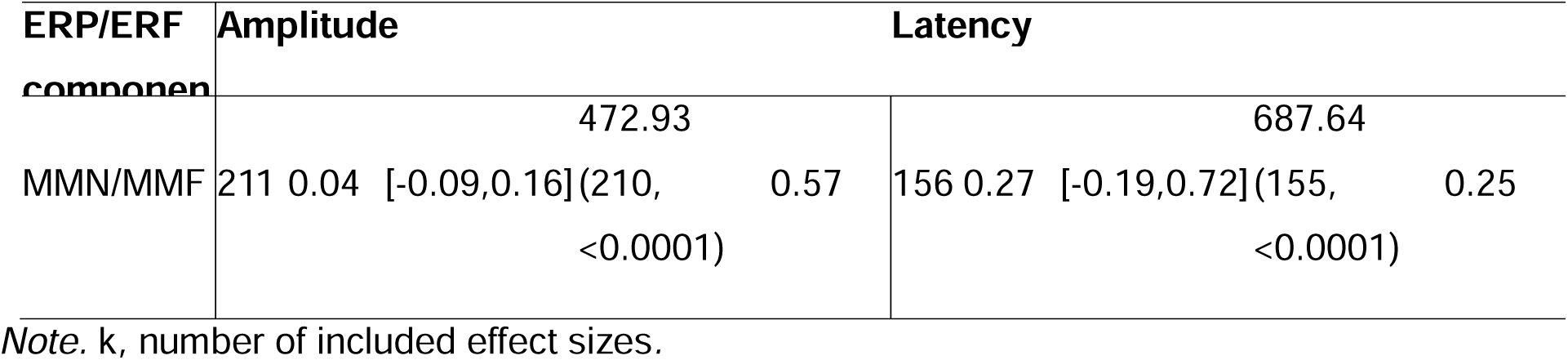
Overall mean estimated true effect-sizes for random-effects models/multilevel linear models.

#### 3.2.1. Amplitudes

No significant alterations emerged for the amplitude analyses.

#### 3.2.2. Latencies

A significant positive mean estimated SMD in the latencies of four ERP/ERF components: P/M50 (SMD=0.44 [0.03, 0.86], p=0.04), P/M100 (SMD=0.18 [0.01, 0.36], p=0.03), N170 (SMD=0.33 [0.10, 0.56], p=0.01), and P/M200 (SMD=0.30 [0.12, 0.48], p=0.001) emerged, indicating longer latencies in autistic compared to non-autistic individuals. Group differences in latencies for other ERP/ERF components remained insignificant (all p values >0.05) or lacked sufficient studies for comparison (k≤2). Forest plots for P/M50, P/M100, N170 and P/M200 latencies are shown in Figure 2-5. Forest plots for amplitudes and all other latencies are provided in the supplement eFigures 1-10.

### 3.3. Moderator effects

The Q_W_-statistics indicate substantial heterogeneity in the distribution of effect-sizes which were explored through moderator analyses (Tables 3 and 4).

**Table 3:**
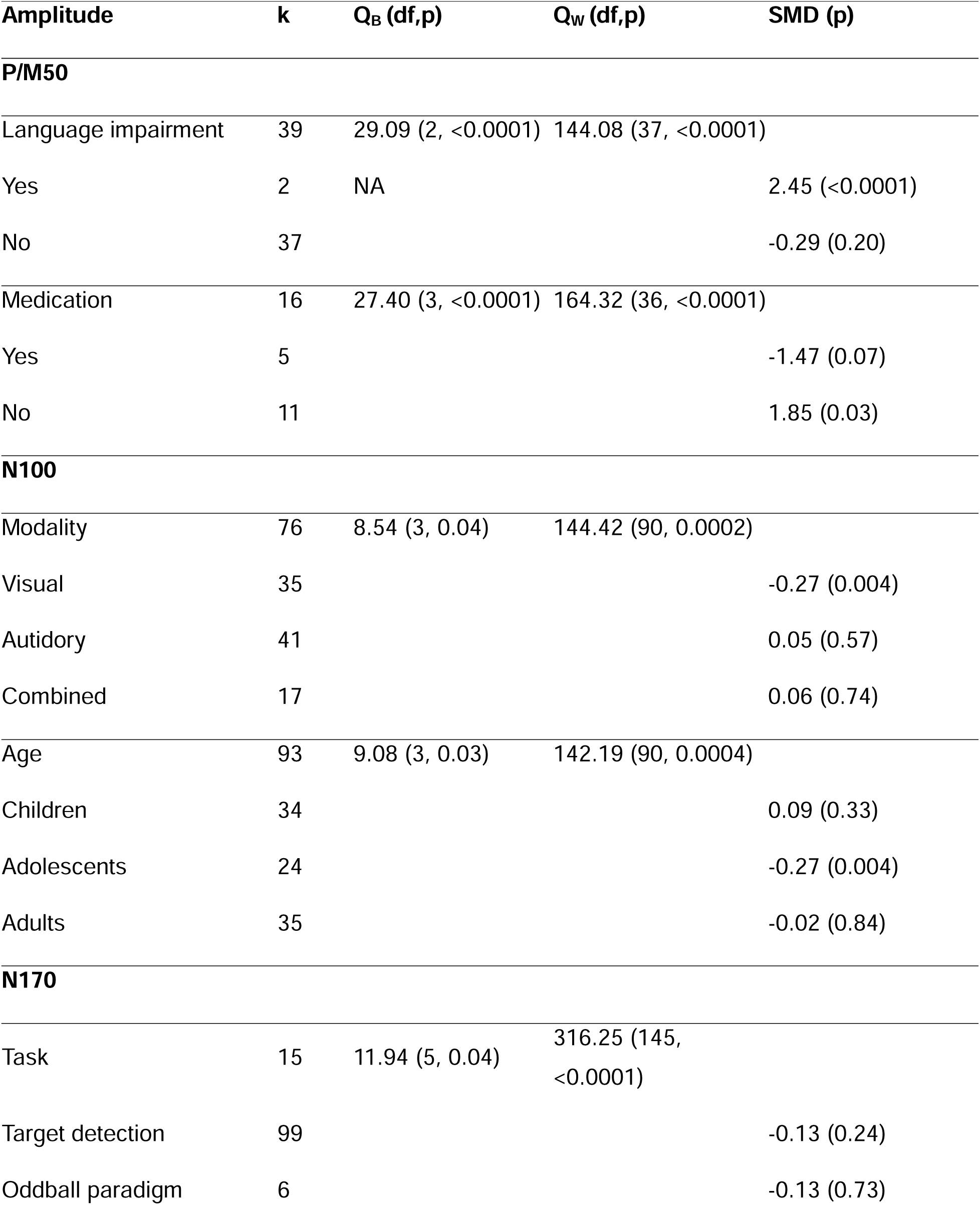
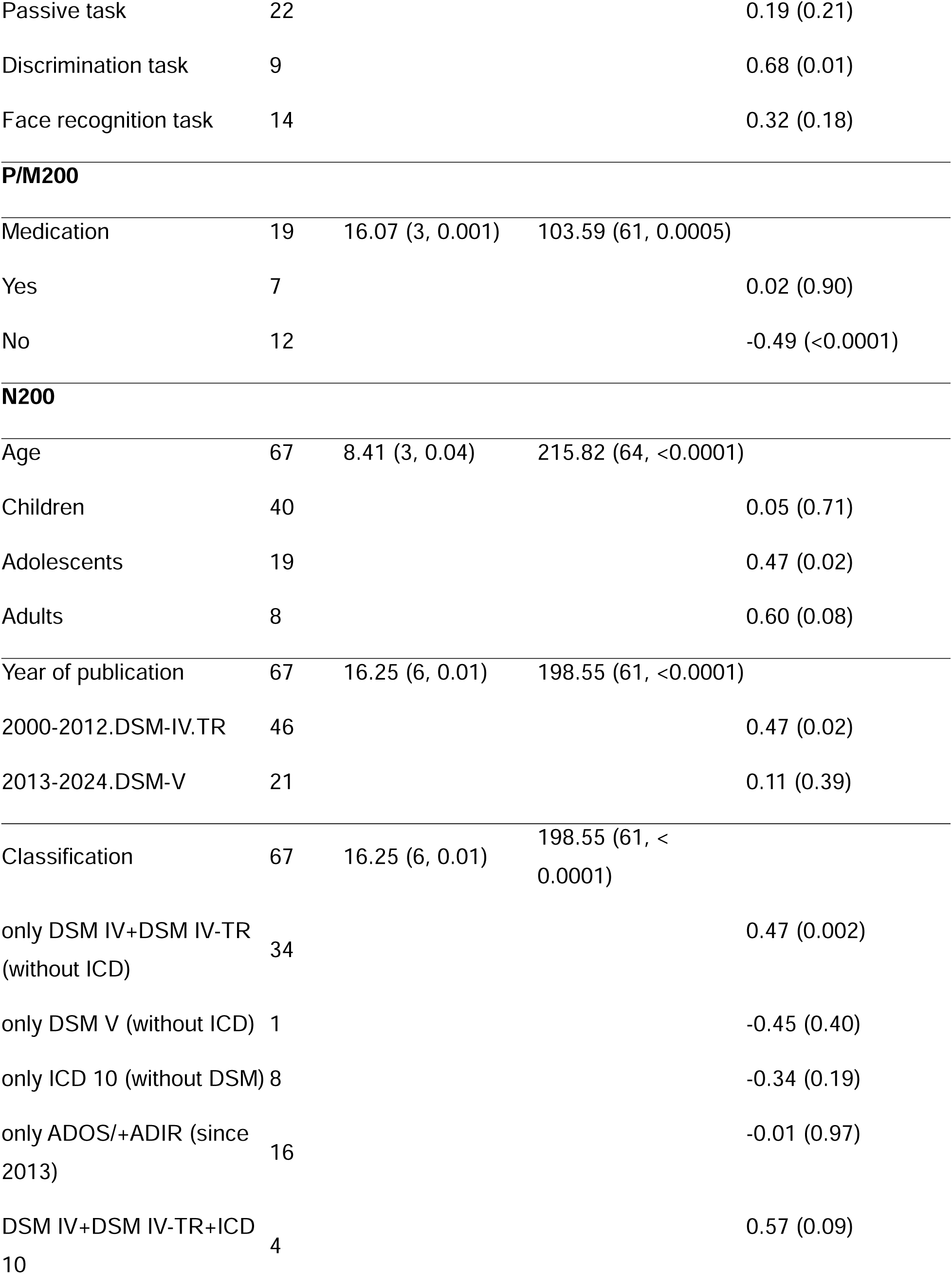
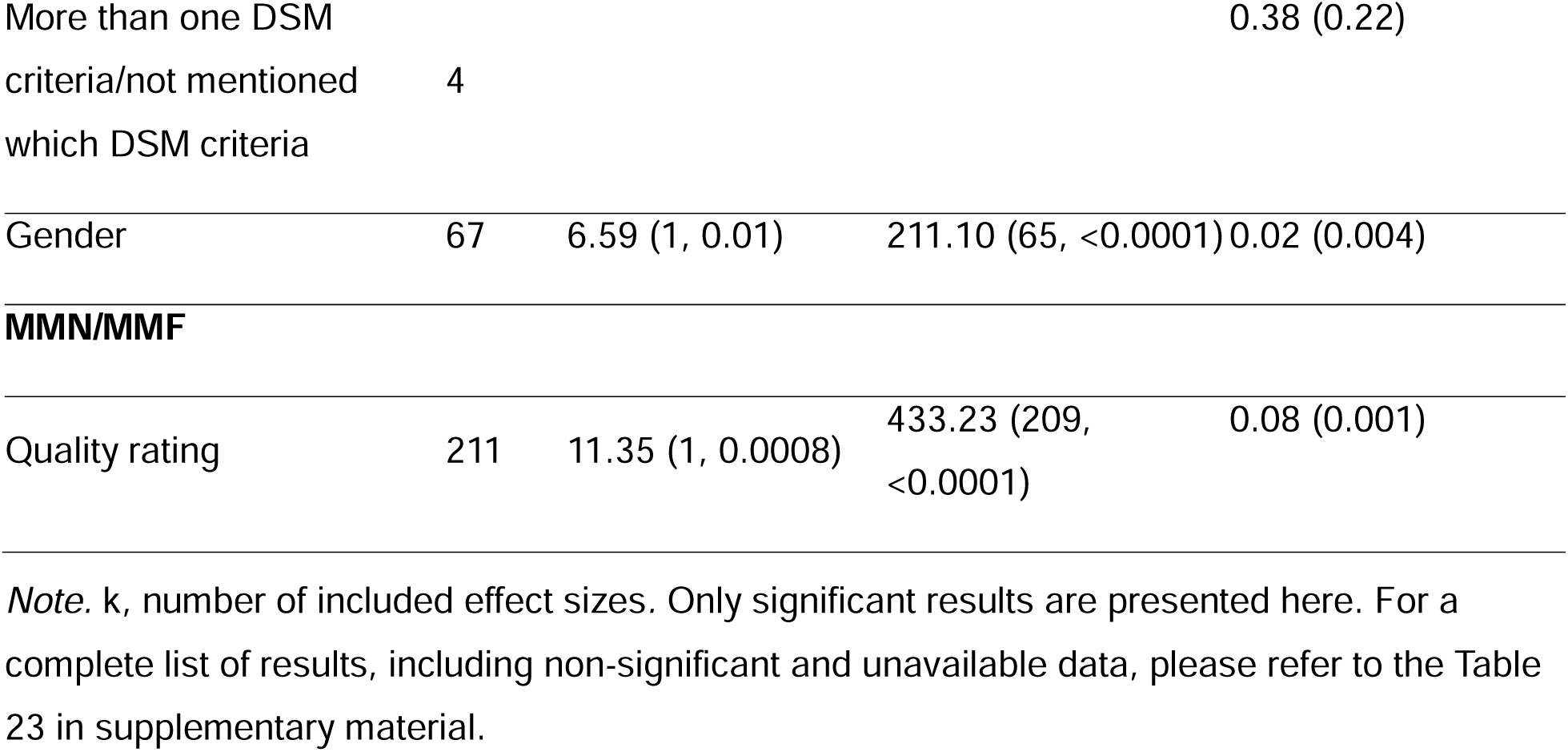
Summary of significant meta-analytic findings for amplitude moderator analyses (mixed-effects models fitted) – P/M50, P/M100, N100, N170, P/M200, N200, MMN/MMF.

**Table 4:**
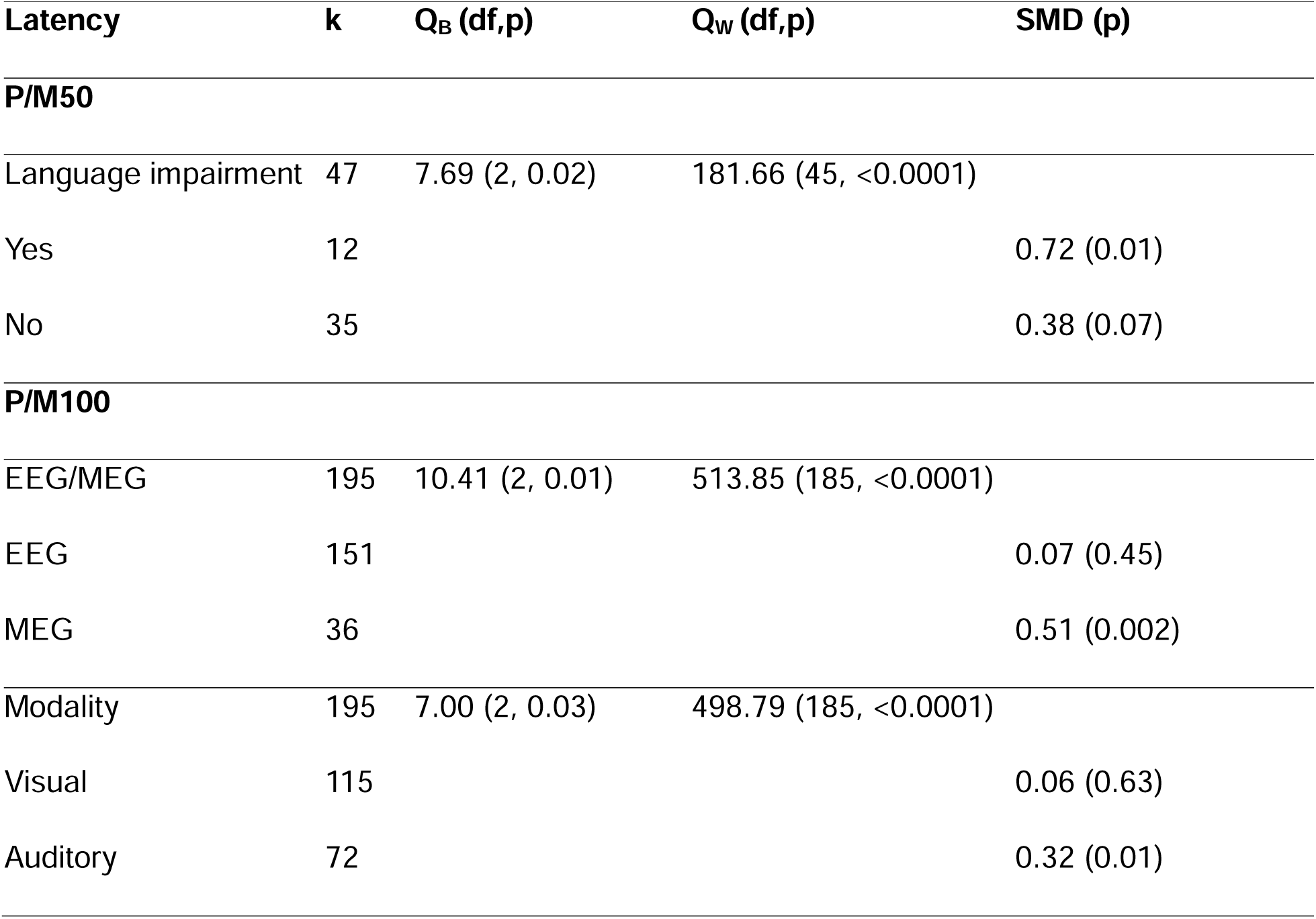
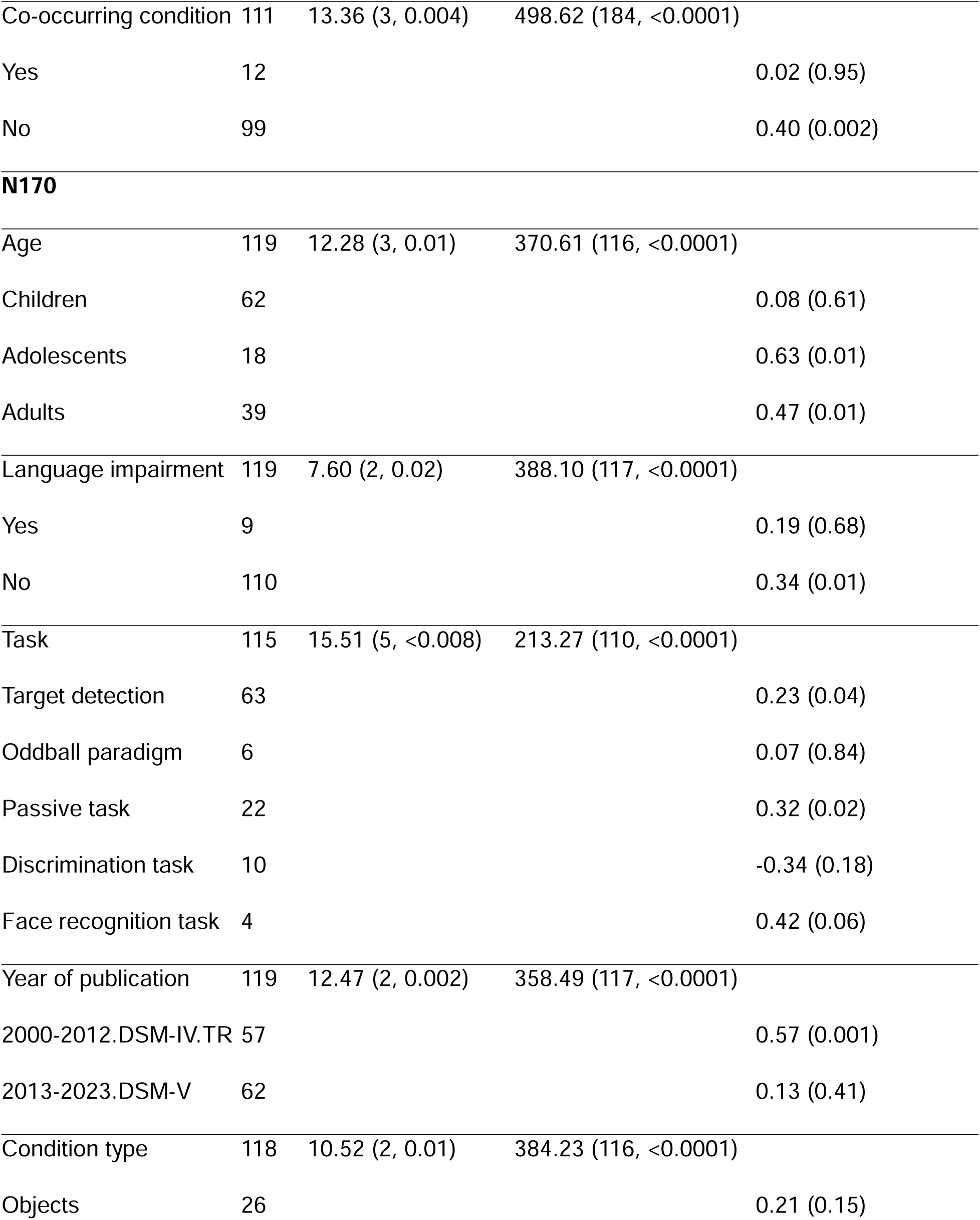
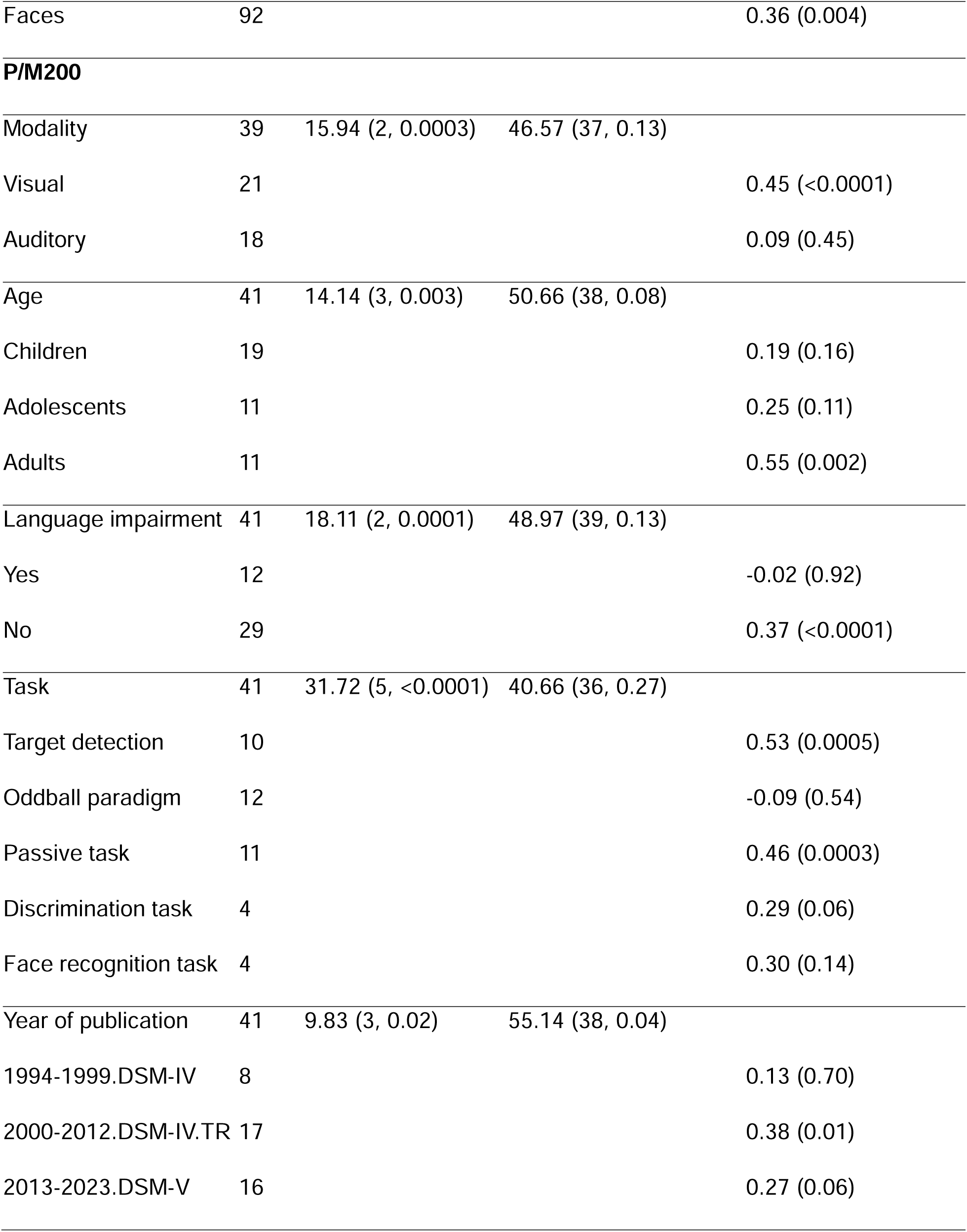
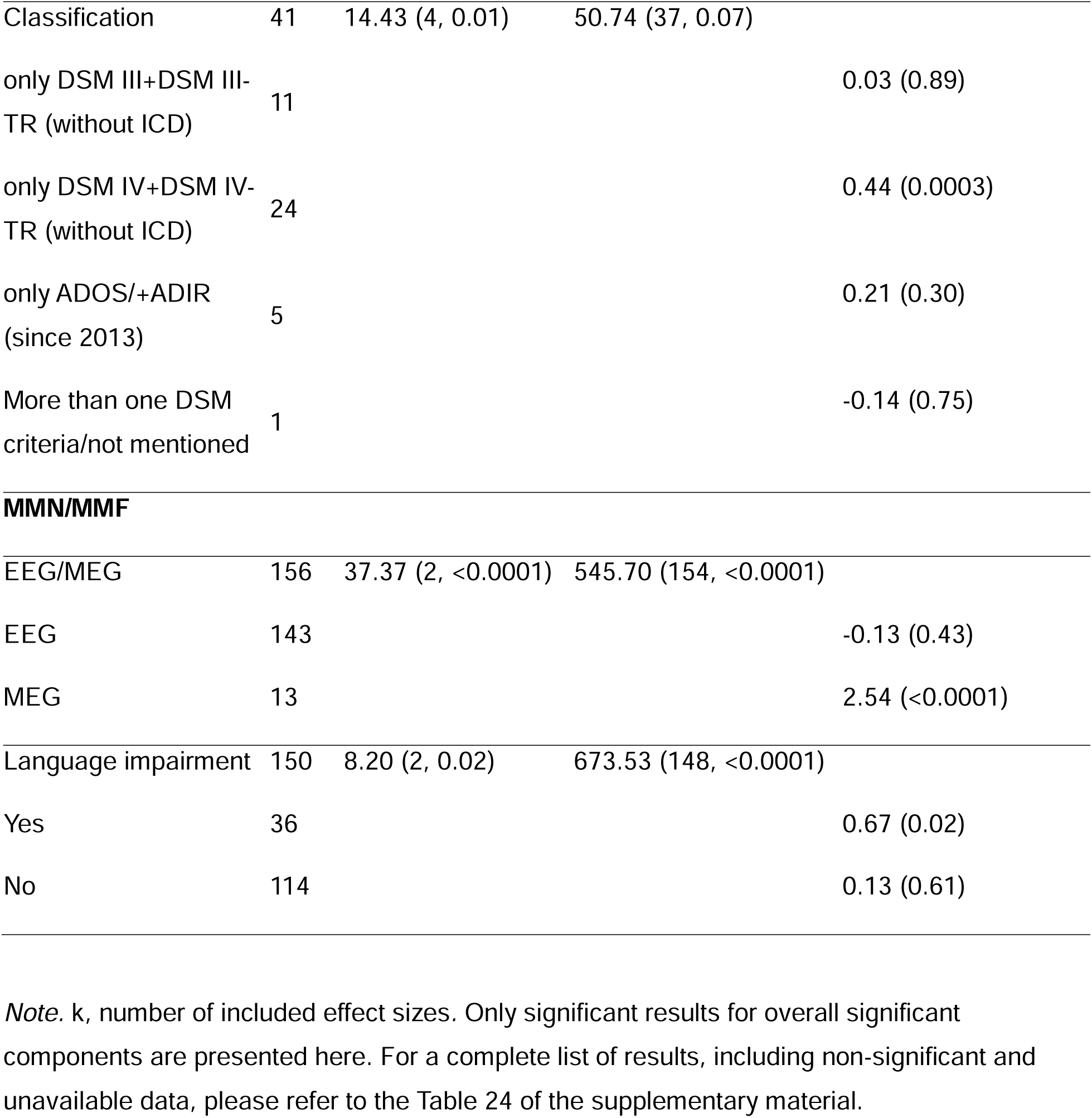
Summary of significant meta-analytic findings for latency moderator analyses (mixed-effects models fitted) – P/M50, P/M100, N100, N170, P/M200, N200, MMN/MMF.

| Latency | k | Q<sub>B</sub> (df,p) | Q<sub>W</sub> (df,p) | SMD (p) |
| --- | --- | --- | --- | --- |
| **P/M50** |  |  |  |  |
| Language impairment | 47 | 7.69 (2, 0.02) | 181.66 (45, <0.0001) |  |
| Yes | 12 |  |  | 0.72 (0.01) |
| No | 35 |  |  | 0.38 (0.07) |
| **P/M100** |  |  |  |  |
| EEG/MEG | 195 | 10.41 (2, 0.01) | 513.85 (185, <0.0001) |  |
| EEG | 151 |  |  | 0.07 (0.45) |
| MEG | 36 |  |  | 0.51 (0.002) |
| Modality | 195 | 7.00 (2, 0.03) | 498.79 (185, <0.0001) |  |
| Visual | 115 |  |  | 0.06 (0.63) |
| Auditory | 72 |  |  | 0.32 (0.01) |

| **Latency** | **k** | **Q<sub>B</sub> (df,p)** | **Q<sub>W</sub> (df,p)** | **SMD (p)** |
| --- | --- | --- | --- | --- |
| Co-occurring condition | 111 | 13.36 (3, 0.004) | 498.62 (184, <0.0001) |  |
| Yes | 12 |  |  | 0.02 (0.95) |
| No | 99 |  |  | 0.40 (0.002) |
| **N170** |  |  |  |  |
| Age | 119 | 12.28 (3, 0.01) | 370.61 (116, <0.0001) |  |
| Children | 62 |  |  | 0.08 (0.61) |
| Adolescents | 18 |  |  | 0.63 (0.01) |
| Adults | 39 |  |  | 0.47 (0.01) |
| Language impairment | 119 | 7.60 (2, 0.02) | 388.10 (117, <0.0001) |  |
| Yes | 9 |  |  | 0.19 (0.68) |
| No | 110 |  |  | 0.34 (0.01) |
| Task | 115 | 15.51 (5, <0.008) | 213.27 (110, <0.0001) |  |
| Target detection | 63 |  |  | 0.23 (0.04) |
| Oddball paradigm | 6 |  |  | 0.07 (0.84) |
| Passive task | 22 |  |  | 0.32 (0.02) |
| Discrimination task | 10 |  |  | -0.34 (0.18) |
| Face recognition task | 4 |  |  | 0.42 (0.06) |
| Year of publication | 119 | 12.47 (2, 0.002) | 358.49 (117, <0.0001) |  |
| 2000-2012.DSM-IV.TR | 57 |  |  | 0.57 (0.001) |
| 2013-2023.DSM-V | 62 |  |  | 0.13 (0.41) |
| Condition type | 118 | 10.52 (2, 0.01) | 384.23 (116, <0.0001) |  |
| Objects | 26 |  |  | 0.21 (0.15) |
| Faces | 92 |  |  | 0.36 (0.004) |
| **P/M200** |  |  |  |  |
| Modality | 39 | 15.94 (2, 0.0003) | 46.57 (37, 0.13) |  |
| Visual | 21 |  |  | 0.45 (<0.0001) |
| Auditory | 18 |  |  | 0.09 (0.45) |
| Age | 41 | 14.14 (3, 0.003) | 50.66 (38, 0.08) |  |
| Children | 19 |  |  | 0.19 (0.16) |
| Adolescents | 11 |  |  | 0.25 (0.11) |
| Adults | 11 |  |  | 0.55 (0.002) |
| Language impairment | 41 | 18.11 (2, 0.0001) | 48.97 (39, 0.13) |  |
| Yes | 12 |  |  | -0.02 (0.92) |
| No | 29 |  |  | 0.37 (<0.0001) |
| Task | 41 | 31.72 (5, <0.0001) | 40.66 (36, 0.27) |  |
| Target detection | 10 |  |  | 0.53 (0.0005) |
| Oddball paradigm | 12 |  |  | -0.09 (0.54) |
| Passive task | 11 |  |  | 0.46 (0.0003) |
| Discrimination task | 4 |  |  | 0.29 (0.06) |
| Face recognition task | 4 |  |  | 0.30 (0.14) |
| Year of publication | 41 | 9.83 (3, 0.02) | 55.14 (38, 0.04) |  |
| 1994-1999.DSM-IV | 8 |  |  | 0.13 (0.70) |
| 2000-2012.DSM-IV.TR | 17 |  |  | 0.38 (0.01) |
| 2013-2023.DSM-V | 16 |  |  | 0.27 (0.06) |
| Classification | 41 | 14.43 (4, 0.01) | 50.74 (37, 0.07) |  |
| only DSM III+DSM III-TR (without ICD) | 11 |  |  | 0.03 (0.89) |
| only DSM IV+DSM IV-TR (without ICD) | 24 |  |  | 0.44 (0.0003) |
| only ADOS/+ADIR<br>(since 2013) | 5 |  |  | 0.21 (0.30) |
| More than one DSM<br>criteria/not mentioned | 1 |  |  | -0.14 (0.75) |
| **MMN/MMF** |  |  |  |  |
| EEG/MEG | 156 | 37.37 (2, <0.0001) | 545.70 (154, <0.0001) |  |
| EEG | 143 |  |  | -0.13 (0.43) |
| MEG | 13 |  |  | 2.54 (<0.0001) |
| Language impairment | 150 | 8.20 (2, 0.02) | 673.53 (148, <0.0001) |  |
| Yes | 36 |  |  | 0.67 (0.02) |
| No | 114 |  |  | 0.13 (0.61) |

*Note.* k, number of included effect sizes. Only significant results for overall significant components are presented here. For a complete list of results, including non-significant and unavailable data, please refer to the Table 24 of the supplementary material.

The effect-sizes for the autistic relative to the non-autistic group are described below:

#### 3.3.1. Type of measurement (EEG/MEG)

A significant moderator-effect was found for P/M100-latency (Q_B_(2)=10.41, p=0.01) and MMN/MMF-latency (Q_B_(2)=37.37, p<0.0001) for MEG-studies only, revealing longer latencies in autism.

#### 3.3.2. Modality of sensory processing (auditory/visual/tactile/multisensory)

A significant moderator-effect revealed a larger mean effect size for N100-amplitude (Q_B_(3)=8.54, p=0.04) and longer P/M200-latency (Q_B_(3)=20.54, p=0.0001) for visual tasks only and longer P/M100-latency (Q_B_(2)=7.00, p=0.03) for auditory tasks only.

#### 3.3.3. Age groups (children/adolescences/adults)

A larger mean effect size was found for N100-amplitude (Q_B_(3)=9.08, p=0.03) and a smaller mean effect size was found for N200-amplitude (Q_B_(3)=8.41, p=0.04) in autistic adolescents only, longer N170-latency (Q_B_(3)=12.28, p=0.01) was found in autistic adolescents and adults only and longer P/M200-latency (Q_B_(3)=14.14, p=0.003) in autistic adults only.

#### 3.3.4. Language impairment (yes/no/unknown)

A significant moderator-effect revealed a larger mean effect size for P/M50-amplitude (Q_B_(2)=29.09, p<0.0001), longer P/M50-latency (Q_B_(2)=7.69, p=0.02) and longer MMN/MMF-latency (Q_B_(2)=8.20, p=0.02) in autistic participants with language impairment only and longer N170-latency (Q_B_(2)=7.60, p=0.02) and P/M200-latency (Q_B_(2)=18.11, p=0.0001) in autistic participants without language impairment only.

#### 3.3.5. Task type (target-detection/oddball-paradigm/passive-task/discrimination-task/coherent motion-task/face-recognition-task/other(unknown))

A smaller effect size was obtained for N170-amplitude (Q_B_(6)=20.50, p=0.002) for discrimination tasks only, longer N170-latency (Q_B_(6)=59.63, p<0.0001) and longer P/M200-latency (Q_B_(5)=31.72, p<0.0001) were observed in target-detection and passive tasks only.

#### 3.3.6. Year of publication (1980-1986/1987-1993/1994-1999/2000-2012/2013-2023)

Longer N170-latency (Q_B_(2)=12.47, p=0.002) and P/M200-latency (Q_B_(3)=9.83, p=0.02) were found in studies published between 2000-2012.

#### 3.3.7. Co-occurring conditions in autism group (yes/no/unknown)

Longer P/M100-latency (Q_B_(3)=13.36, p=0.004) was seen in participants without co-occurring conditions.

#### 3.3.8. Medication in autism group

For P/M50-amplitude (Q_B_(3)=27.40, p<0.001), autistic individuals taking medication showed a smaller effect size, whereas non-medicated autistic individuals exhibited a larger effect size. A smaller effect size was observed for P/M200-amplitude (Q_B_(3)=16.07, p=0.001) in non-medicated autistic individuals only.

#### 3.3.9. Classification system used for diagnosis (for grouping, see eMethods)

Significant moderator-effect was found for a smaller N200-amplitude (Q_B_(6)=16.25, p=0.01) and longer P/M200-latency (Q_B_(4)=14.43, p=0.01) when the diagnosis was based on DSM-IV/DSM-IV-TR only.

#### 3.3.10. IQ

No significant moderator effects were found.

#### 3.3.11. Sex

A smaller effect size was found in N200-amplitude (Q_B_(1)=6.59, p=0.01) in males only.

#### 3.3.12. Condition type (faces/objects)

Longer N170-latency (Q_B_(2)=10.52, p=0.01) was observed for faces only.

### 3.4. Quality assessment analyses

Quality assessment yielded significant results for MMN/MMF-amplitude (SMD=0.08). For details see eResults in supplementary material.

### 3.5. Sensitivity analyses

Sensitivity analyses, excluding outliers, revealed additional significant results for N200-amplitude (SMD=0.24), while P/M100-latency results became non-significant (SMD=0.10). Complete results are available in the eTable 18+19 in the supplement.

### 3.6. Publication bias analyses

Funnel plot asymmetry tests revealed potential publication bias for: P/M100-amplitude, N170-amplitude, P/M50-latency, N200-latency, and MMN/MMF-latency. Trim-and-fill analysis identified 18 missing studies for N200-amplitude and 15 for MMN/MMF-amplitude, but these adjustments did not affect their non-significant results. For P/M100-amplitude, the effect-size became significant (−0.15 to −0.07), while N170-amplitude shifted significantly in the opposite direction (−0.13 to 0.57). All funnel plots are provided in eFigures 11-17.

## 4. Discussion

### 4.1. Early ERPs and ERFs as potential brain-based biomarkers for autism

Sensory processing alterations are a key aspect of autism with growing recognition (Robertson & Baron-Cohen, 2017), affecting functioning and daily-life quality of autistic people (Yela-González et al., 2021). This meta-analysis provides insights into the neurophysiological mechanisms and timings associated with sensory processing in autism by quantitatively synthesizing EEG and MEG studies comparing sensory processing in autistic and non-autistic children, adolescents, and adults and by exploring demographic and methodological moderators influencing between-group differences. Significant group-level differences between autistic and non-autistic individuals were found for the latencies of four ERP/ERF components: P/M50, P/M100, N170, and P/M200. As anticipated, substantial heterogeneity was observed and addressed by moderator-analysis.

With a moderate effect-size the earliest measured ERP/ERF component, the P/M50 yielded the most substantial alteration, indicating longer latencies in autistic individuals. The P/M50 is linked to filtering irrelevant stimuli and early sensory gating (Pratt, 2011) and has been prominently studied as a potential biomarker for schizophrenia, where larger gating ratios and smaller amplitude differences reflect impaired sensory filtering of irrelevant stimuli (Bramon et al., 2004; Freedman et al., 2020; Shen et al., 2020). In autism, this meta-analysis highlights delayed P/M50-latencies, not amplitudes, suggesting slower sensory processing rather than reduced filtering capacity. These delays may hinder real-time comprehension and lead to sensory overwhelm, potentially causing autistic individuals to avoid overstimulating situations (Kootz et al., 1982). Longer P/M50-latencies in autism may also suggest slower arousal and processing of new stimuli, making it harder to cope with unexpected or changing auditory input in everyday situations. Moderator analysis showed even greater delays in individuals with co-occurring language impairment, leading to a notably large effect-size. This positions the P/M50 as a valuable biomarker-candidate for autistic individuals, especially with language impairment. Since language impairment is highly prevalent among autistic people (Schaeffer et al., 2023), and affects academic, social, and adaptive skills (McKernan & Kim, 2022), these delays may reflect underlying mechanisms to these alterations. With eleven of the fourteen studies including auditory tasks and the strong association between language impairment and auditory processing (Linke et al., 2017; Richards & Goswami, 2018), auditory support in autistic individuals may provide significant benefits. Sensitivity analysis revealed slight publication bias in P/M50-latency, suggesting a lower true effect-size.

Prolonged P/M100-latencies in autism may reflect delayed attention to sensory inputs, including spatial information, indicating altered arousal regulation (Key et al., 2005; Luck et al., 2000). These alterations were particularly evident in auditory tasks, although as a trend, with longer latencies in autistic individuals with language impairments. As with P/M50, the link between language impairment and auditory processing suggests sensory delays may contribute to communication alterations in autism. Since P/M100, like P/M50, is associated with auditory sensory gating (Key et al., 2005), this result further supports the value of auditory support for autistic individuals with language impairment and sensory delays. Interestingly, autistic individuals without co-occurring conditions showed longer P/M100-latencies. ADHD, among the most common co-occurring condition in autism, is associated with shorter P100-latencies (Kaiser et al., 2020), which could partly explain this finding. The shift in P/M100-latency-effects after outlier removal highlights the need for cautious interpretation.

The N170-component is linked to face processing (Hinojosa et al., 2015). This meta-analysis confirms previous findings of prolonged N170-latency in autistic individuals (Kang et al., 2018; Mason et al., 2022), showing greater delays for faces than objects. These delays may contribute to altered social behaviour, as face processing is closely tied to social interaction (Corbett et al., 2014). These findings highlight the potential to target face processing and social cognition in autism, which may improve social engagement and interactions. Moderator analysis revealed significant delays in autistic adolescents and adults but not children, suggesting slower face processing becomes more pronounced with age. A possible explanation is that autistic individuals from an early age spend less time engaged in face-to-face interactions or eye contact (Chawarska et al., 2013; W. Jones & Klin, 2013; Shic et al., 2014) impacting the development of brain regions otherwise specialised for face processing (Pierce et al., 2001). This highlights the importance of examining developmental changes in autism, as social cognitive alterations may be less apparent in early childhood but more measurable and diagnostically relevant with age. Thus, the N170 may be a valuable biomarker for social cognitive processing in autistic adolescents and adults. Alterations were found in both target-detection and passive tasks, though limited comparative studies constrain conclusions. Prolonged N170-latency was also noted in autistic individuals without language impairments, but again the limited number of studies constrains generalisability. The effect-size for N170 is smaller than that for P/M50, suggesting early sensory processing alterations in P/M50 may influence later face processing reflected in N170.

Furthermore, this meta-analysis showed prolonged P/M200-latencies in autistic individuals. Among others, P/M200 is linked to the processing of facial configuration (Schweinberger & Neumann, 2016) and decoding of verbal emotional expressions (Kotz & Paulmann, 2011; Paulmann & Kotz, 2008). Moderator analysis revealed these delays were significant in visual tasks, reflecting slower processing of social and emotional cues, which may contribute to challenges in social interaction and emotional understanding. Significant delays were observed in autistic adults specifically. Similar to the N170-component, this may be due to maturational processes and differing social experiences over time. Delays were noted in both, passive and target-detection tasks, and for autistic individuals without language impairment. However, cautious interpretation is warranted due to limited evidence.

Although some studies reported amplitude alterations (Farashi et al., 2023; Gonçalves & Monteiro, 2023), this meta-analysis unexpectedly found none. Following Dinstein et al. 2015, one potential explanation may be that greater amplitude variability in autistic individuals might obscure group-level alterations (Dinstein et al., 2015). This would be supported by quality assessment, sensitivity, and publication bias analyses conducted in our study, which revealed that several previously non-significant amplitude results became significant once these methodological factors were accounted for. However, this meta-analysis rather implies that while neural response magnitude may be similar across groups, response timing, reflected in latency alterations, may be a more robust marker for distinguishing autistic individuals.

Whenever differences between EEG and MEG were found in the moderator analysis, significant effect-sizes were observed only for MEG, suggesting it more reliably detects sensory alterations in early ERP/ERF components. Significant results in autism were observed only in auditory and visual tasks, suggesting core alterations in these domains. No alterations were found in tactile or multisensory tasks. However, the limited number of studies (2 for tactile and 5 for multisensory) allows only tentative conclusions. No eligible studies examined olfactory or gustatory functions, highlighting the need for future research in these areas. Significant differences between autistic and non-autistic individuals appeared in discrimination, target-detection, and passive tasks. However, k-values for other tasks were significantly smaller and often too small for reliable comparisons. Beyond P/M100-latency, co-occurring conditions had no effect on other components. Medication, sex and IQ had no moderator effect on overall significant components. However, medication and sex had moderator effects on not overall significant components (P/M50-, P/M200- and N200-amplitude, respectively, for details see supplement). Significant results in the quality assessment analysis of MMN/MMF-amplitudes suggest potential limitations in data reliability for this component, highlighting the need for cautious interpretation. The emergence of significant N200-amplitude differences after outlier removal suggests that extreme data points may have masked group differences in this component. Publication bias analysis revealed significant impacts on detecting effects for P/M100- and N170-amplitudes. For P/M100, the shift to significance suggests earlier analyses slightly underestimated its effect. For N170, a large shift from non-significance to significance indicates that pronounced bias obscured meaningful effects. Further exploration of these amplitudes is warranted.

### 4.2. Practical implications

ERPs/ERFs provide a non-invasive, cost-effective method to detect sensory processing alterations complementing self-reports or caregiver accounts and are generally easy to measure in clinical settings. P/M50-latencies in individuals with language impairment and N170-latencies in adolescents and adults could serve as potential biomarker-candidates. These neural markers could be used as diagnostic aid for early detection of sensory processing alterations.

Furthermore, this evidence suggests that subgroups of autistic individuals may be identifiable based on alterations in P/M50 and N170, with P/M50 linked to language impairments and N170 observed in adolescents and adults addressing more social cognition alterations. This may not only enhance diagnostic precision but also guide more personalised support, better aligned with the sensory characteristics of these subgroups.

However, while objectively measuring sensory issues, ERPs/ERFs can only reveal sensory processing alterations on a neurophysiological level, which may not always align with sensory experiences described by autistic individuals or reported by caregivers. Additionally, the observed low effect-sizes and substantial heterogeneity limit the practical application as reliable biomarkers in clinical settings and routine diagnostics. Furthermore, studies are needed assessing the specificity of those markers and further exploring their value in clinical practice, enhancing and expanding traditional assessments.

### 4.3. Limitations & future directions

Availability of studies varied substantially between ERP/ERF components and was too scarce for analysis in some cases. Similarly, the number of studies within each subgroup for moderator analysis varied considerably. Further, varying instruments assessing the degree of autism expression and sensory features in autistic individuals prohibited the exploration of these factors as moderators. Insufficient reporting of co-occurring conditions and medication, along with the exclusion of individuals with intellectual disabilities further limited analyses and generalizability. Finally, due to the imbalance in the sex of the participants and the fact that most studies in this field are conducted in Western countries, the results of this study may not be fully generalizable to female and other underrepresented participants or to populations outside Western contexts. Future studies should aim for a more balanced gender distribution and ensure inclusion of participants from diverse cultural and ethnic backgrounds, report co-occurring conditions and medications, include individuals with intellectual disabilities, and use standardized tools for degree of autism expression and questionnaires for sensory perception, to offer a more comprehensive understanding of sensory variations within the autistic community. They should further explore how sensory processing alterations affect emotional regulation, adaptive behaviour, and distress, offering insights into targeted support strategies in autism.

### 4.4. Conclusions

This meta-analysis found prolonged P/M50-, P/M100-, N170-, and P/M200-latencies as potential biomarker-candidates for sensory processing alterations in autism. Prolonged P/M50-latency, reflecting sensory filtering challenges, could guide future studies on early autism detection, especially in individuals with language impairments. N170-latency findings suggest social cognition difficulties may not be evident in children but become more detectable in adolescents and adults, with small effect-sizes. However, low effect-sizes, heterogeneity, and limited number of studies constrain practical applications, warranting further research.

#### Language

We adhered to the language guidelines proposed by AIMS-2-Trials, ensuring consistent and respectful terminology when describing autistic and non-autistic individuals(Eliza Eaton for AIMS-2-TRIALS, n.d.).

## Data Availability

All data produced in the present study are available upon reasonable request to the authors

