## Supplementary material for "Neurophysiological Alterations during Sensory Processing in Autism - A Meta-Analysis"

### **eMethods**

#### **Search strategy**

PubMed

("autism spectrum disorder"[MeSH Terms] OR "autism s"[All Fields] OR "autisms"[All Fields] OR "autistic disorder"[MeSH Terms] OR ("autistic"[All Fields] AND "disorder"[All Fields]) OR "autistic disorder"[All Fields] OR "autism"[All Fields]) AND ("electroencephalography"[MeSH Terms] OR "electroencephalography"[All Fields] OR "eeg"[All Fields] OR "evoked potentials"[MeSH Terms] OR "magnetoencephalography"[MeSH Terms] OR "magnetoencephalography"[All Fields] OR "meg"[All Fields]) AND ("perception"[MeSH Terms] OR "perception"[All Fields] OR ("sensory"[All Fields] AND "processing"[All Fields]) OR "sensory"[All Fields])

PsycINFO

(MAINSUBJECT.EXACT("Autism Spectrum Disorders") OR "autism s" OR autisms OR "autistic disorder" OR autism) AND (MAINSUBJECT.EXACT("Electroencephalography") OR electroencephalography OR eeg OR MAINSUBJECT.EXACT("Evoked Potentials") OR MAINSUBJECT.EXACT("Magnetoencephalography") OR magnetoencephalography OR meg) AND (MAINSUBJECT.EXACT("Perception") OR perception OR (sensory AND processing) OR sensory)

Web Of Science

(ALL=("autism spectrum disorder") OR ALL=("autism s") OR ALL=(autisms) OR ALL=("autistic disorder") OR ALL=(autism)) AND (ALL=(electroencephalography) OR ALL=(eeg) OR ALL=("evoked potentials") OR ALL=(magnetoencephalography) OR ALL=(meg)) AND (ALL=(PERCEPTION) OR ALL=(SENSORY))

Clinical Trials Register

("autism s"[All Fields] OR "autisms"[All Fields] OR "autistic disorder"[MeSH Terms] OR ("autistic"[All Fields] AND "disorder"[All Fields]) OR "autistic disorder"[All Fields] OR "autism"[All Fields]) AND (("electroencephalography"[MeSH Terms] OR "electroencephalography"[All Fields] OR "eeg"[All Fields]) OR "meg"[All Fields]) AND ("perception"[MeSH Terms] OR "perception"[All Fields] OR ("sensory"[All Fields] AND "processing"[All Fields]) OR "sensory processing"[All Fields])

#### **Data coding and data extraction**

The coding and part of the data extraction were performed using the Covidence^1^ software.

Study type/reference

- First author
- Year of publication
- Peer-reviewed journal
- Study quality rating (QR)^[1]^

Autistic group

- Sample size. n
- Mean age. years

1 child (< 12 years)

2 adolescent (12 – 17.99 years)

3 adult (≥ 18 years)

- Age range. years
- Handerness

1 right

2 left

3 ambidextrous

- Male. %
- Inteligence Quotient (IQ)
- Verbal IQ
- Non-verbal IQ
- Degree of autism expression (symptom severity questionnaire)
- Degree of autism expression (symptom severity score)
- Education. years
- Socioeconomical level
- Co-occuring condition

1 yes

2 no

3 no information

- Type of co-occuring condition
- Number of participants with co-occuring conditions
- Medication status

1 yes

2 no

3 no information

- Type of medication
- Number of participants medicated. %
- Socioeconomic status
- Classification system used for diagnosis

1 only DSM III+DSM III-TR (without ICD)

2 only DSM IV+DSM IV-TR (without ICD)

3 only DSM V (without ICD)

4 only ICD 10 (without DSM)

5 only ADOS/+ADIR (until year 2012)

6 only ADOS/+ADIR (since 2013)

7 DSM III+DSM III-TR+ICD 10

8 DSM IV+DSM IV-TR+ICD 10

9 DSM V+ICD 10

10 More than one DSM criteria/not mentioned which DSM criteria

- Pervasive developmental disorder (PDD)

1 yes

2 no

3 no information

- Language impairment

1 yes

2 no

3 only some participants with language impairment

- Sensory difficulties

1 mentioned

2 not mentioned

- Sensory profile
- 1 Low registration

2 Sensory seeking

3 Sensory sensitivity

4 Sensory avoidance

5 Total

6 Hyporesponsive

7 Hyperresponsive

8 Auditory item score

9 Touch score

Non-autistic group

- Sample size. n
- Mean age. years

1 child (< 12 years)

2 adolescent (12 – 17.99 years)

3 adult (≥ 18 years)

- Age range. years
- Handerness

1 right

2 left

3 ambidextrous

- Male. %
- IQ
- Verbal IQ
- Non-verbal IQ
- Education. years
- Socioeconomical level
- Co-occuring condition

1 yes

2 no

3 no information

- Type of co-occuring condition
- Number of participants with co-occuring conditions
- Medication status

1 yes

2 no

3 no information

- Type of medication
- Number of participants medicated. %
- Socioeconomic status
- Sensory difficulties

1 mentioned

2 not mentioned

- Sensory profile
- 1 Low registration

2 Sensory seeking

3 Sensory sensitivity

4 Sensory avoidance

5 Total

6 Hyporesponsive

7 Hyperresponsive

8 Auditory item score

9 Touch score

Moderators

- Type of measurement

1 EEG

2 MEG

- EEG/MEG system (type)
- Commercial EEG/MEG system (type)

1 yes

2 no

- Type of stimuli
- Task type category

1 Target detection

2 Oddball paradigm

3 Passive task

4 Discrimination task

5 Coherent motion task

6 Other tasks/not mentioned

7 Face recognition task

- Modality

1 visual

2 auditory

3 tactile

4 visual-auditory (multimodal)

5 visual-tactile (multimodal)

6 auditory-tactile (multimodal)

- Latency range
- Interstimulus intervals. ms
- Electrodes (type)

1 Ag/Ag-Cl electrodes

2 No information

3 Other

- Sensors (types)

1 Magnetometers

2 Gradiometers

3 No information

4 Other

- Number of total electrodes (chanels)/sensors
- Name of analyzed electrodes

1 Fz

2 Fcz

3 Cz

4 Cpz

5 Oz

6 Pz

7 Poz

8 Other

- Central midline electrodes assessed (only for EGG)

1 yes

2 no

- Region of Interest (only for MEG)

1 left anterior

2 right anterior

3 left posterior

4 right posterior

5 left occipital

6 right occipital

7 left hemisphere

8 right hemisphere

9 Other

- Region of Interest specific (only for MEG)
- Number of trials to compute average/ERPs
- Reference channel
- Impedance (only for EEG). Kohms
- Electrooculogram recorded

1 yes

2 no

- Eye movement correction

1 yes

2 no

- Artifact rejection criterion. uV
- Interpolation of noisy channels

1 yes

2 no

- Baseline correction

1 yes

2 no

- Filter range offline. Hz
- Montage/system (only for EEG)
- Reference (only for EEG)

1 Appropiate (Cz. FCz. vertex. mastoids....)

2 Other/inappropriate

- Number of coils (only MEG)
- Measure of SNR (only MEG)

ERP/F components

Coding of means (M) and standard deviations (SD) for all ERP/F components: P/M50. P/M100. P/M120. P/M130. P/M200. P/M220. P/M290. P/M300. P/M330. N80. N100. N130. N140. N160. N170. N200. N250. N270. N300. N350. Nc. MMN/MMF. ERP/ ERF. ERN. – amplitudes and latencies. Only those reported in more than 1 study where analyzed.

Performance data

Coding of means (M) and standard deviations (SD) for (behavioral) performance data.

#### **Quality assessment**

Modified version of the Newcastle-Ottawa Scale (NOS)^2^

| **Item and responses** | **Points** |
| --- | --- |
| SELECTION | |
| **1. Is the case definition adequate?** | |
| a) Yes. With independent validation (DSM, ICD, ADOS, ADI-R) | **1** |
| b) Yes. E.g. record linkage or based on self reports | **0** |
| c) No description | **0** |
| **2. Representativeness of the cases** | |
| a) Consecutive or obviously representative series of cases | **1** |
| b) Potential for selection biases not stated | **0** |
| **3. Selection of controls** | |
| a) Community controls | **1** |
| b) Hospital controls | **0** |
| c) No description | **0** |
| **4. Definition of controls** | |
| a) No current (psychiatric) diagnosis | **1** |
| b) No description of source | **0** |
| COMPARABILITY | |
| 1. **Comparability (if matched)** | |
| a) Study controls for age (most importante factor) | **1** |
| b) Study controls for sex and IQ (any additional factors) | **1** |
| EXPOSURE | |
| 1. **Ascertainment of exposure** | |
| a) Secure record | **1** |
| b) Structured interview where blind to case/control status | **1** |
| c) Interview not blinded to case/control status | **1** |
| d) Written self report or medical record only | **0** |
| e) No description | **0** |
| 2. **Same method of ascertainment for cases and controls** | |
| a) Yes | **1** |
| b) No | **0** |

Self-constructed scale for EEG and MEG signal quality

| **Item and responses** | **Points** |
| --- | --- |
| **SIGNAL QUALITY** | |
| **1. Impedance (only EEG)** | |
| a) Below 20 kohms | **1** |
| b) Below 50 kohms | **0** |
| c) Above 50 Kohms | **0** |
| **2. Electrodes/sensors (type) (EEG/MEG)** | |
| a) Ag/Ag-Cl electrodes | **1** |
| b) Magnetometers/Gradiometers | **1** |
| c) Other | **0** |
| **3. Central midline electrodes assessed (only EEG)** | **1** |
| **4. Montage/system (only EEG)** | |
| c) 10-20 Internationl classification | **1** |
| a) Other | **0** |
| **5. Number of trials to compute average/ERPs (EEG/MEG)** | |
| a) minimum of 20 sweeps/trials* | **1** |
| b) < 20 sweeps/trials or no information | **1** |
| **6. Electrooculogram recorded (EEG/MEG)** | **1** |
| **7. Eye movement correction performed (EEG/MEG)** | **1** |
| **8. Artifact rejection criterion (EEG)** | |
| <100 uV | **0** |
| 100 uV | **1** |
| 150 uV | **1** |
| 200 uV | **0** |
| > 200 uV | **0** |
| None | **0** |
| **Other quality control and data preprocessing strategies** | |
| **10a. Interpolation of noisy channels (EEG/MEG)** | **1** |
| **10b. Baseline correction (EEG/MEG)** | **1** |
| **10c. Reference (only for EEG)** | |
| a) Appropriate* (Cz. FCz. vertex. mastoids. average...) | **1** |
| b) Other/inappropriate | **0** |
| **10d. Is a filter range defined? (EEG/MEG)** | **1** |
| **ONLY MEG** | |
| **11. ROI defined** | **1** |
| **12. Number of COILS mentioned** | **1** |
| **9. Artifact rejection criterion mentioned (MEG)** | **1** |
| **Other added items** | |
| **1. Have they defined the component?** | **1** |
| **2. Have they non-significant data published or they send it tous?** | **1** |

### **eResults**

#### **eTable 1. Characteristics of included studies**

| **Study ID** | **Age group** | **n**  **_TOTAL_** | **n_autistic_** | **Age. mean (years)** | **Male. mean (%)** | **IQ. mean** | **Co-occuring condition** | **Medication** | **Language impairment** | **n_non-autistic_** | **Age. mean (years)** | **Male. mean (%)** | **IQ mean** | **Co-occuring condition** | **Medication** | **Type** | **Modality** | **Task type/neuropsychological paradigm** | **TotalQR** |
| --- | --- | --- | --- | --- | --- | --- | --- | --- | --- | --- | --- | --- | --- | --- | --- | --- | --- | --- | --- |
| Abdeltawwab 2015^3^ | children | 61 | 31 | 11.37 | 77.42 | NA | NA | NA | yes | 30 | 11.20 | 60.00 | NA | NA | NA | EEG | auditory | Oddball paradigm | 11 |
| Ahlfors 2024^5^ | adolescences | 53 | 22 | 13.60 | 81.80 | NA | yes | NA | no | 31 | 13.10 | 87.10 | NA | no | NA | MEG | auditory | Passive task | 14 |
| Arnett 2018^6^ | adolescences | 103 | 76 | 12.12 | 80.26 | NA | NA | NA | yes | 27 | 13 | 62.96 | NA | no | NA | EEG | auditory | Passive task | 12 |
| Baruth 2010^7^ | adolescences | 30 | 15 | 13.90 | 86.67 | 92.50 | no | NA | no | 15 | 15.50 | 73.33 | NA | no | NA | EEG | visual | Target detection; Oddball paradigm | 15 |
| Batty 2011^8^ | children | 30 | 15 | 10.55 | 86.67 | NA | NA | NA | yes | 15 | 10.51 | 60.00 | NA | NA | NA | EEG | visual | Target detection | 11 |
| Borgolte 2021^9^ | adults | 29 | 14 | 40.30 | 50.00 | NA | Yes | NA | no | 15 | 42.40 | 60.00 | NA | no | NA | EEG | visual; auditory | Other: perception task | 18 |
| Brennan 2016^10^ | children | 25 | 12 | 9.30 | 91.67 | 98.40 | no | NA | no | 13 | 9.70 | 92.31 | 116.00 | no | NA | MEG | auditory | Passive task | 16 |
| **Study ID** | **Age group** | **n**  **_TOTAL_** | **n_autistic_** | **Age. mean (years)** | **Male. mean (%)** | **IQ. mean** | **Co-occuring condition** | **Medication** | **Language impairment** | **n_non-autistic_** | **Age. mean (years)** | **Male. mean (%)** | **IQ mean** | **Co-occuring condition** | **Medication** | **Type** | **Modality** | **Task type/neuropsychological paradigm** | **TotalQR** |
| Bruneau 1999^11^ | children | 32 | 16 | NA | 75.00 | NA | yes | NA | yes | 16 | NA | 75.00 | NA | no | NA | EEG | auditory | Passive task | 13 |
| Bruneau 2003^12^ | children | 42 | 26 | 5.92 | 84.62 | NA | no | no | yes | 16 | NA | 75.00 | NA | no | no | EEG | auditory | Passive task | 14 |
| Cardy 2005^13^ | children | 16 | 7 | 11.90 | 100.00 | NA | NA | NA | no | 9 | 11.90 | 55.56 | NA | NA | NA | MEG | auditory | Oddball paradigm | 10 |
| Cary 2024^14^ | adolescences | 26 | 13 | 12.81 | 84.62 | 103.69 | no | NA | no | 13 | 12.53 | 46.15 | 115.08 | no | NA | EEG | auditory | Oddball paradigm; Passive task | 15 |
| Čeponiene 2003 ^15^ | children | 19 | 9 | 8.90 | 88.89 | NA | NA | no | yes | 10 | 8.40 | 90.00 | NA | NA | NA | EEG | auditory | Oddball paradigm | 12 |
| Charpentier 2018^16^ | children | 30 | 15 | 10.00 | 86.67 | NA | no | yes | no | 15 | 9.80 | 80.00 | NA | no | NA | EEG | auditory | Oddball paradigm; Passive task | 13 |
| Chen 2021^17^ | children | 48 | 24 | 7.60 | NA | NA | NA | no | no | 24 | 7.46 | NA | NA | NA | no | EEG | auditory | Passive task | 17 |
| Chien 2018^18^ | adults | 72 | 37 | 21.00 | 94.59 | 99.30 | NA | no | no | 35 | 20.50 | 91.43 | 110.60 | no | no | EEG | auditory | Oddball paradigm | 18 |
| Chien 2019^19^ | adults | 68 | 34 | 20.60 | 94.12 | 100.80 | NA | no | no | 34 | 20.40 | 94.12 | 110.50 | NA | no | EEG | auditory | Other: paired-click paradigm | 18 |
| Churches 2012a^21^ | adults | 22 | 11 | 31.82 | 100.00 | 120.18 | no | NA | no | 11 | 30.10 | 100.00 | 116.10 | no | NA | EEG | visual | Discrimination task | 13 |

| **Study ID** | **Age group** | **n**  **_TOTAL_** | **n_autistic_** | **Age. mean (years)** | **Male. mean (%)** | **IQ. mean** | **Co-occuring condition** | **Medication** | **Language impairment** | **n_non-autistic_** | **Age. mean (years)** | **Male. mean (%)** | **IQ mean** | **Co-occuring condition** | **Medication** | **Type** | **Modality** | **Task type/neuropsychological paradigm** | **TotalQR** |
| --- | --- | --- | --- | --- | --- | --- | --- | --- | --- | --- | --- | --- | --- | --- | --- | --- | --- | --- | --- |
| Churches 2012b^22^ | adults | 23 | 10 | 30.61 | 100.00 | 115.20 | no | NA | no | 13 | 29.54 | 100.00 | 118.17 | no | NA | EEG | visual | Target detection | 14 |
| Cléry 2013a^23^ | children | 24 | 12 | 11.58 | 83.33 | 86.00 | no | no | no | 12 | 11.25 | 83.33 | NA | no | no | EEG | visual | Oddball paradigm; Passive task | 16 |
| Cléry 2013b^24^ | adults | 26 | 13 | 26.17 | 84.62 | 89.00 | NA | NA | no | 13 | 24.25 | 61.54 | NA | no | no | EEG | visual | Oddball paradigm; Passive task; Other: distractive task | 15 |
| Constable 2012^25^ | adults | 16 | 9 | 36.60 | 100.00 | 111.00 | NA | NA | no | 7 | 48.90 | 100.00 | 104 | NA | NA | EEG | visual | Passive task; Discrimination task | 11 |
| Cotter 2023^26^ | children | 216 | 84 | 11.40 | 85.70 | 101.50 | NA | no | no | 132 | 11.50 | 46.97 | 109.50 | no | no | EEG | auditory | Simple reaction time task. target detection | 17 |
| Crasta 2021^27^ | children | 36 | 18 | 8.41 | 72.22 | 98.72 | NA | NA | no | 18 | 8.03 | 72.22 | 111.39 | no | NA | EEG | auditory | Passive task; Other: Sensory Gating paradigm | 16 |
| Crasta 2024^28^ | adults | 48 | 24 | 23.31 | 70.80 | NA | NA | NA | no | 24 | 23.70 | 50.00 | NA | no | NA | EEG | auditory | modified paired-click sensory gating EEG paradigm | 15 |
| Dawson 2004^29^ | children | 51 | 29 | 3.73 | 89.66 | NA | no | NA | no | 22 | 3.64 | 86.36 | NA | no | NA | EEG | visual | Passive task; Other: Emotion recognition | 18 |
| **Study ID** | **Age group** | **n**  **_TOTAL_** | **n_autistic_** | **Age. mean (years)** | **Male. mean (%)** | **IQ. mean** | **Co-occuring condition** | **Medication** | **Language impairment** | **n_non-autistic_** | **Age. mean (years)** | **Male. mean (%)** | **IQ mean** | **Co-occuring condition** | **Medication** | **Type** | **Modality** | **Task type/neuropsychological paradigm** | **TotalQR** |
| Day 2024^30^ | adolescences | 126 | 49 | 14.50 | 85.70 | NA | NA | NA | no | 77 | 13.80 | 59.74 | NA | NA | NA | EEG | auditory | discrimination task | 15 |
| Demopoulos 2015^31^ | children | 37 | 25 | 11.47 | 72.00 | 84.16 | no | yes | yes | 12 | 13.78 | 58.33 | 111.08 | no | no | MEG | auditory | Passive task | 13 |
| Demopoulos 2017^32^ | children | 37 | 18 | 9.82 | 100.00 | NA | no | yes | no | 19 | 9.79 | 100.00 | NA | no | yes | MEG | auditory; tactile | Passive task | 17 |
| Donkers 2015^33^ | children | 67 | 28 | 7.62 | 78.57 | NA | no | yes | no | 39 | 7.03 | 79.49 | NA | no | no | EEG | auditory | Oddball paradigm; Passive task | 17 |
| Donkers 2020^34^ | children | 67 | 28 | 7.60 | 78.57 | NA | no | no | no | 39 | 7.00 | 76.92 | NA | no | no | EEG | auditory | Oddball paradigm; Passive task | 17 |
| Dunham-Carr 2023^35^ | children | 50 | 25 | 8.90 | NA | NA | NA | NA | no | 25 | 9.10 | NA | NA | NA | NA | EEG | auditory; visual | passive with attention task between trials | 14 |
| Dunn 2008^36^ | children | 68 | 34 | 9.29 | 73.53 | 93.71 | no | NA | yes | 34 | 9.50 | 58.82 | 115.41 | no | NA | EEG | auditory | Oddball paradigm | 17 |
| Dwyer 2021^37^ | children | 211 | 130 | 3.21 | 84.60 | 65.25 | NA | yes | no | 81 | 3.09 | 64.20 | 106.37 | NA | yes | EEG | auditory | passive task | 15 |
| Edgar 2014^20^ | children | 104 | 68 | 10.26 | NA | NA | NA | NA | no | 36 | 10.87 | NA | NA | NA | NA | MEG | auditory | Passive task | 14 |
| Edgar 2015^38^ | children | 115 | 52 | 10.10 | 88.46 | 107.00 | no | yes | no | 63 | 9.80 | 92.06 | 112.60 | no | no | MEG | auditory | Passive task | 14 |
| Falter 2013^39^ | adults | 33 | 16 | 24.08 | 93.75 | 115.00 | no | no | no | 17 | 26.25 | 76.47 | 117 | no | no | MEG | visual | Discrimination task | 14 |
| Falter-Wagner 2024^40^ | adults | 33 | 15 | 26.83 | 93.30 | 113.00 | no | no | no | 18 | 26.50 | 83.33 | 116.00 | no | NA | MEG | visual | Discrimination task | 17 |
| **Study ID** | **Age group** | **n**  **_TOTAL_** | **n_autistic_** | **Age. mean (years)** | **Male. mean (%)** | **IQ. mean** | **Co-occuring condition** | **Medication** | **Language impairment** | **n_non-autistic_** | **Age. mean (years)** | **Male. mean (%)** | **IQ mean** | **Co-occuring condition** | **Medication** | **Type** | **Modality** | **Task type/neuropsychological paradigm** | **TotalQR** |
| Fan 2014^41^ | adults | 40 | 20 | 21.50 | 95.00 | 105.00 | no | NA | no | 20 | 22.00 | 95.00 | 107 | no | NA | EEG | auditory | Oddball paradigm; Passive task | 19 |
| Fanghella 2022^42^ | adults | 38 | 19 | 40.47 | 94.74 | NA | NA | NA | no | 19 | 40.84 | 94.74 | NA | NA | NA | EEG | visual; tactile | Passive task; Discrimination task; Other: emotion and gender tasks | 15 |
| Ferri 2003^43^ | adolescences | 20 | 10 | 12.30 | 100.00 | NA | yes | NA | no | 10 | 12.20 | 100.00 | NA | NA | NA | EEG | auditory | Oddball paradigm | 12 |
| Frey 2013^44^ | children | 51 | 22 | 11.30 | 95.45 | NA | NA | NA | no | 29 | 12.30 | 68.97 | NA | no | no | EEG | visual | Target detection; Other: control task | 13 |
| Fujita 2011^45^ | adults | 24 | 12 | 28.10 | 66.67 | 112.00 | NA | NA | no | 12 | 26.30 | 58.33 | NA | NA | NA | EEG | visual | Passive task; Other: memorizing task | 15 |
| Fujita 2013^46^ | adults | 19 | 9 | 30.90 | 77.78 | 105.70 | NA | Yes | no | 10 | 26.80 | 80.00 | NA | no | no | EEG | visual | Target detection | 13 |
| Gaetz 2017^47^ | children | 30 | 15 | 9.95 | 80.00 | NA | no | yes | no | 15 | 10.21 | 86.67 | NA | no | yes | MEG | tactile | Passive task | 15 |
| Gage 2003^48^ | children | 32 | 15 | 11.40 | 100.00 | NA | NA | NA | yes | 17 | 13.50 | 70.59 | NA | no | NA | MEG | auditory | Passive task | 9 |
| Gomot 2002^49^ | children | 30 | 15 | 6.83 | 80.00 | 57.00 | no | no | NA | 15 | 6.75 | 80.00 | NA | no | no | EEG | auditory | Oddball paradigm | 18 |
| Gomot 2011^50^ | children | 54 | 27 | 8.33 | 77.78 | 51.00 | no | no | no | 27 | 8.33 | 77.78 | NA | no | no | EEG | auditory | Oddball paradigm | 16 |
| Green 2020^51^ | children | 17 | 7 | 7.29 | 71.43 | NA | NA | NA | yes | 10 | 8.50 | 70.00 | NA | NA | NA | EEG | auditory | Oddball paradigm | 11 |
| **Study ID** | **Age group** | **n**  **_TOTAL_** | **n_autistic_** | **Age. mean (years)** | **Male. mean (%)** | **IQ. mean** | **Co-occuring condition** | **Medication** | **Language impairment** | **n_non-autistic_** | **Age. mean (years)** | **Male. mean (%)** | **IQ mean** | **Co-occuring condition** | **Medication** | **Type** | **Modality** | **Task type/neuropsychological paradigm** | **TotalQR** |
| Green 2023^52^ | children | 127 | 66 | 7.70 | 86.40 | NA | NA | no | no | 61 | 7.50 | 81.97 | NA | NA | no | MEG | auditory | Passive task | 14 |
| Grisoni 2019^53^ | adults | 42 | 20 | 38.00 | 55.00 | NA | NA | NA | no | 22 | 31.90 | 36.36 | NA | no | NA | EEG | auditory | Oddball paradigm; Passive task; Other: distraction-oddball paradigm | 14 |
| Gunji 2009^54^ | children | 17 | 8 | 10.80 | 87.50 | 97.00 | NA | NA | no | 9 | 11.30 | 44.44 | NA | no | NA | EEG | visual | Target detection | 13 |
| Gunji 2013^55^ | children | 18 | 9 | 7.50 | 100.00 | 83.00 | no | NA | no | 9 | 8.00 | 100.00 | NA | no | NA | EEG | visual | Target detection | 11 |
| Haigh 2022^56^ | adults | 52 | 24 | 28.50 | 79.17 | 111.71 | NA | NA | no | 28 | 28.70 | 67.86 | NA | no | no | EEG | auditory | Discrimination task; Other: Behavioral Pitch Discrimination Task | 17 |
| Hileman 2011^57^ | adolescences | 49 | 27 | 13.30 | 85.19 | >70 | no | NA | no | 22 | 14.39 | 81.82 | >70 | NA | NA | EEG | visual | Other: face processing task | 16 |
| Høyland 2019^58^ | adolescences | 98 | 49 | 15.60 | 73.47 | 91.90 | yes | yes | yes (1 participant) | 49 | 15.60 | 63.27 | NA | no | NA | EEG | visual | Other: Go-NoGo task | 18 |
| Huang 2018^59^ | children | 35 | 18 | 9.80 | 88.89 | NA | Yes | NA | yes | 17 | 9.40 | 88.24 | NA | no | NA | EEG | auditory | Oddball paradigm | 18 |
| **Study ID** | **Age group** | **n**  **_TOTAL_** | **n_autistic_** | **Age. mean (years)** | **Male. mean (%)** | **IQ. mean** | **Co-occuring condition** | **Medication** | **Language impairment** | **n_non-autistic_** | **Age. mean (years)** | **Male. mean (%)** | **IQ mean** | **Co-occuring condition** | **Medication** | **Type** | **Modality** | **Task type/neuropsychological paradigm** | **TotalQR** |
| Hudac 2018^60^ | adolescences | 133 | 102 | 12.29 | 80.39 | NA | NA | NA | no | 31 | 13.27 | 67.74 | NA | NA | NA | EEG | auditory | Oddball paradigm; Passive task | 12 |
| Isenstein 2022^61^ | adolescences | 31 | 15 | 15.08 | 73.33 | 97.02 | NA | NA | no | 16 | 13.66 | 31.25 | NA | no | NA | EEG | auditory | Passive task; Other: auditory habituation task | 12 |
| Ji 2019^62^ | children | 38 | 20 | 11.05 | 90.00 | NA | no | NA | no | 18 | 9.39 | 77.78 | NA | no | NA | EEG | visual | Discrimination task; Other: The tachistoscopic visual half-field paradigm | 15 |
| Jones 2018^63^ | children | 34 | 18 | 1.98 | NA | NA | NA | NA | no | 16 | 1.94 | NA | NA | NA | NA | EEG | visual | Passive task; Other: mother/stranger ERP paradigm | 12 |
| Kadlaskar 2021^64^ | children | 28 | 14 | 10.13 | 78.57 | NA | no | NA | no | 14 | 9.95 | 78.57 | NA | NA | NA | EEG | auditory; tactile | Oddball paradigm; Passive task | 17 |
| Kamita 2021^65^ | children | 30 | 15 | 9.07 | 86.67 | NA | no | NA | yes | 15 | NA | NA | NA | no | NA | EEG | auditory | Oddball paradigm | 10 |
| Kasai 2005^66^ | adults | 28 | 9 | 27.20 | 66.67 | 57.20 | no | yes | no | 19 | 27.30 | 68.42 | NA | no | NA | MEG | auditory | Oddball paradigm; Passive task | 16 |
| Kemner 2002^67^ | children | 23 | 12 | 10.40 | 83.33 | 96.20 | NA | no | no | 11 | 10.30 | 100.00 | 98.50 | NA | no | EEG | auditory | Passive task | 12 |
| **Study ID** | **Age group** | **n**  **_TOTAL_** | **n_autistic_** | **Age. mean (years)** | **Male. mean (%)** | **IQ. mean** | **Co-occuring condition** | **Medication** | **Language impairment** | **n_non-autistic_** | **Age. mean (years)** | **Male. mean (%)** | **IQ mean** | **Co-occuring condition** | **Medication** | **Type** | **Modality** | **Task type/neuropsychological paradigm** | **TotalQR** |
| Key 2014^68^ | children | 24 | 13 | 10.76 | 84.62 | 103.17 | no | no | no | 11 | 10.43 | 81.82 | 123.50 | no | no | EEG | visual | Target detection | 18 |
| Key 2024^69^ | adolescences | 47 | 25 | 14.53 | 80.00 | NA | NA | NA | no | 22 | 13.84 | 81.81 | NA | NA | NA | EEG | auditory | Discrimination task | 15 |
| Knight 2020^70^ | adolescences | 40 | 21 | NA | 90.48 | NA | no | yes | no | 19 | NA | 36.84 | NA | no | yes | EEG | auditory | Oddball paradigm | 15 |
| Korpilahti 2007^71^ | children | 27 | 14 | 11.20 | 100.00 | 110.00 | no | NA | no | 13 | 10.80 | 100.00 | NA | no | NA | EEG | auditory | Oddball paradigm; Passive task | 19 |
| Kovarski 2016^72^ | adults | 42 | 20 | 18.20 | 80.00 | NA | NA | NA | no | 22 | 18.40 | 77.27 | NA | NA | NA | EEG | visual | Passive task | 14 |
| Kovarski 2019^73^ | children | 36 | 18 | NA | 94.44 | NA | NA | NA | no | 18 | 7.50 | 83.33 | NA | NA | NA | EEG | visual | Passive task | 14 |
| Kröger 2014^74^ | children | 38 | 17 | 11.90 | 100.00 | 77.60 | no | NA | no | 21 | 11.63 | 100.00 | 59.10 | no | NA | EEG | visual | Discrimination task; Other: biological motion recognition | 17 |
| Kujala 2005^75^ | adults | 16 | 8 | 33.00 | 50.00 | 114.00 | no | no | no | 8 | 32 | 50.00 | NA | no | no | EEG | auditory | Target detection; Oddball paradigm; Passive task; Discrimination task; Other: stimulus-identification task | 12 |
| **Study ID** | **Age group** | **n**  **_TOTAL_** | **n_autistic_** | **Age. mean (years)** | **Male. mean (%)** | **IQ. mean** | **Co-occuring condition** | **Medication** | **Language impairment** | **n_non-autistic_** | **Age. mean (years)** | **Male. mean (%)** | **IQ mean** | **Co-occuring condition** | **Medication** | **Type** | **Modality** | **Task type/neuropsychological paradigm** | **TotalQR** |
| Kujala 2007^76^ | adults | 18 | 8 | 27.00 | 75.00 | 106.00 | no | no | no | 10 | 30 | 80.00 | 112 | no | no | EEG | auditory | Oddball paradigm; Other: multi-feature paradigm | 12 |
| Kujala 2010^77^ | children | 28 | 15 | 10.75 | 73.33 | 108.00 | no | no | no | 13 | 10.48 | 84.62 | 111 | no | NA | EEG | auditory | Other: multi-feature paradigm | 16 |
| Lacroix 2024^78^ | adults | 89 | 41 | 29.36 | 51.20 | 117.87 | yes | yes | no | 48 | 30.30 | 50.00 | 118.45 | yes | yes | EEG | visual | oddball paradigm. target detetion | 16 |
| Lambrechts 2017^79^ | adults | 36 | 18 | 25.25 | 94.44 | 112.00 | no | no | no | 18 | 26.33 | 72.22 | 116 | no | no | MEG | auditory | Discrimination task; Other: Duration and Pitch tasks | 16 |
| Lepistö 2005^80^ | children | 30 | 15 | 9.40 | 86.67 | NA | yes | no | yes (3 participants) | 15 | 9.40 | 86.67 | NA | no | NA | EEG | auditory | Oddball paradigm; Passive task | 17 |
| Lepistö 2006^81^ | children | 20 | 10 | 8.11 | 80.00 | NA | yes | no | no | 10 | 8.10 | 80.00 | NA | no | NA | EEG | auditory | Oddball paradigm; Discrimination task; Other: sound-identification task | 16 |
| **Study ID** | **Age group** | **n**  **_TOTAL_** | **n_autistic_** | **Age. mean (years)** | **Male. mean (%)** | **IQ. mean** | **Co-occuring condition** | **Medication** | **Language impairment** | **n_non-autistic_** | **Age. mean (years)** | **Male. mean (%)** | **IQ mean** | **Co-occuring condition** | **Medication** | **Type** | **Modality** | **Task type/neuropsychological paradigm** | **TotalQR** |
| Lepistö 2007^82^ | adults | 18 | 9 | 27.00 | 77.78 | NA | no | no | no | 9 | 30 | 88.89 | NA | NA | NA | EEG | auditory | Oddball paradigm | 14 |
| Lepistö 2008^83^ | children | 26 | 10 | 9.10 | 90.00 | NA | no | no | yes | 16 | 9 | 93.75 | NA | no | no | EEG | auditory | Oddball paradigm | 15 |
| Lepistö 2009^84^ | children | 30 | 16 | 8.10 | 81.25 | NA | no | no | no | 14 | 8.10 | 85.71 | NA | no | NA | EEG | auditory | Oddball paradigm; Other: Segregated and Integrated | 13 |
| Lincoln 1993^85^ | adolescences | 18 | 8 | 12.40 | NA | 71.10 | no | no | yes | 10 | 10.80 | NA | 108.60 | no | no | EEG | auditory | Oddball paradigm | 14 |
| Lincoln 1995^86^ | children | 19 | 9 | NA |  | >70 | no | no | yes | 10 | NA | NA | >70 | no | no | EEG | auditory | Passive task | 14 |
| Lindström 2016^87^ | children | 23 | 10 | 10.50 | 90.00 | NA | NA | no | yes | 13 | 10.00 | 92.31 | NA | no | NA | EEG | auditory | Oddball paradigm | 19 |
| Lindström 2018^88^ | children | 31 | 15 | 10.40 | 100.00 | NA | no | yes | no | 16 | 10.10 | 100.00 | NA | no | no | EEG | auditory | Oddball paradigm | 19 |
| Luckhardt 2017^89^ | adolescences | 37 | 21 | 12.50 | 95.24 | 103.40 | no | no | no | 16 | 13.00 | 81.25 | 106.90 | no | NA | EEG | visual | Target detection; Other: facial emotion recognition task | 15 |
| Ludlow 2014^90^ | adolescences | 22 | 11 | 13.00 | 100.00 | NA | no | NA | no | 11 | 13.70 | 100.00 | NA | no | NA | EEG | auditory | Oddball paradigm | 20 |
| Lv 2014^91^ | children | 70 | 39 | 5.79 | 94.87 | NA | no | NA | no | 31 | 6.06 | 74.19 | NA | NA | NA | EEG | auditory | Other: not mentioned | 13 |
| **Study ID** | **Age group** | **n**  **_TOTAL_** | **n_autistic_** | **Age. mean (years)** | **Male. mean (%)** | **IQ. mean** | **Co-occuring condition** | **Medication** | **Language impairment** | **n_non-autistic_** | **Age. mean (years)** | **Male. mean (%)** | **IQ mean** | **Co-occuring condition** | **Medication** | **Type** | **Modality** | **Task type/neuropsychological paradigm** | **TotalQR** |
| Maekawa 2011^92^ | adults | 18 | 9 | 28.00 | 77.78 | 107.00 | NA | NA | no | 9 | 28.90 | 44.44 | NA | NA | NA | EEG | visual | Oddball paradigm | 12 |
| Magnée 2008^93^ | adults | 25 | 12 | 21.50 | 100.00 | 122.40 | no | no | no | 13 | 23.00 | 100.00 | 127 | no | NA | EEG | visual; auditory | Discrimination task; Other: concurrent gender recognition task | 14 |
| Marco 2012^94^ | children | 14 | 7 | 9.40 | 100.00 | 84.40 | NA | no | no | 7 | 8.90 | 100.00 | 114.70 | NA | no | MEG | tactile | Oddball paradigm; Other: slow-rate stimuli paradigm and fast-rate stimuli paradigm | 13 |
| Marsicano 2024^95^ | children | 39 | 19 | 11.21 | 94.70 | 102.11 | no | no | no | 20 | 11.25 | 80.00 | NA | no | NA | EEG | visual | visual attentional task (target task) | 17 |
| Mason 2022^96^ | children | 88 | 50 | 9.75 | NA | NA | NA | NA | no | 38 | 10.04 | NA | NA | no | NA | EEG | visual | Passive task | 16 |
| Matsuzaki 2012^97^ | children | 21 | 9 | 9.64 | 100.00 | 102.17 | NA | NA | no | 12 | 10.08 | 100.00 | NA | no | NA | MEG | auditory | Other: NA | 12 |
| Matsuzaki 2019a^98^ | adults | 38 | 19 | 23.80 | 100.00 | 108.36 | no | NA | no | 19 | 26.97 | 100.00 | 113.75 | no | NA | MEG | auditory | Passive task | 15 |
| **Study ID** | **Age group** | **n**  **_TOTAL_** | **n_autistic_** | **Age. mean (years)** | **Male. mean (%)** | **IQ. mean** | **Co-occuring condition** | **Medication** | **Language impairment** | **n_non-autistic_** | **Age. mean (years)** | **Male. mean (%)** | **IQ mean** | **Co-occuring condition** | **Medication** | **Type** | **Modality** | **Task type/neuropsychological paradigm** | **TotalQR** |
| Matsuzaki 2019b^99^ | children | 48 | 21 | 10.67 | 85.71 | 83.88 | no | yes | yes | 27 | 10.14 | 92.59 | 112.96 | no | NA | MEG | auditory | Oddball paradigm | 14 |
| Matsuzaki 2019c^100^ | adults | 25 | 9 | 22.22 | 100.00 | 106.22 | no | yes | no | 16 | 27.25 | 100.00 | 114.10 | no | no | MEG | auditory | Oddball paradigm | 16 |
| McPartland 2004^101^ | adults | 23 | 9 | 21.20 | 88.89 | NA | no | NA | no | 14 | 24.60 | 92.86 | NA | no | no | EEG | visual | Target detection | 14 |
| McPartland 2011^102^ | children | 54 | 36 | 11.20 | 88.89 | 105.20 | no | no | no | 18 | 12.60 | 83.33 | 112.90 | no | no | EEG | visual | Other: face recognition | 16 |
| Megnin 2012^103^ | adolescences | 28 | 14 | 16.90 | 100.00 | NA | no | NA | no | 14 | 16.90 | 100.00 | NA | no | NA | EEG | visual; auditory | Target detection | 19 |
| Molholm 2020^104^ | children | 99 | 45 | 9.40 | 91.11 | 102.30 | NA | NA | no | 54 | 9.30 | 59.26 | 110.80 | no | NA | EEG | visual; auditory | Target detection | 13 |
| Neuhaus 2016^105^ | children | 118 | 52 | 11.30 | NA | 97.40 | no | no | no | 66 | 10.10 | NA | 116.50 | no | no | EEG | visual | Target detection | 15 |
| O'Connor 2005^106^ | adults | 30 | 15 | 24.60 | NA | NA | no | yes | no | 15 | 24.80 | NA | NA | no | NA | EEG | visual | Other: Explicit face recognition task | 17 |
| O'Connor 2007^107^ | adults | 30 | 15 | 23.50 | 100.00 | NA | no | yes | no | 15 | 23.80 | 100.00 | NA | no | NA | EEG | visual | Target detection; Discrimination task | 17 |
| **Study ID** | **Age group** | **n**  **_TOTAL_** | **n_autistic_** | **Age. mean (years)** | **Male. mean (%)** | **IQ. mean** | **Co-occuring condition** | **Medication** | **Language impairment** | **n_non-autistic_** | **Age. mean (years)** | **Male. mean (%)** | **IQ mean** | **Co-occuring condition** | **Medication** | **Type** | **Modality** | **Task type/neuropsychological paradigm** | **TotalQR** |
| OramCardy 2004^108^ | children | 18 | 10 | 11.80 | 100.00 | NA | NA | NA | yes | 8 | 12.90 | 37.50 | NA | NA | NA | MEG | auditory | Passive task | 9 |
| OramCardy 2008^109^ | adolescences | 30 | 14 | NA | 85.71 | NA | NA | NA | no | 16 | NA | 50.00 | NA | NA | NA | MEG | auditory | Passive task | 12 |
| Orekhova 2008^110^ | children | 42 | 21 | 5.92 | 80.95 | NA | no | yes | no | 21 | 5.92 | 85.71 | NA | NA | NA | EEG | auditory | Other: paired clicks sensory gating paradigm | 16 |
| Parker 2021^111^ | children | 56 | 30 | 10.50 | 80.00 | 99.79 | no | no | no | 26 | 11.21 | 57.69 | 99.63 | no | no | EEG | visual | Target detection | 15 |
| Peristeri 2023^112^ | adults | 24 | 10 | 21.80 | 20.00 | 98.40 | no | NA | no | 14 | 25.40 | 28.57 | 102.70 | no | NA | EEG | visual | picture naming task | 11 |
| Piatti 2021^113^ | children | 29 | 12 | 3.31 | 66.67 | 88.29 | NA | NA | no | 17 | 3.18 | 70.59 | NA | NA | NA | EEG | auditory | Oddball paradigm; Passive task | 16 |
| Portnova 2023^114^ | children | 50 | 25 | 4.84 | 56.00 | 87.20 | no | no | no | 25 | 5.25 | 52.00 | 90.10 | no | no | EEG | auditory | Passive task | 18 |
| Richards 2024^115^ | children | 63 | 35 | 5.45 | 97.10 | 66.00 | no | NA | no | 28 | 5.20 | 82.14 | 104.00 | NA | NA | EEG | visual | Passive task | 11 |
| Roberts 2010^116^ | children | 42 | 25 | 10.20 | NA | NA | no | yes | yes | 17 | 10.77 | NA | NA | no | NA | MEG | auditory | Passive task | 16 |
| Roberts 2011^117^ | children | 45 | 18 | 8.47 | 94.44 | 89.55 | no | NA | yes | 27 | 10065.00 | 44.44 | 109.25 | no | NA | MEG | auditory | Oddball paradigm; Passive task | 17 |
| **Study ID** | **Age group** | **n**  **_TOTAL_** | **n_autistic_** | **Age. mean (years)** | **Male. mean (%)** | **IQ. mean** | **Co-occuring condition** | **Medication** | **Language impairment** | **n_non-autistic_** | **Age. mean (years)** | **Male. mean (%)** | **IQ mean** | **Co-occuring condition** | **Medication** | **Type** | **Modality** | **Task type/neuropsychological paradigm** | **TotalQR** |
| Roberts 2019^118^ | children | 50 | 16 | 9.85 | 81.25 | NA | no | no | yes | 34 | 10.18 | 85.29 | 113.56 | no | NA | MEG | auditory | Passive task | 16 |
| Randeniya 2022^119^ | adults | 46 | 23 | 24.35 | 43.48 | NA | yes | yes | no | 23 | 24.04 | 47.83 | NA | no | no | EEG | visual; auditory | Oddball paradigm; Other: Stochastic frequency paradigm and a simultaneous.  visual 2-back tas | 19 |
| Ruiz-Martínez 2020^120^ | children | 31 | 16 | 8.96 | 93.75 | NA | NA | NA | yes | 15 | 8.86 | 93.33 | NA | no | NA | EEG | auditory | Oddball paradigm | 16 |
| Sakihara 2023^121^ | children | 17 | 12 | 11.00 | 83.30 | NA | no | no | no | 5 | 10.00 | 40.00 | NA | no | no | EEG | visual | Discrimination coherent motion | 14 |
| Schwartz 2023^122^ | adolescences | 78 | 40 | 15.94 | 72.50 | NA | NA | NA | yes | 38 | 14.75 | 55.26 | NA | no | NA | EEG | auditory | Passive task | 12 |
| Senju 2005^123^ | adolescences | 26 | 11 | 12.08 | 100.00 | NA | NA | NA | no | 15 | 12.08 | 86.67 | NA | NA | NA | EEG | visual | Target detection; Oddball paradigm | 12 |
| Shen 2017^124^ | children | 24 | 12 | 6.93 | 100.00 | NA | no | NA | yes | 12 | 7.05 | 100.00 | NA | no | NA | EEG | visual | Target detection | 15 |
| **Study ID** | **Age group** | **n**  **_TOTAL_** | **n_autistic_** | **Age. mean (years)** | **Male. mean (%)** | **IQ. mean** | **Co-occuring condition** | **Medication** | **Language impairment** | **n_non-autistic_** | **Age. mean (years)** | **Male. mean (%)** | **IQ mean** | **Co-occuring condition** | **Medication** | **Type** | **Modality** | **Task type/neuropsychological paradigm** | **TotalQR** |
| Shuffrey 2018^125^ | children | 32 | 16 | 8.04 | 68.75 | 105.00 | no | no | no | 16 | 9.44 | 56.25 | 110.56 | no | no | EEG | visual | Passive task | 15 |
| Sokhadze 2009^126^ | adolescences | 22 | 11 | 16.80 | 90.91 | 95.40 | no | NA | no | 11 | 19.40 | 81.82 | NA | no | NA | EEG | visual | Target detection; Oddball paradigm | 15 |
| Sokhadze 2012^127^ | adolescences | 32 | 16 | 12.60 | 87.50 | 95.35 | yes | yes | no | 16 | 14.60 | 81.25 | NA | no | NA | EEG | visual | Target detection; Oddball paradigm | 15 |
| Sokhadze 2016a^128^ | adolescences | 60 | 30 | 15.63 | 73.33 | >80 | no | no | no | 30 | 15.76 | 76.67 | NA | no | no | EEG | visual | Target detection; Coherent motion; Other: Cued Posner spatial attention task | 13 |
| Sokhadze 2016b^129^ | children | 32 | 18 | 11.06 | 83.33 | NA | no | NA | no | 14 | 12.60 | 85.71 | NA | no | NA | EEG | auditory | Oddball paradigm; Passive task | 9 |
| Tanaka 2023^130^ | adults | 15 | 7 | 36.00 | NA | 101.10 | NA | NA | no | 8 | 35.90 | NA | 112.60 | NA | NA | EEG | visual | Target detection | 15 |
| Tavares 2016^131^ | adults | 25 | 9 | 23.10 | 100.00 | 95.20 | NA | yes | no | 16 | 23.40 | 100.00 | 103.10 | NA | NA | EEG | visual | Discrimination task; Other: Decision task | 16 |
| Toffoli 2021^132^ | children | 86 | 29 | 11.04 | 75.86 | 110.90 | NA | NA | no | 57 | 10.50 | 56.14 | 113.53 | NA | NA | EEG | visual | Coherent motion | 14 |
| **Study ID** | **Age group** | **n**  **_TOTAL_** | **n_autistic_** | **Age. mean (years)** | **Male. mean (%)** | **IQ. mean** | **Co-occuring condition** | **Medication** | **Language impairment** | **n_non-autistic_** | **Age. mean (years)** | **Male. mean (%)** | **IQ mean** | **Co-occuring condition** | **Medication** | **Type** | **Modality** | **Task type/neuropsychological paradigm** | **TotalQR** |
| vanLaarhoven 2019^133^ | adults | 60 | 30 | 18.55 | 73.33 | 103.00 | no | no | no | 30 | 18.83 | 80.00 | 111.97 | no | no | EEG | auditory | Other: motor-auditory task | 20 |
| Vlaskamp 2017^134^ | children | 73 | 35 | 11.10 | 80.00 | 98.50 | no | yes | no | 38 | 10.90 | 71.05 | 107.60 | no | NA | EEG | auditory | Oddball paradigm; Passive task | 15 |
| Wagner 2013^135^ | adolescences | 38 | 18 | 17.00 | 100.00 | 111.20 | yes | yes | no | 20 | 17.90 | 100.00 | 116.50 | no | no | EEG | visual | Target detection | 16 |
| Wang 2017^136^ | children | 31 | 16 | 10.40 | NA | NA | NA | NA | no | 15 | 10.30 | NA | NA | NA | NA | EEG | auditory | Oddball paradigm; Passive task | 17 |
| Webb 2006^4^ | children | 45 | 27 | 3.80 | NA | NA | NA | NA | no | 18 | 3.70 | NA | NA | NA | NA | EEG | visual | Passive task | 11 |
| Weismüller 2015^137^ | children | 33 | 18 | 9.40 | 100.00 | 99.30 | no | yes | no | 15 | 10.60 | 100.00 | 118.30 | no | NA | EEG | auditory | Oddball paradigm; Passive task | 16 |
| Whitehouse 2008^138^ | children | 30 | 15 | 10.40 | 100.00 | NA | no | NA | no | 15 | 10.60 | 73.33 | NA | no | NA | EEG | auditory | Oddball paradigm; Passive task; Discrimination task | 15 |
| Yamasaki 2011^139^ | adults | 24 | 12 | NA | 75.00 | 100.30 | NA | NA | no | 12 | NA | 75.00 | NA | NA | NA | EEG | visual | Passive task; Coherent motion | 14 |
| Yamasaki 2017^140^ | adults | 28 | 14 | 29.90 | 64.29 | 104.90 | NA | NA | no | 14 | 28.60 | 64.29 | 113.10 | no | NA | EEG | visual | Passive task | 15 |
| Yoshimura 2016^141^ | children | 70 | 35 | 6.29 | 77.14 | 95.60 | NA | NA | no | 35 | 6.23 | 77.14 | 99.40 | NA | NA | MEG | auditory | Oddball paradigm; Passive task | 13 |
| **Study ID** | **Age group** | **n**  **_TOTAL_** | **n_autistic_** | **Age. mean (years)** | **Male. mean (%)** | **IQ. mean** | **Co-occuring condition** | **Medication** | **Language impairment** | **n_non-autistic_** | **Age. mean (years)** | **Male. mean (%)** | **IQ mean** | **Co-occuring condition** | **Medication** | **Type** | **Modality** | **Task type/neuropsychological paradigm** | **TotalQR** |
| Yoshimura 2021^142^ | children | 75 | 29 | 6.23 | 72.41 | 91.40 | NA | NA | no | 46 | 5.86 | 89.13 | 104.70 | NA | NA | MEG | auditory | Oddball paradigm; Passive task | 13 |
| Yu 2015a1^143^ | children | 34 | 18 | 9.30 | 88.89 | NA | no | NA | yes | 16 | 9.50 | 81.25 | NA | no | NA | EEG | auditory | Oddball paradigm; Passive task | 17 |
| Yu 2015a2^143^ | children | 34 | 16 | 9.60 | 93.75 | NA | no | NA | yes | 18 | 9.30 | 66.67 | NA | no | NA | EEG | auditory | Oddball paradigm; Passive task | 17 |
| Yu 2018^144^ | children | 31 | 15 | 9.60 | 93.33 | NA | no | no | yes | 16 | 9.80 | 81.25 | NA | no | no | EEG | auditory | Oddball paradigm; Passive task | 16 |
| Yu 2021^145^ | adolescences | 49 | 22 | 12.10 | 90.91 | NA | NA | NA | yes | 27 | 11.70 | 77.78 | NA | NA | NA | EEG | auditory | Passive task | 13 |

#### **eTable 2. Study Distribution and demographic characteristics displayed for P/M50 amplitude**

| **P/M50 amplitude** | | | | | |
| --- | --- | --- | --- | --- | --- |
|  | **k** | **Autistic n** | **Non-autistic n** | **Total n** | |
| Total | 14 | 321 | 268 | 589 | |
| EEG | 8 | 182 | 173 | 355 | |
| MEG | 6 | 130 | 95 | 225 | |
| Auditory | 9 | 238 | 159 | 397 | |
| Visual | 1 | 15 | 15 | 30 | |
| Tactile | 2 | 22 | 22 | 44 | |
| Combined modalities | 2 | 37 | 38 | 75 | |
| Children | 10 | 220 | 176 | 396 | |
| Adolescents | 1 | 15 | 15 | 30 | |
| Adults | 3 | 77 | 77 | 154 | |
|  | **Autistic** | **Non-autistic** | **t** | **df** | **p** |
| **Total** | | | | | |
| Age (years). M (SD) | 13,36 (9,17) | 14,23 (10,08) | -1.09 | 546.08 | 0.28 |
| Male (%). M (SD) | 87,23 (9,35) | 82,46 (16,17) | 4.27 | 410.41 | **<0.001** |
| IQ. M (SD) | 97,63 (17,17) | 110,67 (13,30) | -6.91 | 253.51 | **<0.001** |
| **Children** | | | | | |
| Age (years). M (SD) | 8,95 (2,83) | 8,76 (2,70) | 0.70 | 385.00 | 0.48 |
| Male (%). M (SD) | 87,34 (9,10) | 84,09 (14,21) | 2.64 | 281.17 | **0.01** |
| IQ. M (SD) | 97,35 (17,52) | 110,75 (14,43) | -5.58 | 166.44 | **<0.0001** |
| **Adolescents** | | | | | |
| Age (years). M (SD) | 13,9 (3,45) | 15,5 (4,21) | -1.14 | 26.96 | 0.26 |
| Male (%). M (SD) | 86,67 (0) | 73,33 (0) | NA | NA | NA |
| IQ. M (SD) | 92,5 (15,1) | NA | NA | NA | NA |
| **Adults** | | | | | |
| Age (years). M (SD) | 26,35 (9,89) | 26,47 (10,71) | -0.07 | 151 | 0.94 |
| Male (%). M (SD) | 87,01 (10,89) | 80,52 (20,54) | 2.45 | 116 | **0.02** |
| IQ. M (SD) | 100,80 (16,20) | 110,50 (10,60) | -2.92 | 57 | **0.005** |

#### **eTable 3. Study Distribution and demographic characteristics displayed for P/M50 latency**

| **P/M50 latency** | | | | | |
| --- | --- | --- | --- | --- | --- |
|  | **k** | **Autistic n** | **Non-autistic n** | **Total n** | |
| Total | 14 | 486 | 420 | 906 | |
| EEG | 4 | 99 | 91 | 190 | |
| MEG | 10 | 387 | 329 | 716 | |
| Auditory | 11 | 438 | 371 | 809 | |
| Visual | 1 | 15 | 15 | 30 | |
| Tactile | 1 | 15 | 15 | 30 | |
| Combined modalities | 1 | 18 | 19 | 37 | |
| Children | 11 | 428 | 362 | 790 | |
| Adolescents | 1 | 15 | 15 | 30 | |
| Adults | 2 | 43 | 43 | 86 | |
|  | **Autistic** | **Non-autistic** | **t** | **df** | **p** |
| **Total** | | | | | |
| Age (years). M (SD) | 10.17 (4.54) | 10.47 (5.39) | -0.95 | 915.90 | 0.34 |
| Male (%). M (SD) | 87.24 (6.70) | 82.38 (12.40) | 7.18 | 622.58 | 2.01 |
| IQ. M (SD) | 102.64 (20.54) | 113.05 (14.73) | -5.53 | 347.70 | 6.31 |
| **Children** | | | | | |
| Age (years). M (SD) | 9.06 (2.31) | 8.76 (2.29) | 2.01 | 897.15 | **0.04** |
| Male (%). M (SD) | 87.62 (5.33) | 83.98 (9.32) | 6.58 | 553.08 | 1.12 |
| IQ. M (SD) | 102.90 (20.73) | 112.95 (14.61) | -4.90 | 291.08 | 1.59 |
| **Adolescents** | | | | | |
| Age (years). M (SD) | 13.9 (3.45) | 15.5 (4.21) | -1.14 | 26.96 | 0.26 |
| Male (%). M (SD) | 86.67 (0) | 73.3 (0) | NA | NA | NA |
| IQ. M (SD) | 92.5 (15.1) | NA | NA | NA | NA |
| **Adults** | | | | | |
| Age (years). M (SD) | 23.53 (5.03) | 25.14 (3.20) | -1.78 | 71.22 | 0.08 |
| Male (%). M (SD) | 83.72 (14.48) | 72.09 (24.83) | 2.65 | 67.62 | **0.01** |
| IQ. M (SD) | 108.36 (19.81) | 113.75 (15.75) | -0.93 | 34.05 | 0.36 |

#### **eTable 4. Study Distribution and demographic characteristics displayed for P/M100 amplitude**

| **P/M100 amplitude** | | | | | |
| --- | --- | --- | --- | --- | --- |
|  | **k** | **Autistic n** | **Non-autistic n** | **Total n** | |
| Total | 59 | 1205 | 1118 | 2323 | |
| EEG | 54 | 1059 | 1000 | 2059 | |
| MEG | 5 | 146 | 118 | 264 | |
| Auditory | 26 | 733 | 664 | 1397 | |
| Visual | 30 | 394 | 367 | 761 | |
| Tactile | 0 | 0 | 0 | 0 | |
| Combined modalities | 3 | 78 | 87 | 165 | |
| Children | 35 | 840 | 807 | 1647 | |
| Adolescents | 11 | 224 | 169 | 393 | |
| Adults | 13 | 141 | 142 | 283 | |
|  | **Autistic** | **Non-autistic** | **t** | **df** | **p** |
| **Total** | | | | | |
| Age (years). M (SD) | 12.78 (836.96) | 13.22 (8.21) | -0.02 | 1678.35 | 0.98 |
| Male (%). M (SD) | 85.58 (9.38) | 72.00 (16.77) | 24.22 | 1812.51 | **<0.0001** |
| IQ. M (SD) | 92.87 (24.83) | 108.40 (17.27) | -14.38 | 1474.89 | **<0.000** |
| **Children** | | | | | |
| Age (years). M (SD) | 9.32 (2.49) | 9.57 (2.44) | -2.91 | 1.976.98 | **0.004** |
| Male (%). M (SD) | 86.53 (8.71) | 77.94 (15.80) | 22.04 | 1.312.47 | **<0.0001** |
| IQ. M (SD) | 88.72 (26.62) | 113.06 (14.25) | -14.92 | 1.178.49 | **<0.0001** |
| **Adolescents** | | | | | |
| Age (years). M (SD) | 14.07 (3.04) | 14.52 (3.12) | -1.87 | 614.867 | **0.06** |
| Male (%). M (SD) | 86.61 (7.21) | 78.23 (15.42) | 6.16 | 188.34 | **<0.0001** |
| IQ. M (SD) | 104.29 (15.46) | 112.99 (12.35) | -3.31 | 109.91 | **0.001** |
| **Adults** | | | | | |
| Age (years). M (SD) | 26.35 (9.70) | 26.99 (8.57) | -0.01 | 302.01 | 0.99 |
| Male (%). M (SD) | 85.11 (12.02) | 71.58 (18.53) | 8.04 | 323.63 | **0.0001** |
| IQ. M (SD) | 112.27 (14.39) | 115.92 (11.69) | -1.97 | 196.58 | 0.05 |

#### **eTable 5. Study Distribution and demographic characteristics displayed for P/M100 latency**

| **P/M100 latency** | | | | | |
| --- | --- | --- | --- | --- | --- |
|  | **k** | **Autistic n** | **Non-autistic n** | **Total n** | |
| Total | 54 | 1075 | 975 | 2050 | |
| EEG | 40 | 735 | 652 | 1387 | |
| MEG | 14 | 340 | 323 | 663 | |
| Auditory | 27 | 682 | 604 | 1286 | |
| Visual | 27 | 393 | 371 | 764 | |
| Tactile | 0 | 0 | 0 | 0 | |
| Combined modalities | 0 | 0 | 0 | 0 | |
| Children | 33 | 802 | 691 | 1493 | |
| Adolescents | 6 | 106 | 98 | 204 | |
| Adults | 15 | 167 | 186 | 353 | |
|  | **Autistic** | **Non-autistic** | **t** | **df** | **p** |
| **Total** | | | | | |
| Age (years). M (SD) | 12.35 (7.57) | 12.89 (8.15) | -1.83 | 2.810.06 | 0.07 |
| Male (%). M (SD) | 84.09 (11.56) | 77.74 (14.56) | 10.85 | 1.856.17 | 1.18 |
| IQ. M (SD) | 93.26 (25.45) | 108.19 (18.06) | -13.30 | 1.483.95 | 3.39 |
| **Children** | | | | | |
| Age (years). M (SD) | 8.28 (3.51) | 8.20 (3.34) | 0.55 | 1931.083 | 0.58 |
| Male (%). M (SD) | 84.66 (9.27) | 78.15 (13.29) | 10.81 | 1.207.07 | 4.65 |
| IQ. M (SD) | 88.61 (25.85) | 106.28 (19.03) | -13.46 | 1.160.90 | 1.79 |
| **Adolescents** | | | | | |
| Age (years). M (SD) | 14.34 (2.76) | 14.52 (3.08) | -0.63 | 367.49 | 0.53 |
| Male (%). M (SD) | 87.74 (5.48) | 80.61 (1.76) | 12.69 | 127.95 | 2.12 |
| IQ. M (SD) | 100.65 (12.47) | 106.90 (6.55) | -2.28 | 45.82 | **0.03** |
| **Adults** | | | | | |
| Age (years). M (SD) | 25.66 (7.18) | 26.77 (6.96) | -1.87 | 569.83 | 0.06 |
| Male (%). M (SD) | 79.04 (19.83) | 74.73 (20.96) | 1.98 | 350.03 | 0.05 |
| IQ. M (SD) | 111.60 (14.92) | 115.43 (12.73) | -2.36 | 288.33 | **0.02** |

#### **eTable 6. Study Distribution and demographic characteristics displayed for P/M200 amplitude**

| **P/M200 amplitude** | | | | | |
| --- | --- | --- | --- | --- | --- |
|  | **k** | **Autistic n** | **Non-autistic n** | **Total n** | |
| Total | 22 | 528 | 606 | 1134 | |
| EEG | 20 | 492 | 569 | 1061 | |
| MEG | 2 | 36 | 37 | 73 | |
| Auditory | 10 | 307 | 381 | 688 | |
| Visual | 8 | 119 | 112 | 231 | |
| Tactile | 0 | 0 | 0 | 0 | |
| Combined modalities | 4 | 102 | 113 | 215 | |
| Children | 9 | 258 | 311 | 569 | |
| Adolescents | 6 | 155 | 179 | 334 | |
| Adults | 7 | 115 | 116 | 231 | |
|  | **Autistic** | **Non-autistic** | **t** | **df** | **p** |
| **Total** | | | | | |
| Age (years). M (SD) | 13.98 (7.59) | 13.75 (7.52) | 0.51 | 1107.87 | 0.61 |
| Male (%). M (SD) | 85.18 (10.08) | 85.18 (10.08) | 22.42 | 886.70 | **<0.0001** |
| IQ. M (SD) | 100.60 (18.35) | 100.60 (18.35) | -7.58 | 452.00 | **<0.0001** |
| **Children** | | | | | |
| Age (years). M (SD) | 9.18 (3.17) | 9.61 (3.22) | -1.58 | 550.56 | 0.12 |
| Male (%). M (SD) | 89.29 (7.31) | 89.29 (7.31) | 19.19 | 356.82 | **<0.0001** |
| IQ. M (SD) | 101.4 (17.91) | 101.4 (17.91) | -5.15 | 226.57 | **<0.0001** |
| **Adolescents** | | | | | |
| Age (years). M (SD) | 15.40 (3.06) | 14.48 (3.50) | 1.56 | 331.97 | 0.12 |
| Male (%). M (SD) | 80.65 (6.44) | 61.45 (13.39) | 17.04 | 264.06 | **<0.0001** |
| IQ. M (SD) | 94.93 (20.82) | NA | NA | NA | NA |
| **Adults** | | | | | |
| Age (years). M (SD) | 23.31 (8.76) | 23.71 (9.47) | -0,34 | 227,92 | 0,73 |
| Male (%). M (SD) | 83 (15.12) | 74.26 (16.24) | 3,95 | 198,25 | **0,00** |
| IQ. M (SD) | 101.94 (17.33) | 112.40 (10.12) | -4,48 | 135,88 | **<0,0001** |

#### **eTable 7. Study Distribution and demographic characteristics displayed for P/M200 latency**

| **P/M200 latency** | | | | | |
| --- | --- | --- | --- | --- | --- |
|  | **k** | **Autistic n** | **Non-autistic n** | **Total n** | |
| Total | 14 | 264 | 270 | 534 | |
| EEG | 13 | 246 | 251 | 497 | |
| MEG | 1 | 18 | 19 | 37 | |
| Auditory | 6 | 153 | 165 | 318 | |
| Visual | 7 | 93 | 86 | 179 | |
| Tactile | 0 | 0 | 0 | 0 | |
| Combined modalities | 1 | 18 | 19 | 37 | |
| Children | 6 | 92 | 72 | 164 | |
| Adolescents | 4 | 104 | 130 | 234 | |
| Adults | 4 | 68 | 68 | 136 | |
|  | **Autistic** | **Non-autistic** | **t** | **df** | **p** |
| **Total** | | | | | |
| Age (years). M (SD) | 13.61 (6.46) | 13.08 (5.82) | 1.06 | 577.90 | 0.29 |
| Male (%). M (SD) | 87.88 (7.91) | 74.81 (19.90) | 10.01 | 353.52 | 6.19 |
| IQ. M (SD) | 97.89 (19.12) | NA | NA | NA | NA |
| **Children** | | | | | |
| Age (years). M (SD) | 7.44 (3.06) | 6.14 (2.97) | 3.02 | 195.98 | **0.00** |
| Male (%). M (SD) | 94.57 (5.76) | 95.83 (6.28) | -1.33 | 145.90 | 0.19 |
| IQ. M (SD) | NA | NA | NA | NA | NA |
| **Adolescents** | | | | | |
| Age (years). M (SD) | 14.50 (2.68) | 13.99 (2.88) | 1.42 | 226.71 | 0.16 |
| Male (%). M (SD) | 82.69 (4.58) | 61.54 (14.16) | 16.02 | 161.47 | 3.42 |
| IQ. M (SD) | 94.76 (22.30) | NA | NA | NA | NA |
| **Adults** | | | | | |
| Age (years). M (SD) | 20.02 (5.89) | 19.77 (5.11) | 0.29 | 160.83 | 0.77 |
| Male (%). M (SD) | 86.76 (7.94) | 77.94 (18.23) | 3.66 | 91.53 | **0.00** |
| IQ. M (SD) | 99.28 (17.35) | NA | NA | NA | NA |

#### **eTable 8. Study Distribution and demographic characteristics displayed for N100 amplitude**

| **N100 amplitude** | | | | | | | |
| --- | --- | --- | --- | --- | --- | --- | --- |
|  | **k** | | **Autistic n** | **Non-autistic n** | | **Total n** | |
| Total | 26 | | 629 | 693 | | 1322 | |
| EEG | 26 | | 629 | 693 | | 1322 | |
| MEG | 0 | | 0 | 0 | | 0 | |
| Auditory | 13 | | 376 | 424 | | 800 | |
| Visual | 11 | | 194 | 200 | | 394 | |
| Tactile | 0 | | 0 | 0 | | 0 | |
| Combined modalities | 2 | | 59 | 69 | | 128 | |
| Children | 7 | | 213 | 271 | | 484 | |
| Adolescents | 10 | | 260 | 284 | | 544 | |
| Adults | 9 | | 156 | 138 | | 294 | |
|  | | **Autistic** | **Non-autistic** | | **t** | **df** | **p** |
| **Total** | | | | | | | |
| Age (years). M (SD) | | 15.37 (6.48) | 14.99 (6.82) | | 1.02 | 1.287 | 0.31 |
| Male (%). M (SD) | | 80.24 (12.97) | 63.73 (16.58) | | 19.58 | 1.216 | **<0.0001** |
| IQ. M (SD) | | 100.34 (17.61) | 110.07 (11.16) | | -8.93 | 685 | **<0.0001** |
| **Children** | | | | | | | |
| Age (years). M (SD) | | 9.78 (2.75) | 10.00 (2.97) | | -0.81 | 469.20 | 0.42 |
| Male (%). M (SD) | | 87.77 (6.76) | 57.38 (14.86) | | 27.92 | 348.00 | **<0.0001** |
| IQ. M (SD) | | 102.15 (17.09) | 110.01 (11.17) | | -5.26 | 301.66 | **<0.0001** |
| **Adolescents** | | | | | | | |
| Age (years). M (SD) | | 14.96 (3.22) | 14.73 (3.43) | | 0.78 | 541.66 | 0.44 |
| Male (%). M (SD) | | 79.62 (7.54) | 65.84 (13.90) | | 14.53 | 444.10 | **<0.0001** |
| IQ. M (SD) | | 93.59 (19.24) | NA | | NA | NA | NA |
| **Adults** | | | | | | | |
| Age (years). M (SD) | | 25.19 (7.89) | 25.32 (8.91) | | -0.13 | 266.41 | 0.90 |
| Male (%). M (SD) | | 71.21 (19.89) | 70.29 (20.37) | | 0.38 | 267.89 | 0.71 |
| IQ. M (SD) | | 103.63 (15.19) | 110.19 (11.11) | | -3.61 | 207.23 | **0.00** |

#### **eTable 9. Study Distribution and demographic characteristics displayed for N100 latency**

| **N100 latency** | | | | | | | |
| --- | --- | --- | --- | --- | --- | --- | --- |
|  | **k** | | **Autistic n** | **Non-autistic n** | | **Total n** | |
| Total | 19 | | 369 | 402 | | 771 | |
| EEG | 18 | | 347 | 371 | | 718 | |
| MEG | 1 | | 22 | 31 | | 53 | |
| Auditory | 10 | | 201 | 232 | | 433 | |
| Visual | 9 | | 168 | 170 | | 338 | |
| Tactile | 0 | | 0 | 0 | | 0 | |
| Combined modalities | 0 | | 0 | 0 | | 0 | |
| Children | 5 | | 75 | 72 | | 147 | |
| Adolescents | 9 | | 226 | 261 | | 487 | |
| Adults | 5 | | 68 | 69 | | 137 | |
|  | | **Autistic** | **Non-autistic** | | **t** | **df** | **p** |
| **Total** | | | | | | | |
| Age (years). M (SD) | | 14.68 (4.9) | 13.80 (4.97) | | 2.51 | 799.22 | **0.01** |
| Male (%). M (SD) | | 81.43 (9.78) | 70.56 (16.67) | | 11.10 | 640.01 | **<0.0000** |
| IQ. M (SD) | | 98.36 (17.59) | 110.96 (15.25) | | -4.12 | 46.35 | **0.00** |
| **Children** | | | | | | | |
| Age (years). M (SD) | | 6.56 (2.82) | 4.40 (3.87) | | 3.85 | 130 | **0.00** |
| Male (%). M (SD) | | 90.90 (6.44) | 87.23 (8.25) | | 2.54 | 83 | **0.01** |
| IQ. M (SD) | | 110.00 (0.00) | NA | | NA | NA | NA |
| **Adolescents** | | | | | | | |
| Age (years). M (SD) | | 13.70 (3.36) | 13.14 (3.55) | | 1.82 | 515.88 | 0.07 |
| Male (%). M (SD) | | 81.89 (8.53) | 70.86 (16.27) | | 9.86 | 429.81 | **<0.0000** |
| IQ. M (SD) | | 93.11 (18.38) | 109.20 (18.55) | | -3.33 | 21.30 | **0.00** |
| **Adults** | | | | | | | |
| Age (years). M (SD) | | 27.14 (4.70) | 26.27 (5.29) | | 1.01 | 133.61 | 0.31 |
| Male (%). M (SD) | | 70.59 (4.96) | 57.97 (11.15) | | 8.58 | 94.17 | **<0.0000** |
| IQ. M (SD) | | 107.43 (13.77) | 113.10 (9.40) | | -1.74 | 32.25 | 0.09 |

#### **eTable 10. Study Distribution and demographic characteristics displayed for N170 amplitude**

| **N170 amplitude** | | | | | |
| --- | --- | --- | --- | --- | --- |
|  | **k** | **Autistic n** | **Non-autistic n** | **Total n** | |
| Total | 28 | 732 | 670 | 1402 | |
| EEG | 28 | 732 | 670 | 1402 | |
| MEG | 0 | 0 | 0 | 0 | |
| Auditory | 0 | 0 | 0 | 0 | |
| Visual | 27 | 718 | 656 | 1374 | |
| Tactile | 0 | 0 | 0 | 0 | |
| Combined modalities | 1 | 14 | 14 | 28 | |
| Children | 11 | 299 | 260 | 559 | |
| Adolescents | 6 | 196 | 171 | 367 | |
| Adults | 11 | 237 | 239 | 476 | |
|  | **Autistic** | **Non-autistic** | **t** | **df** | **p** |
| **Total** | | | | | |
| Age (years). M (SD) | 15.50 (8.25) | 16.33 (8.62) | -1.82 | 1375.69 | 0.07 |
| Male (%). M (SD) | 86.48 (15.24) | 79.78 (18.11) | 5.39 | 693.76 | **<0.0001** |
| IQ. M (SD) | 100.22 (21.53) | 112.60 (14.38) | -8.27 | 535.44 | **<0.0001** |
| **Children** | | | | | |
| Age (years). M (SD) | 9.37 (3.81) | 9.33 (3.37) | 0.11 | 556.85 | 0.91 |
| Male (%). M (SD) | 90.53 (6.84) | 78.10 (16) | 8.49 | 176.16 | **<0.0001** |
| IQ. M (SD) | 93.52 (22.89) | 111.29 (15.19) | -8.32 | 303.02 | **<0.0001** |
| **Adolescents** | | | | | |
| Age (years). M (SD) | 14.68 (2.33) | 15.11 (2.29) | -1.78 | 359.80 | 0.08 |
| Male (%). M (SD) | 94.51 (6.34) | 89.66 (8.48) | 4.31 | 159.05 | **<0.0001** |
| IQ. M (SD) | 107.00 (14.46) | 112.23 (12.79) | -1.66 | 72.88 | 0.10 |
| **Adults** | | | | | |
| Age (years). M (SD) | 23.92 (7.20) | 24.80 (7.03) | -1.34 | 473.53 | 0.18 |
| Male (%). M (SD) | 73.63 (20.80) | 75 (21.95) | -0.50 | 236.01 | 0.62 |
| IQ. M (SD) | 109.91 (16.11) | 114.70 (13.39) | -2.24 | 179.32 | **0.03** |

#### **eTable 11. Study Distribution and demographic characteristics displayed for N170 latency**

| **N170 latency** | | | | | | | |
| --- | --- | --- | --- | --- | --- | --- | --- |
|  | **k** | | **Autistic n** | **Non-autistic n** | | **Total n** | |
| Total | 25 | | 344 | 327 | | 671 | |
| EEG | 25 | | 344 | 327 | | 671 | |
| MEG | 0 | | 0 | 0 | | 0 | |
| Auditory | 0 | | 0 | 0 | | 0 | |
| Visual | 24 | | 330 | 313 | | 643 | |
| Tactile | 0 | | 0 | 0 | | 0 | |
| Combined modalities | 1 | | 14 | 14 | | 28 | |
| Children | 11 | | 171 | 141 | | 312 | |
| Adolescents | 5 | | 73 | 67 | | 140 | |
| Adults | 9 | | 100 | 119 | | 219 | |
|  | | **Autistic** | **Non-autistic** | | **t** | **df** | **p** |
| **Total** | | | | | | | |
| Age (years). M (SD) | | 15.23 (8.18) | 15.97 (8.60) | | -1.61 | 1316.813 | 0.11 |
| Male (%). M (SD) | | 85.46 (15.29) | 76.76 (18.02) | | 6.73 | 640.12 | 3.76 |
| IQ. M (SD) | | 98.76 (21.84) | 111.47 (14.18) | | -7.83 | 457.44 | 3.52 |
| **Children** | | | | | | | |
| Age (years). M (SD) | | 9.37 (3.85) | 9.35 (3.41) | | 0.04 | 562.94 | 0.97 |
| Male (%). M (SD) | | 90.64 (6.70) | 75.89 (16.66) | | 9.88 | 177.22 | 1.31 |
| IQ. M (SD) | | 92.74 (23.30) | 110.32 (15.12) | | -7.83 | 277.60 | 1.01 |
| **Adolescents** | | | | | | | |
| Age (years). M (SD) | | 14.54 (2.23) | 14.75 (2.04) | | -1.25 | 325.13 | 0.21 |
| Male (%). M (SD) | | 93.15 (6.39) | 86.57 (7.20) | | 5.70 | 132.42 | 7.34 |
| IQ. M (SD) | | 103.40 (8.90) | 106.9 (6.55) | | -1.38 | 34.98 | 0.18 |
| **Adults** | | | | | | | |
| Age (years). M (SD) | | 23.63 (7.15) | 24.52 (7.10) | | -1.34 | 451.00 | 0.18 |
| Male (%). M (SD) | | 70.99 (19.99) | 72.27 (21.41) | | -0.46 | 214.58 | 0.65 |
| IQ. M (SD) | | 109.27 (16.21) | 114.18 (13.13) | | -2.16 | 157.70 | **0.03** |

#### **eTable 12. Study Distribution and demographic characteristics displayed for N200 amplitude**

| **N200 amplitude** | | | | | | | |
| --- | --- | --- | --- | --- | --- | --- | --- |
|  | **k** | | **Autistic n** | **Non-autistic n** | | **Total n** | |
| Total | 23 | | 422 | 460 | | 882 | |
| EEG | 23 | | 422 | 460 | | 882 | |
| MEG | 0 | | 0 | 0 | | 0 | |
| Auditory | 14 | | 291 | 299 | | 590 | |
| Visual | 7 | | 105 | 134 | | 239 | |
| Tactile | 0 | | 0 | 0 | | 0 | |
| Combined modalities | 2 | | 26 | 27 | | 53 | |
| Children | 15 | | 287 | 316 | | 603 | |
| Adolescents | 6 | | 110 | 118 | | 228 | |
| Adults | 2 | | 25 | 26 | | 51 | |
|  | | **Autistic** | **Non-autistic** | | **t** | **df** | **p** |
| **Total** | | | | | | | |
| Age (years). M (SD) | | 10.92 (5.33) | 10.81 (4.95) | | 0.32 | 858.18 | 0.75 |
| Male (%). M (SD) | | 81.75 (11.66) | 72.17 (16.23) | | 10.13 | 833.17 | **<0.0001** |
| IQ. M (SD) | | 91.25 (26.76) | 102.48 (26.83) | | -3.86 | 314.28 | **0.00** |
| **Children** | | | | | | | |
| Age (years). M (SD) | | 8.43 (3,17) | 8.60 (2.82) | | -0.69 | 574.84 | 0.49 |
| Male (%). M (SD) | | 80.14 (11.50) | 73.10 (13.57) | | 6.89 | 598.17 | **<0.0001** |
| IQ. M (SD) | | 88.07 (27.09) | 100,10 (26.56) | | -3.77 | 279.56 | **0.00** |
| **Adolescents** | | | | | | | |
| Age (years). M (SD) | | 14.46 (2.97) | 13.90 (3.74) | | 1.26 | 220.45 | 0.21 |
| Male (%). M (SD) | | 83.64 (11.36) | 67.80 (20.37) | | 7.32 | 185.88 | **<0.0001** |
| IQ. M (SD) | | 96.33 (23.39) | NA | | NA | NA | NA |
| **Adults** | | | | | | | |
| Age (years). M (SD) | | 23.93 (5.11) | 23.63 (2.57) | | 0.27 | 35.08 | 0.79 |
| Male (%). M (SD) | | 92 (7.69) | 80.77 (19.23) | | 2.76 | 33.05 | **0.01** |
| IQ. M (SD) | | 105.03 (22.51) | 127.00 (14.4) | | -3.65 | 34.24 | **0.00** |

#### **eTable 13. Study Distribution and demographic characteristics displayed for N200 latency**

| **N200 latency** | | | | | | | |
| --- | --- | --- | --- | --- | --- | --- | --- |
|  | **k** | | **Autistic n** | **Non-autistic n** | | **Total n** | |
| Total | 18 | | 327 | 366 | | 693 | |
| EEG | 18 | | 327 | 366 | | 693 | |
| MEG | 0 | | 0 | 0 | | 0 | |
| Auditory | 11 | | 222 | 232 | | 454 | |
| Visual | 7 | | 105 | 134 | | 239 | |
| Tactile | 0 | | 0 | 0 | | 0 | |
| Combined modalities | 0 | | 0 | 0 | | 0 | |
| Children | 13 | | 255 | 284 | | 539 | |
| Adolescents | 4 | | 59 | 69 | | 128 | |
| Adults | 1 | | 13 | 13 | | 26 | |
|  | | **Autistic** | **Non-autistic** | | **t** | **df** | **p** |
| **Total** | | | | | | | |
| Age (years). M (SD) | | 9.96 (5.14) | 9.63 (4.71) | | 0,92 | 659,19 | 0,36 |
| Male (%). M (SD) | | 81.65 (11.04) | 71.13 (16.15) | | 9,35 | 622,16 | **<0,0000** |
| IQ. M (SD) | | 88.27 (26.89) | 98.34 (27.81) | | -3,05 | 241,44 | **0,003** |
| **Children** | | | | | | | |
| Age (years). M (SD) | | 8.26 (3.28) | 8.01 (2.93) | | 0,93 | 512,36 | 0,35 |
| Male (%). M (SD) | | 80 (11.11) | 72.86 (14.65) | | 6,30 | 498,37 | **<0,0000** |
| IQ. M (SD) | | 86.61 (27.88) | 98.34 (27.81) | | -3,31 | 242,10 | **0,001** |
| **Adolescents** | | | | | | | |
| Age (years). M (SD) | | 13.73 (3.06) | 13.46 (4.23) | | 0,41 | 122,77 | 0,68 |
| Male (%). M (SD) | | 88.14 (9.29) | 69.57 (21.31) | | 6,55 | 96,00 | **<0,0000** |
| IQ. M (SD) | | 96.33 (23.39) | NA | | NA | NA | NA |
| **Adults** | | | | | | | |
| Age (years). M (SD) | | 26.17 (5) | 24.25 (2) | | 1,29 | 15,74 | 0,22 |
| Male (%). M (SD) | | 84.62 (0) | 61.54 (0) | | NA | NA | NA |
| IQ. M (SD) | | 89 (19) | NA | | NA | NA | NA |

#### **eTable 14. Study Distribution and demographic characteristics displayed for MMN/MMF amplitude**

| **MMN/MMF amplitude** | | | | | | | |
| --- | --- | --- | --- | --- | --- | --- | --- |
|  | **k** | | **Autistic n** | **Non-autistic n** | | **Total n** | |
| Total | 39 | | 871 | 744 | | 1615 | |
| EEG | 36 | | 754 | 674 | | 1428 | |
| MEG | 3 | | 117 | 70 | | 187 | |
| Auditory | 36 | | 798 | 664 | | 1462 | |
| Visual | 2 | | 50 | 57 | | 107 | |
| Tactile | 0 | | 0 | 0 | | 0 | |
| Combined modalities | 1 | | 23 | 23 | | 46 | |
| Children | 23 | | 506 | 432 | | 938 | |
| Adolescents | 5 | | 157 | 84 | | 241 | |
| Adults | 11 | | 208 | 228 | | 436 | |
|  | | **Autistic** | **Non-autistic** | | **t** | **df** | **p** |
| **Total** | | | | | | | |
| Age (years). M (SD) | | 13.77 (8.13) | 15.02 (8.49) | | -3.01 | 1548.09 | **0.00** |
| Male (%). M (SD) | | 82.46 (13.87) | 73.11 (19.68) | | 10.74 | 1279.65 | **<0.0001** |
| IQ. M (SD) | | 95.74 (25.23) | 113.33 (13.42) | | -12.66 | 752.83 | **<0.0001** |
| **Children** | | | | | | | |
| Age (years). M (SD) | | 9.32 (2.49) | 9.57 (2.44) | | -1.56 | 918.51 | 0.12 |
| Male (%). M (SD) | | 86.53 (8.71) | 77.94 (15.80) | | 9.90 | 623.44 | **<0.0001** |
| IQ. M (SD) | | 88.72 (26.62) | 113.06 (14.25) | | -12.91 | 467.86 | **<0.0001** |
| **Adolescents** | | | | | | | |
| Age (years). M (SD) | | 12.40 (3.48) | 13.03 (2.63) | | 1.38 | 174.51 | 0.17 |
| Male (%). M (SD) | | 84.71 (6.92) | 65.48 (23.30) | | 7.39 | 90.90 | **<0.0001** |
| IQ. M (SD) | | 103.69 (14.95) | 115.08 (10.76) | | -2.23 | 21.80 | **0.04** |
| **Adults** | | | | | | | |
| Age (years). M (SD) | | 26.87 (8.17) | 27.16 (7.38) | | -0.38 | 418.54 | 0.70 |
| Male (%). M (SD) | | 71.15 (20.14) | 67.11 (21.95) | | 2.01 | 433.98 | **0.05** |
| IQ. M (SD) | | 108.76 (16.07) | 113.51 (12.51) | | -2.80 | 282.02 | **0.01** |

#### **eTable 15. Study Distribution and demographic characteristics displayed for MMN/MMF latency**

| **MMN/MMF latency** | | | | | | | |
| --- | --- | --- | --- | --- | --- | --- | --- |
|  | **k** | | **Autistic n** | **Non-autistic n** | | **Total n** | |
| Total | 31 | | 693 | 580 | | 1273 | |
| EEG | 26 | | 560 | 482 | | 1042 | |
| MEG | 5 | | 133 | 98 | | 231 | |
| Auditory | 29 | | 643 | 523 | | 1166 | |
| Visual | 2 | | 50 | 57 | | 107 | |
| Tactile | 0 | | 0 | 0 | | 0 | |
| Combined modalities | 0 | | 0 | 0 | | 0 | |
| Children | 21 | | 439 | 373 | | 812 | |
| Adolescents | 2 | | 112 | 41 | | 153 | |
| Adults | 8 | | 142 | 166 | | 308 | |
|  | | **Autistic** | **Non-autistic** | | **t** | **df** | **p** |
| **Total** | | | | | | | |
| Age (years). M (SD) | | 12.97 (6.97) | 14.53 | | -3.70 | 1.161.77 | **0.00** |
| Male (%). M (SD) | | 84.05 (11.61) | 76.46 (16.80) | | 9.07 | 974.36 | **<0.0000** |
| IQ. M (SD) | | 92.98 (27.45) | 111.90 (13.19) | | -11.12 | 547.81 | **<0.0000** |
| **Children** | | | | | | | |
| Age (years). M (SD) | | 9.21 (2.53) | 9.38 (2.30) | | -0.99 | 806.23 | 0.32 |
| Male (%). M (SD) | | 86.76 (8.02) | 77.37 (15.14) | | 10.54 | 521.57 | **<0.0000** |
| IQ. M (SD) | | 86.18 (28.38) | 109.66 (13.77) | | -9.87 | 305.75 | **<0.0000** |
| **Adolescents** | | | | | | | |
| Age (years). M (SD) | | 12.29 (3.71) | 13.01 (2.85) | | -1.27 | 92.02 | 0.21 |
| Male (%). M (SD) | | 82.14 (5.59) | 75.61 (13.85) | | 2.93 | 44.86 | **0.01** |
| IQ. M (SD) | | NA | NA | | NA | NA | NA |
| **Adults** | | | | | | | |
| Age (years). M (SD) | | 25.12 (6.97) | 26.49 (6.51) | | -1.77 | 291.34 | 0.08 |
| Male (%). M (SD) | | 77.47 (18.94) | 74.33 (20.33) | | 1.23 | 303.76 | 0.22 |
| IQ. M (SD) | | 104.43 (20.92) | 113.51 (12.51) | | -4.28 | 216.89 | <**0.0000** |

#### **Forest plots**

##### **eFigure 1. Forest Plot from amplitude differences in P/M50 component between groups.**

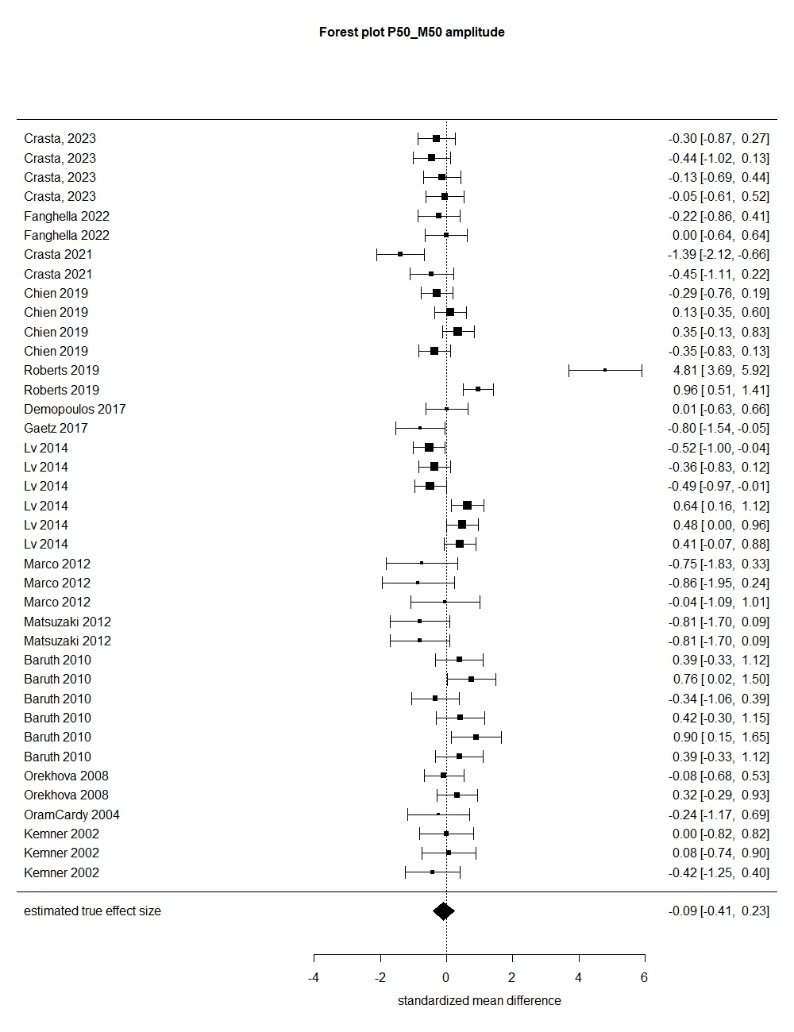

##### **eFigure 2. Forest Plot from amplitude differences in P/M100 component between groups.**

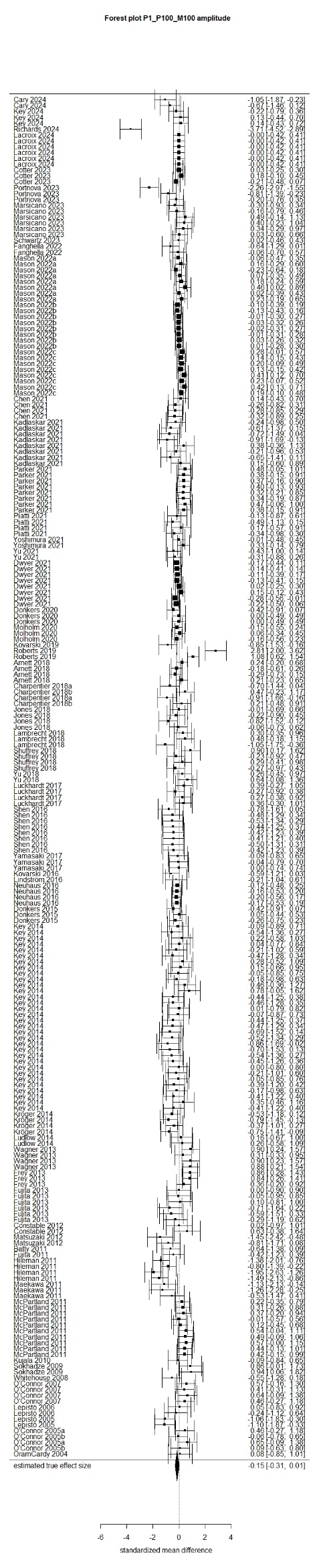

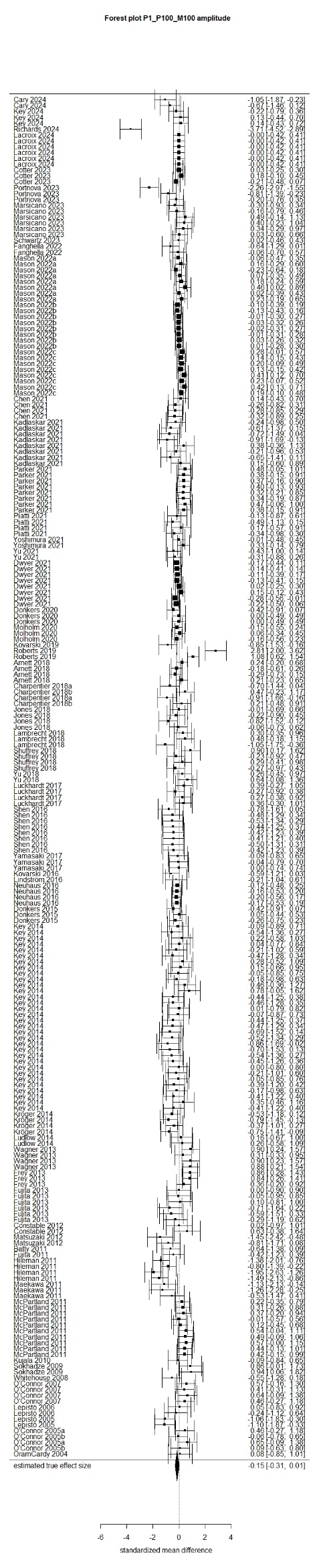

##### **eFigure 3. Forest Plot from amplitude differences in P/M200 component between groups.**

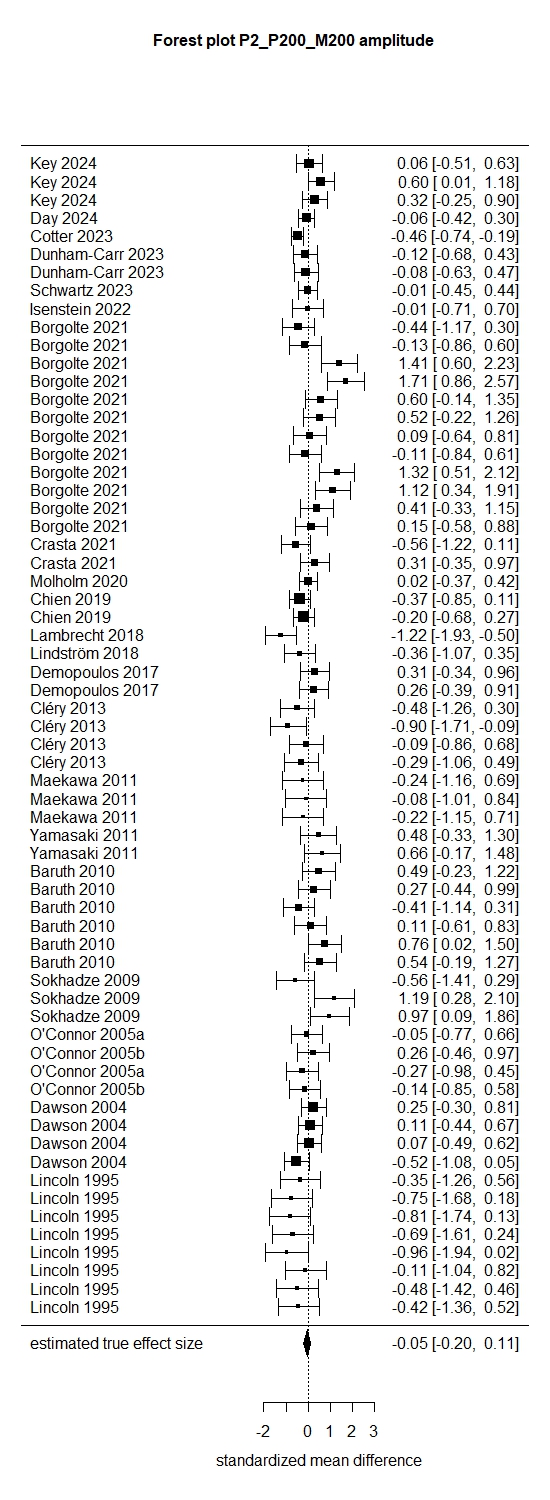

##### **eFigure 4. Forest Plot from amplitude differences in N100 component between groups.**

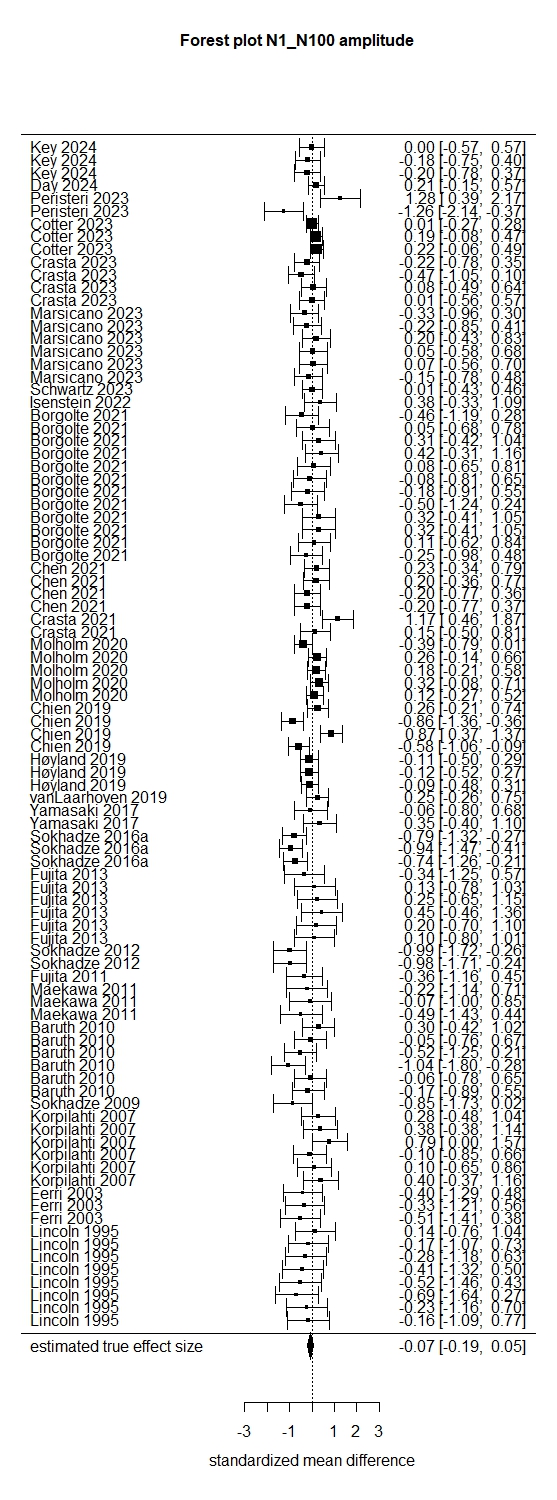

##### **eFigure 5. Forest Plot from latency differences in N100 component between groups.**

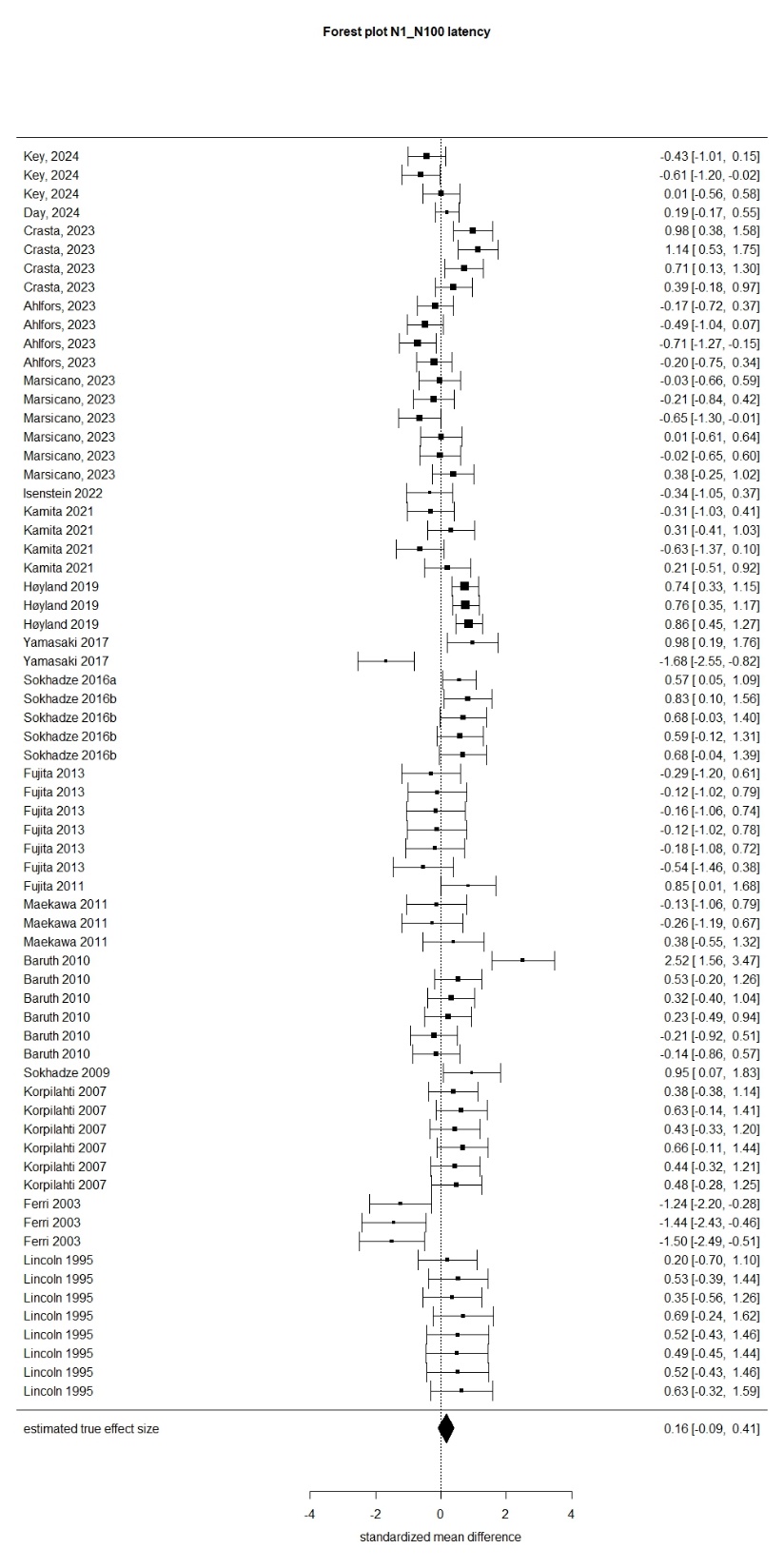

##### **eFigure 6. Forest Plot from amplitude differences in N170 component between groups.**

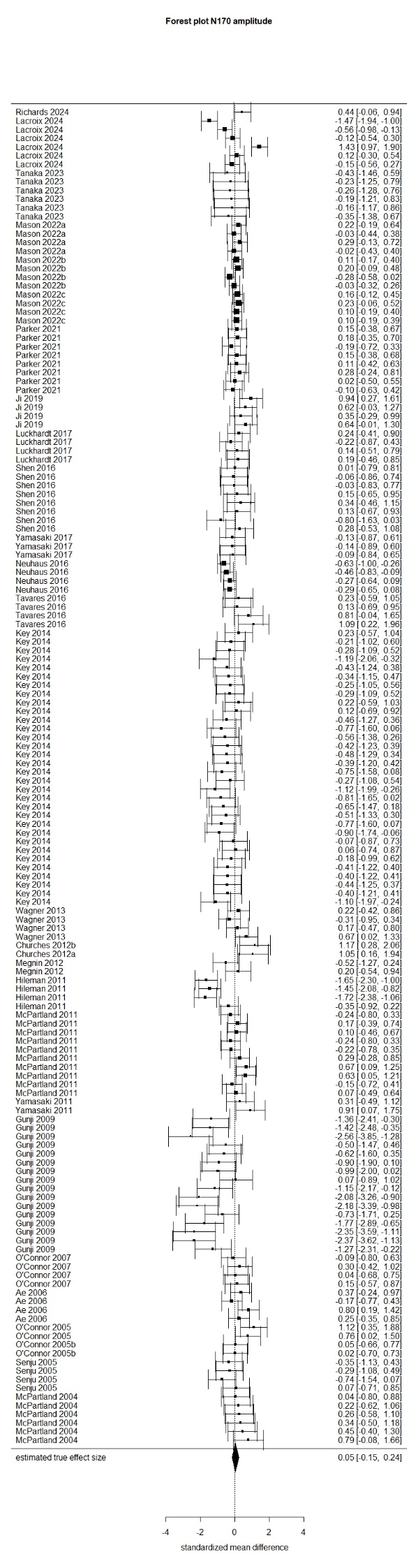

##### **eFigure 7. Forest Plot from amplitude differences in N200 component between groups.**

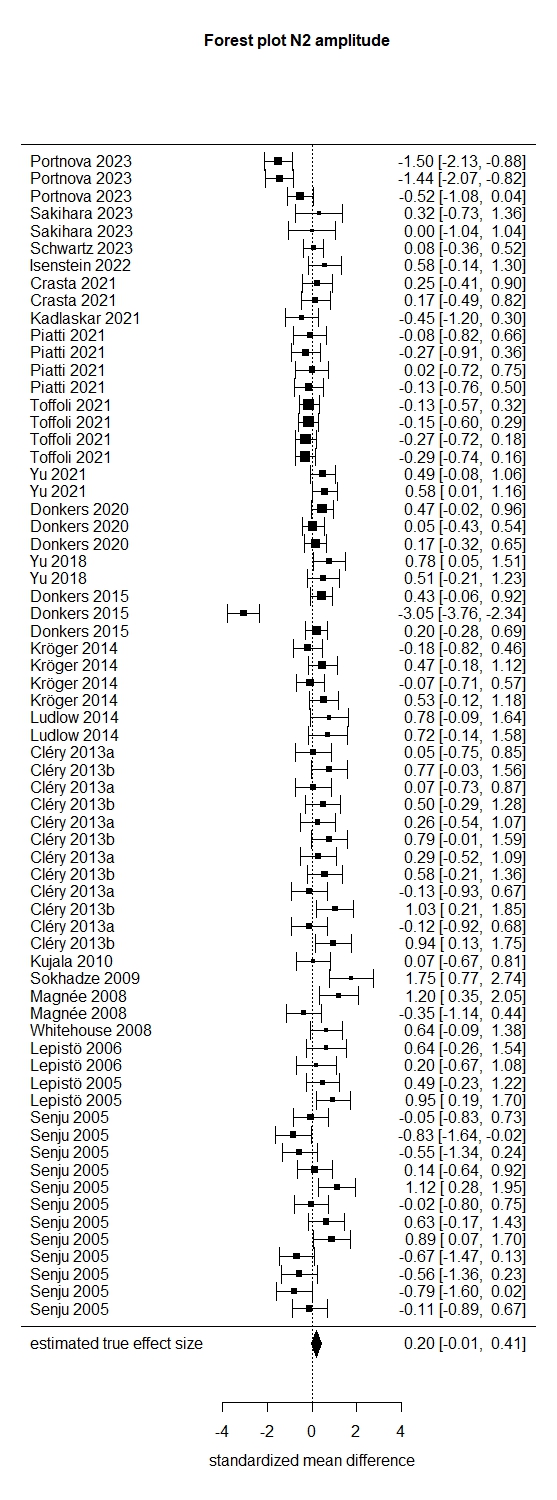

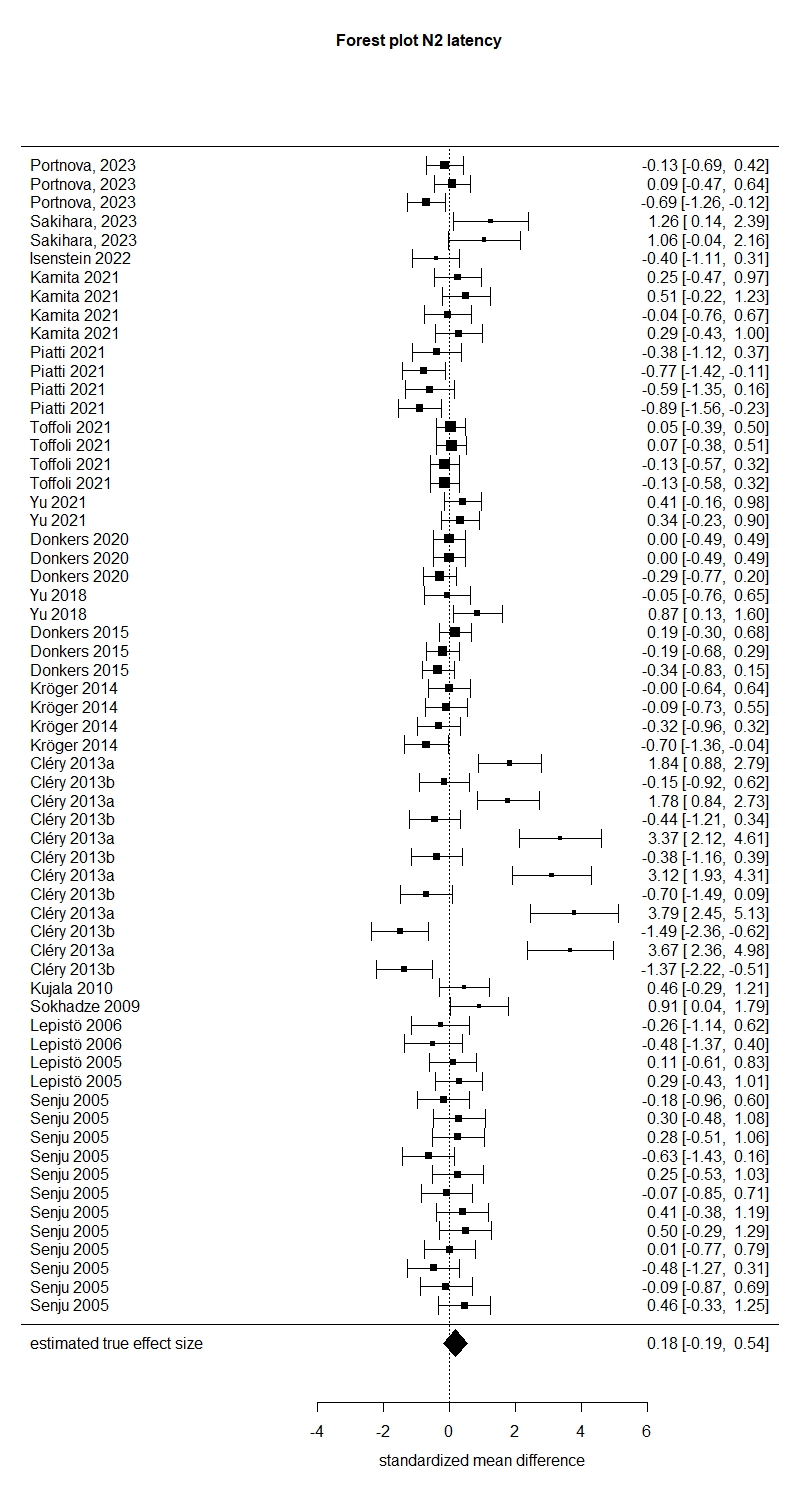

##### **eFigure 8. Forest Plot from latency differences in N200 component between groups.**

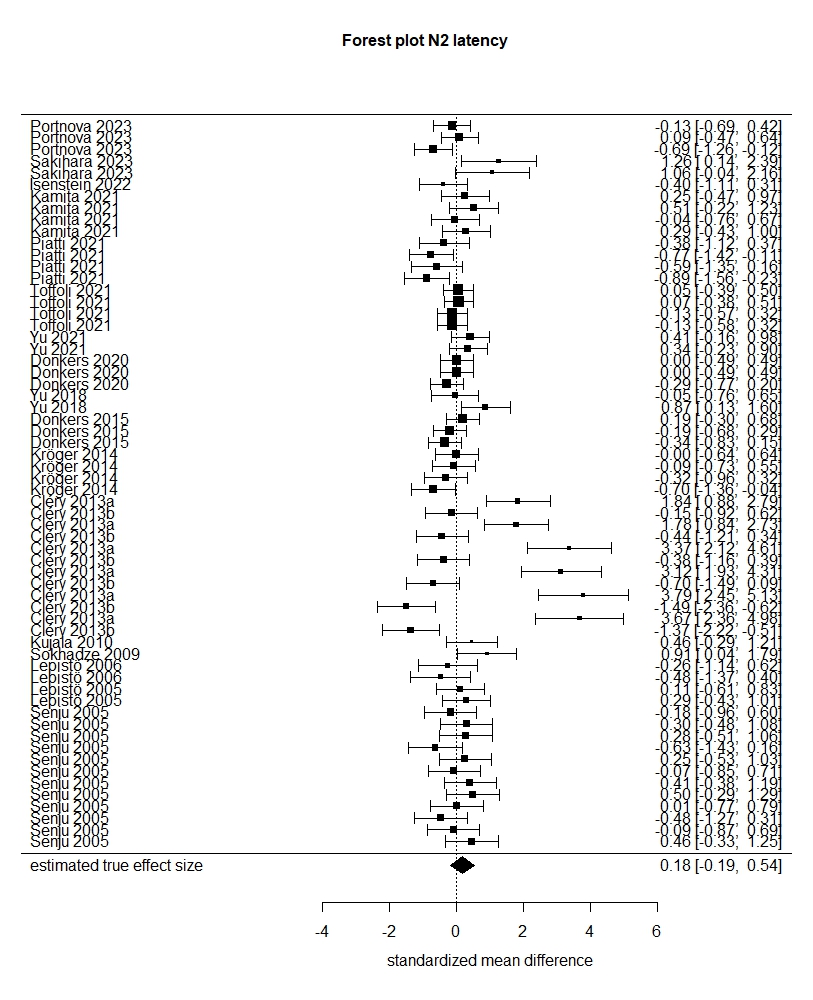

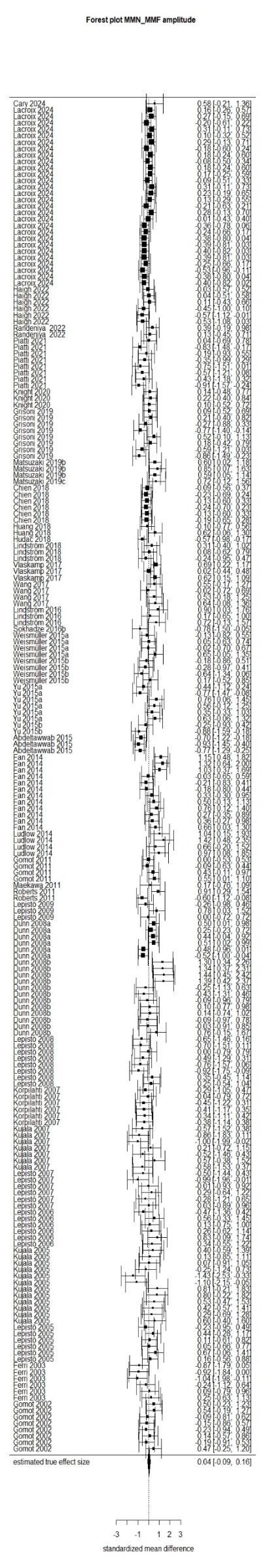

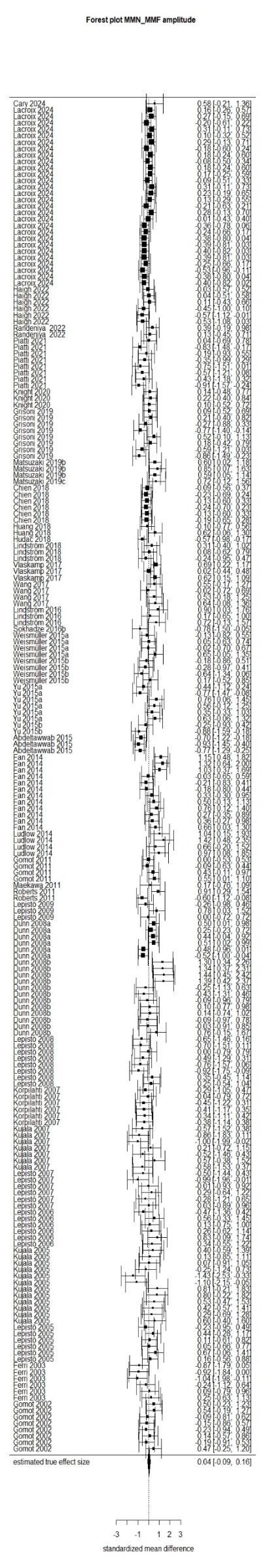

##### **eFigure 9. Forest Plot from amplitude differences in MMN/MMF component between groups.**

##### **eFigure 10. Forest Plot from latency differences in MMN/MMF component between groups.**

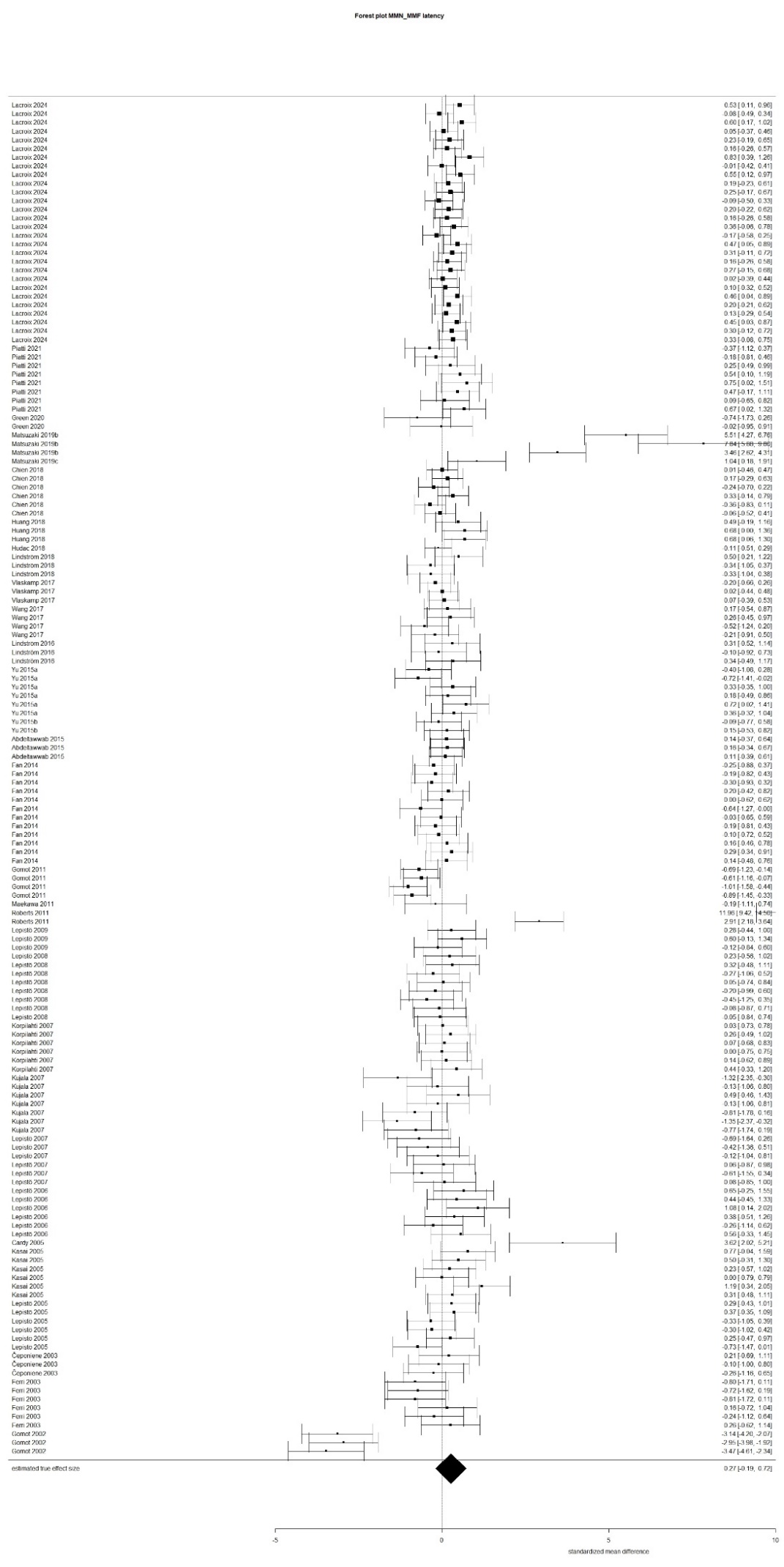

#### **Moderator analyses**

##### **eTable 16. Summary of significant and non-significant meta-analytic findings for amplitude moderator analyses (mixed-effects models fitted).**

| **Amplitude** | **k** | **Q_B_(df.p)** | **Q_W_(df.p)** | **SMD (p)** |
| --- | --- | --- | --- | --- |
| **P/M50** | | | | |
| EEG/MEG | NA | NA | NA | NA |
| Modality | NS | NS | NS | NA |
| Age | NS | NS | NS | NA |
| Language impairment | 14 | **29.09 (2, <.0001)** | 144.08 (37, <.0001) | NA |
| *Yes* | 2 | NA | NA | **2.45 (<.0001)** |
| *No* | 12 | NA | NA | -0.29 (0.20) |
| Task | NA | NA | NA | NA |
| Year of publication | NS | NS | NS | NA |
| Co-occurring condition | NA | NA | NA | NA |
| Medication | **16** | **33.60 (2, <.0001)** | 101.05 (14, <.0001) | NA |
| *Yes* | 4 | NA | NA | -1.47 (0.07) |
| ***No*** | 4 | NA | NA | **1.85 (0.03)** |
| Classification system | NS | NS | NS | NA |
| IQ | NS | NS | NS | NA |
| Sex | NS | NS | NS | NA |
| Quality rating | NS | NS | NS | NA |
| **P/M100** | | | | |
| EEG/MEG | NS | NS | NS | NA |
| Modality | NS | NS | NS | NA |
| Age | NS | NS | NS | NA |
| Language impairment | NS | NS | NS | NA |
| Task | NS | NS | NS | NA |
| Year of publication | NS | NS | NS | NA |
| Co-occurring condition | NS | NS | NS | NA |
| Medication | NS | NS | NS | NA |
| Classification system | NS | NS | NS | NA |
| IQ | NS | NS | NS | NA |
| Sex | NS | NS | NS | NA |
| Quality rating | NS | NS | NS | NA |
| **N100** | | | | |
| EEG/MEG | NA | NA | NA | NA |
| Modality | 93 | **8.54 (3, 0.04)** | 144.42 (90, 0.0002) | NA |
| *Visual* | 35 | NA | NA | -**0.27 (0.005)** |
| *Auditory* | 41 | NA | NA | 0.05 (0.57) |
| *Combined* | 17 | NA | NA | 0.06 (0.74) |
| Age | 93 | **9.08 (3, 0.03)** | 142.19 (90, 0.0004) | NA |
| *Children* | 34 | NA | NA | 0.09 (0.33) |
| *Adolescences* | 24 | NA | NA | **-0.27 (0.004)** |
| *Adults* | 35 | NA | NA | -0.02 (0.84) |
| Language impairment | NS | NS | NS | NA |
| Task | NS | NS | NS | NA |
| Year of publication | NS | NS | NS | NA |
| Co-occurring condition | NS | NS | NS | NA |
| **Amplitude** | **k** | **Q_B_(df.p)** | **Q_W_(df.p)** | **SMD (p)** |
| **N100** | | | | |
| Medication | NS | NS | NS | NA |
| Classification system | NS | NS | NS | NA |
| IQ | NS | NS | NS | NA |
| Sex | NS | NS | NS | NA |
| Quality rating | NS | NS | NS | NA |
| **N170** | | | | |
| EEG/MEG | NA | NA | NA | NA |
| Modality | NS | NS | NS | NA |
| Age | NS | NS | NS | NA |
| Language impairment | NS | NS | NS | NA |
| Task | 150 | 11.94 (5, 0.04) | 316.68 (145, <0.0001) | NA |
| *Target detection* | 99 | NA | NA | -0.13 (0.24) |
| *Oddball paradigm* | 6 | NA | NA | -0.13 (0.73) |
| *Passive task* | 22 | NA | NA | 0.19 (0.21) |
| *Discrimination task* | 9 | NA | NA | **0.68 (0.01)** |
| *Face recognition task* | 14 | NA | NA | 0.32 (0.18) |
| Year of publication | NS | NS | NS | NA |
| Co-occurring condition | NS | NS | NS | NA |
| Medication | NS | NS | NS | NA |
| Classification system | NS | NS | NS | NA |
| IQ | NS | NS | NS | NA |
| Sex | NS | NS | NS | NA |
| Condition type | NS | NS | NS | NA |
| Quality rating | NS | NS | NS | NA |
| **P/M200** | | | | |
| EEG/MEG | NS | NS | NS | NA |
| Modality | NS | NS | NS | NA |
| Age | NS | NS | NS | NA |
| Language impairment | NS | NS | NS | NA |
| Task | NS | NS | NS | NA |
| Year of publication | NS | NS | NS | NA |
| Co-occurring condition | NS | NS | NS | NA |
| Medication | 19 | 30.24 (2, <0.001) | 11.87 (17, 0.80)) | NA |
| *Yes* | 7 | NA | NA | 0.02 (0.90) |
| *No* | 12 | NA | NA | **-0.49 (<.0001)** |
| Classification system | NS | NS | NS | NA |
| IQ | NS | NS | NS | NA |
| Sex | NS | NS | NS | NA |
| Quality rating | NS | NS | NS | NA |
| **N200** | | | | |
| EEG/MEG | NA | NA | NA | NA |
| Modality | NS | NS | NS | NA |
| Age | 67 | 8.41 (3, 0.03) | 215.82 (64, <0.0001) | NA |
| *Children* | 40 | NA | NA | 0.05 (0.71) |
| *Adolescences* | 19 | NA | NA | **0.47 (0.02)** |
| *Adults* | 8 | NA | NA | 0.59. (0.08) |
| Language impairment | NS | NS | NS | NA |
| **Amplitude** | **k** | **Q_B_(df.p)** | **Q_W_(df.p)** | **SMD (p)** |
| **N200** | | | | |
| Task | NS | NS | NS | NA |
| Year of publication | 67 | 16.25 (6. 0.01) | 198.5466 (61. <.0001) | NA |
| *2000-2012.DSM-IV.TR* | 21 | NA | NA | **0.47 (0.02)** |
| *2013-2023.DSM-V* | 46 | NA | NA | 0.10 (0.39) |
| Co-occurring condition | NS | NS | NS | NA |
| Medication | NA | NA | NA | NA |
| Classification system | 67 | 16.25 (6, 0.01) | 198.55 (61, <0.0001) | NA |
| *only DSM IV+DSM IV-TR (without ICD)* | 34 | NA | NA | **0.47 (0.002)** |
| *only DSM V (without ICD)* | 1 | NA | NA | -0.45 (0.40) |
| *only ICD 10 (without DSM)* | 8 | NA | NA | -0.34 (0.19) |
| *only ADOS/+ADIR (since 2013)* | 16 | NA | NA | -0.00 (0.97) |
| *DSM IV+DSM IV-TR+ICD 10* | 4 | NA | NA | 0.57 (0.09) |
| *More than one DSM criteria/not mentioned which DSM criteria* | 4 | NA | NA | 0.38 (0.22) |
| IQ | NS | NS | NS | NA |
| Sex | 67 | 6.59 (1, 0.01) | 211.10 (65, <0.0001) | **-1.30 (0.03)** |
| Quality rating | NS | NS | NS | NA |
| **MMN/MMF** | | | | |
| EEG/MEG | NS | NS | NS | NA |
| modality | NS | NS | NS | NA |
| age | NS | NS | NS | NA |
| language impairment | NS | NS | NS | NA |
| task | NS | NS | NS | NA |
| year of publication | NS | NS | NS | NA |
| co-occurring condition | NS | NS | NS | NA |
| medication | NS | NS | NS | NA |
| Classification system | NS | NS | NS | NA |
| IQ | NS | NS | NS | NA |
| Sex | NS | NS | NS | NA |
| Quality rating | 210 | 11.35 (1, 0.0008) | 433.23 (209, <0.0001) | 0.08 (0.0008) |

*Note.* k, number of included effect sizes; NA, not available; NS, not significant.

##### **eTable 17. Summary of significant and non-significant meta-analytic findings for latency moderator analyses (mixed-effects models fitted).**

| **Latency** | **k** | **QB(df.p)** | **QW(df.p)** | **SMD (p)** |
| --- | --- | --- | --- | --- |
| **P/M50** | | | | |
| EEG/MEG | NS | NS | NS | NA |
| modality | NA | NA | NA | NA |
| age | NS | NS | NS | NA |
| language impairment | 47 | 7.69 (2, 0.02) | 181.66 (45, <0.0001) | NA |
| *Yes* | 12 | NA | NA | **0.72 (0.006)** |
| *No* | 35 | NA | NA | 0.38 (0.07) |
| task | NA | NA | NA | NA |
| year of publication | NS | NS | NS | NA |
| co-occurring condition | NA | NA | NA | NA |
| medication | NS | NS | NS | NA |
| Classification system | NS | NS | NS | NA |
| IQ | NS | NS | NS | NA |
| Sex | NS | NS | NS | NA |
| Quality rating | NS | NS | NS | NA |
| **P/M100** | | | | |
| **EEG/MEG** | 195 | 10.40 (2, 0.01) | 513.85 (185, <0.0001) | NA |
| *EEG* | 151 | NA | NA | 0.07 (0.45) |
| *MEG* | 36 | NA | NA | **0.50 (0.001)** |
| **Modality** | 151 | 7.00 (2, 0.03) | 498.79 (185, <0.0001) | NA |
| *Visual* | 115 | NA | NA | 0.06 (0.63) |
| *Auditory* | 72 | NA | NA | **0.32 (0.007)** |
| age | NS | NS | NS | NA |
| language impairment | NS | NS | NS | NA |
| task | NS | NS | NS | NA |
| year of publication | NS | NS | NS | NA |
| co-occurring condition | 111 | 13.36 (3, 0.004) | 498.62 (184, <0.0001) | NA |
| *Yes* | 12 | NA | NA | 0.02 (0.95) |
| *No* | 99 | NA | NA | **0.40 (0.001)** |
| medication | NS | NS | NS | NA |
| Classification system | NS | NS | NS | NA |
| IQ | NS | NS | NS | NA |
| Sex | NS | NS | NS | NA |
| Quality rating | NS | NS | NS | NA |
| **N100** | | | | |
| EEG/MEG | NS | NS | NS | NA |
| modality | NS | NS | NS | NA |
| age | NS | NS | NS | NA |
| language impairment | NS | NS | NS | NA |
| task | NS | NS | NS | NA |
| year of publication | NS | NS | NS | NA |
| co-occurring condition | NS | NS | NS | NA |
| medication | NS | NS | NS | NA |
| Classification system | NS | NS | NS | NA |
| IQ | NS | NS | NS | NA |
| Sex | NS | NS | NS | NA |
| Quality rating | NS | NS | NS | NA |
| **N170** | | | | |
| EEG/MEG | NA | NA | NA | NA |
| Modality | NA | NA | NA | NA |
| **N170** | | | | |
| Age | 119 | 12.28 (3, 0.007) | 370.61 (116, <0.0001) | NA |
| *Children* | 62 | NA | NA | 0.08 (0.63) |
| *Adolescences* | 18 | NA | NA | **0.63 (0.01)** |
| *Adults* | 39 | NA | NA | **0.47 (0.01)** |
| Language impairment | 119 | 7.60 (2, 0.02) | 388.10 (117, <0.0001) | NA |
| *Yes* | 9 | NA | NA | 0.18 (0.68) |
| *No* | 110 | NA | NA | **0.34 (0.006)** |
| Task | 115 | 15.51 (5, <0.008) | 213.27 (110, <0.0001) | NA |
| *Target detection* | 63 | NA | NA | **0.23 (0.04)** |
| *Oddball paradigm* | 6 | NA | NA | 0.07 (0.84) |
| *Passive task* | 22 | NA | NA | **0.32 (0.02)** |
| *Discrimination task* | 10 | NA | NA | -0.34 (0.18) |
| *Face recognition task* | 4 | NA | NA | 0.42 (0.06) |
| Year of publication | 119 | 12.47 (2; 0.0020) | 358.4879 (117; <.0001) | NA |
| *2000-2012.DSM-IV.TR* | 57 | NA | NA | **0.57 (0.001)** |
| *2013-2023.DSM-V* | 62 | NA | NA | 0.13 (0.41) |
| Co-occurring condition | NA | NA | NA | NA |
| Medication | NS | NS | NS | NA |
| Classification system | NS | NS | NS | NA |
| Condition type | 118 | 10.52 (2, 0.01) | 384.2285 (116, <0.0001) | NA |
| *Objects* | 26 | NA | NA | 0.21 (0.15) |
| *Faces* | 92 | NA | NA | **0.36 (0.004)** |
| IQ | NS | NS | NS | NA |
| Sex | NS | NS | NS | NA |
| Quality rating | NS | NS | NS | NA |
| **P/M200** | | | | |
| EEG/MEG | NA | NA | NA | NA |
| Modality | 39 | 15.94 (2, 0.0003) | 46.57 (37, 0.13) | NA |
| *Visual* | 21 | NA | NA | **0.45 (<.0001)** |
| *Auditory* | 18 | NA | NA | 0.09 (0.45) |
| Age | 41 | 14.14 (3, 0.003) | 50.66 (38, 0.08) |  |
| *Children* | 19 | NA | NA | 0.19 (0.16) |
| *Adolescences* | 11 | NA | NA | 0.25 (0.11) |
| *Adults* | 11 | NA | NA | **0.55** (**0.002**) |
| Language impairment | 41 | 18.11 (2, 0.0001) | 48.97 (39, 0.13) |  |
| *Yes* | 12 | NA | NA | -0.02 (0.92) |
| *No* | 29 | NA | NA | **0.37 (<.0001)** |
| Task | 41 | 31.72 (5, <0.0001) | 40.66 (36, 0.27) |  |
| ***Target detection*** | 10 | NA | NA | **0.53 (0.001)** |
| *Oddball paradigm* | 12 | NA | NA | -0.09 (0.5384) |
| *Passive task* | 11 | NA | NA | **0.46** (**0.0003)** |
| *Discrimination task* | 4 | NA | NA | 0.29 (0.06) |
| *Face recognition task* | 4 | NA | NA | 0.30 (0.14) |
| Year of publication | 41 | 9.83 (3, 0.02) | 55.14 (38, 0.04) | NA |
| *1994-1999.DSM-IV* | 8 | NA | NA | 0.13 (0.70) |
| *2000-2012.DSM-IV.TR* | 17 | NA | NA | **0.38** (**0.01)** |
| *2013-2023.DSM-V* | 16 | NA | NA | 0.27 (0.06) |
| **P/M200** | | | | |
| Co-occurring condition | NA | NA | NA |  |
| Medication | NA | NA | NA |  |
| Classification system | 41 | 14.43 (4, 0.01) | 50.74 (37, 0.07) |  |
| *only DSM III+DSM III-TR (without ICD)* | 11 | NA | NA | 0.03 (0.89) |
| ***only DSM IV+DSM IV-TR (without ICD)*** | 24 | NA | NA | **0.44 (0.0003)** |
| *only ADOS/+ADIR (since 2013)* | 5 | NA | NA | 0.21 (0.29) |
| *More than one DSM criteria/not mentioned* | 1 | NA | NA | -0.14 (0.75) |
| IQ | NS | NS | NS | NA |
| Sex | NS | NS | NS | NA |
| Quality rating | NS | NS | NS | NA |
| **N200** | | | | |
| EEG/MEG | NA | NA | NA | NA |
| Modality | NS | NS | NS | NA |
| Age | NS | NS | NS | NA |
| Language impairment | NS | NS | NS | NA |
| Task | NS | NS | NS | NA |
| Year of publication | NS | NS | NS | NA |
| Co-occurring condition | NS | NS | NS | NA |
| Medication | NS | NS | NS | NA |
| Classification system | NS | NS | NS | NA |
| IQ | NS | NS | NS | NA |
| Sex | NS | NS | NS | NA |
| Quality rating | NS | NS | NS | NA |
| **MMN/MMF** | | | | |
| EEG/MEG | 156 | 37.37 (2, <0.0001) | 545.70 (154, <0.0001) | NA |
| *EEG* | 143 | NA | NA | -0.13 (0.43) |
| *MEG* | 13 | NA | NA | **2.54 (<.0001)** |
| Modality | NS | NS | NS | NA |
| Age | NS | NS | NS | NA |
| Language impairment | 150 | 8.20 (2, 0.02) | 673.53 (148, <0.0001) | NA |
| *Yes* | 36 | NA | NA | **0.67 (0.02)** |
| *No* | 114 | NA | NA | 0.13 (0.61) |
| Task | NA | NA | NA | NA |
| Year of publication | NS | NS | NS | NA |
| Co-occurring condition | NS | NS | NS | NA |
| Medication | NS | NS | NS | NA |
| Classification system | NS | NS | NS | NA |
| IQ | NS | NS | NS | NA |
| Sex | NS | NS | NS | NA |
| Quality rating | NS | NS | NS | NA |

*Note.* k, number of included effect sizes; NA not available; NS not significant.

#### **Sensitivity analyses**

##### **eTable 18.Sensitivity analyses in amplitudes for each component**

|  | **k** | **Effect size** | **95% CI** | **QW (df.p)** | **P-Value** |
| --- | --- | --- | --- | --- | --- |
| **P/M50** | | | | | |
| Outlier analysis | 38 | -0.12 | -0.37 0.14 | 97.92(37.<0.00) | 0.37 |
| **P/M100** | | | | | |
| Outlier analysis | 227 | -0.09 | -0.20 0.03 | 428.48 (226. < 0.00) | 0.14 |
| **P/M200** | | | | | |
| No Outliers | | | | | |
| **N100** | | | | | |
| No Outliers | | | | | |
| **N170** | | | | | |
| Outlier analysis | 151 | 0.05 | -0.12 0.23 | 378.39 (150.< 0.00) | 0.55 |
| **N200** | | | | | |
| Outlier analysis | 65 | 0.24 | 0.04. 0.44 | 164.8269 (64. 0.00) | 0.02 |
| **MMN/MMF** | | | | | |
| No Outliers | | | | | |

*Note:* k, number of included effect sizes

##### **eTable 19.Sensitivity analyses in latencies for each component**

|  | **k** | **Effect size** | **95% CI** | **QW (df.p)** | **P-Value** |
| --- | --- | --- | --- | --- | --- |
| **P/M50** | | | | | |
| Outlier analysis | 45 | 0.25 | 0.04 0.45 | 83.05 (44. 0.00) | 0.02 |
| **P/M100** | | | | | |
| Outlier analysis | 192 | 0.10 | -0.01 0.20 | 348.87 (191. 0.00) | 0.06 |
| **P/M200** | | | | | |
| No Outliers | | | | | |
| **N100** | | | | | |
| No Outliers | | | | | |
| **N170** | | | | | |
| Outlier analysis | 115 | 0.27 | 0.09 0.45 | 227.08 (114. 0.00) | 0.00 |
| **N200** | | | | | |
| Outlier analysis | 58 | 0.10 | -0.17 0.37 | 123.64 (57. < 0.00 | 0.48 |
| **MMN/MMF** | | | | | |
| No Outliers | | | | | |

*Note*: k, number of included effect sizes

#### **Funnel Plots**

##### **eFigure 11. Funnel Plot displaying meta-analytical results obtained from fitting multilevel models to P/M50 component amplitudes and latencies.**

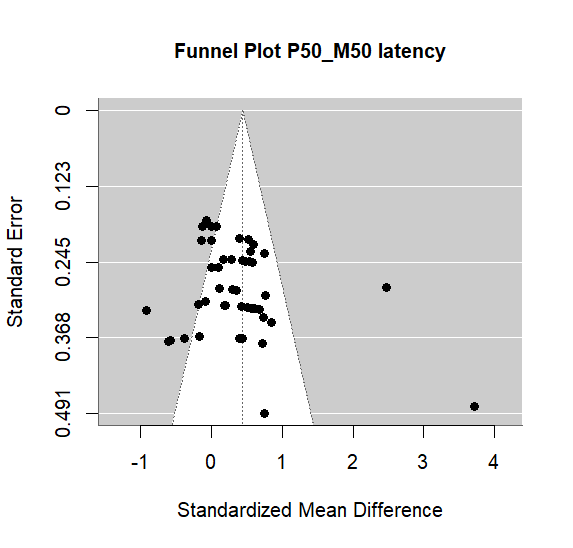

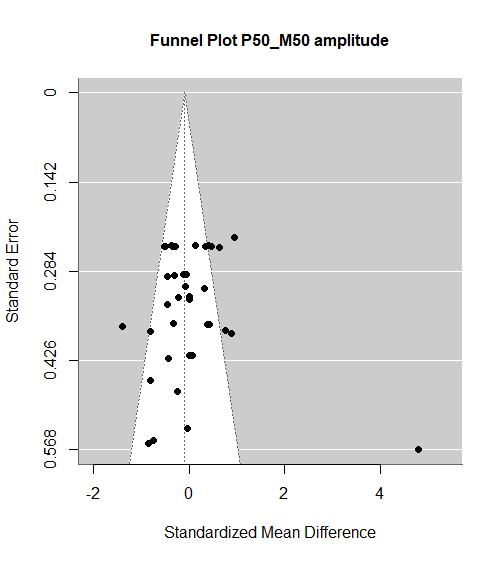

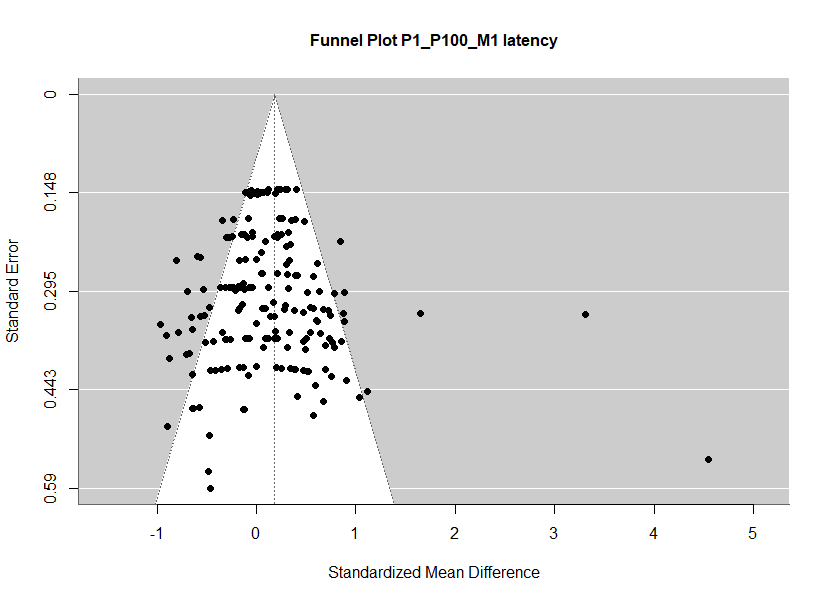

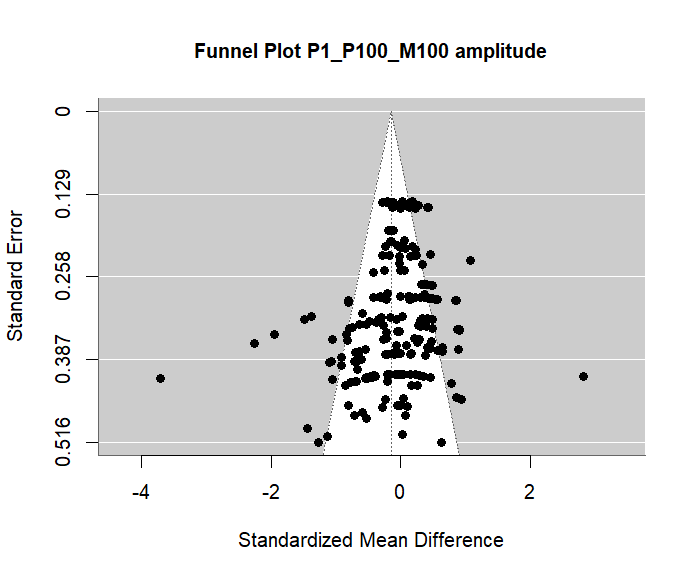

##### **eFigure 12. Funnel Plot displaying meta-analytical results obtained from fitting multilevel models to P/M100 component amplitudes and latencies.**

##### **eFigure 13. Funnel Plot displaying meta-analytical results obtained from fitting multilevel models to P/M200 component amplitudes and latencies.**

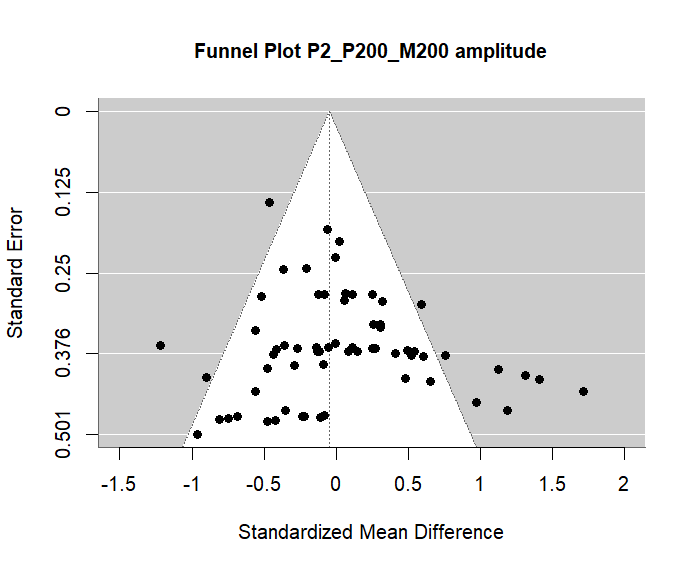

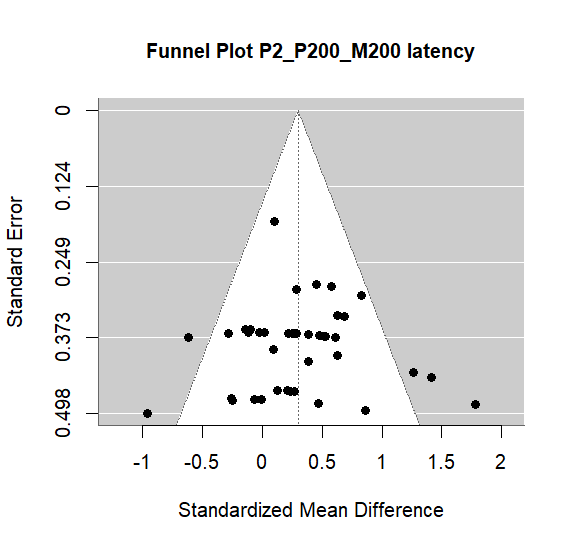

##### **eFigure 14. Funnel Plot displaying meta-analytical results obtained from fitting multilevel models to N100 component amplitudes and latencies**

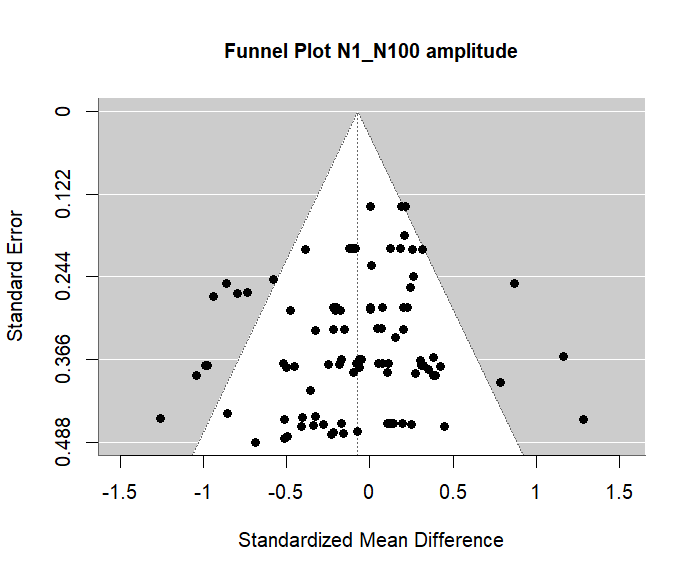

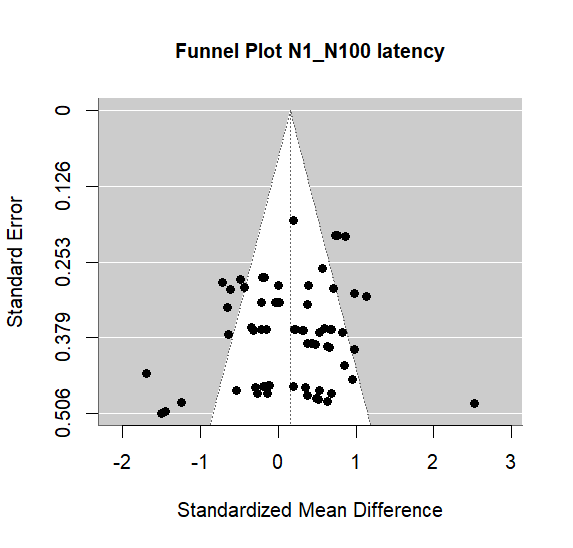

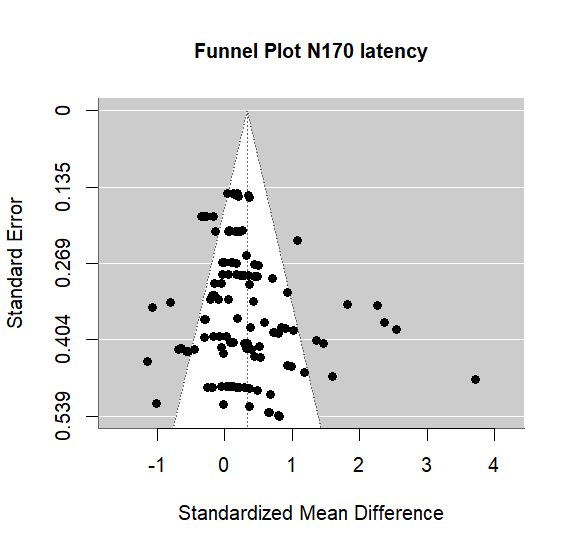

##### **eFigure 15. Funnel Plot displaying meta-analytical results obtained from fitting multilevel models to N170 component amplitudes and latencies.**

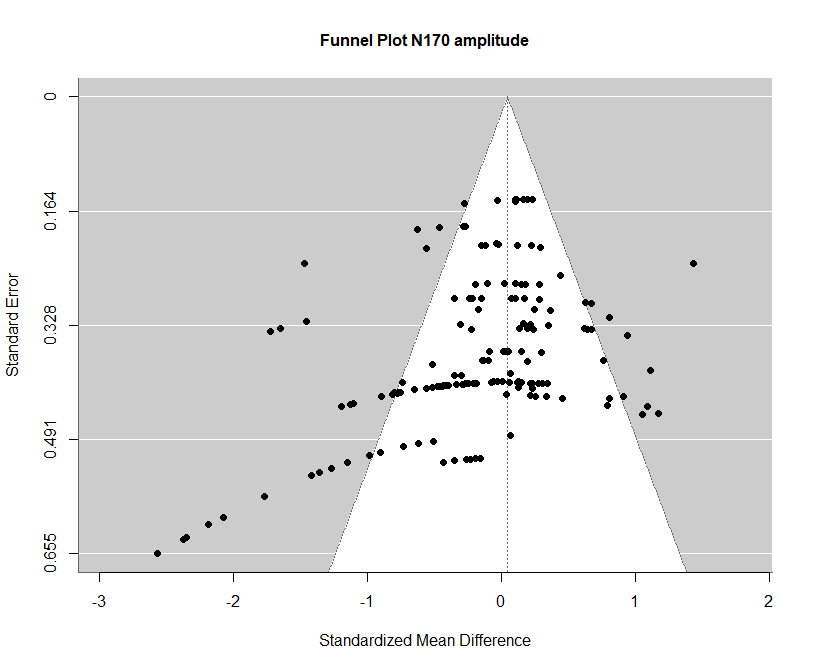

##### **eFigure 16. Funnel Plot displaying meta-analytical results obtained from fitting multilevel models to N200 component amplitudes and latencies.**

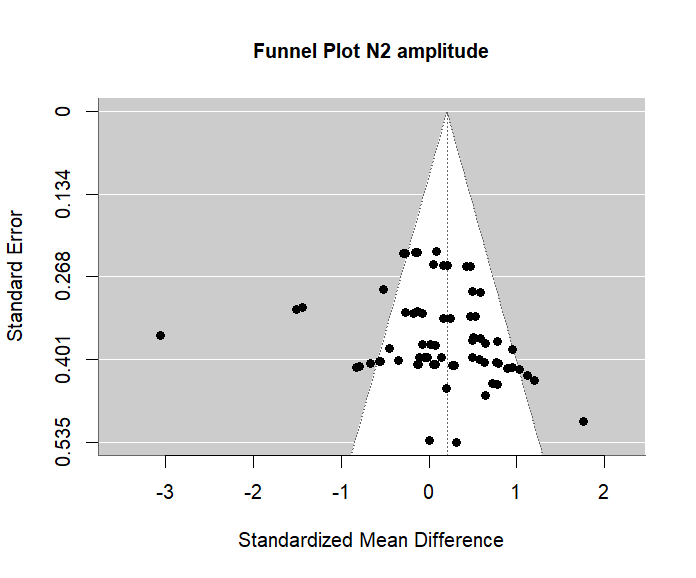

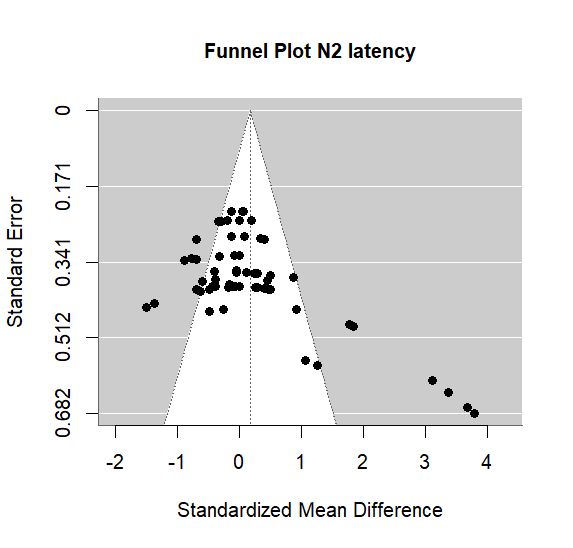

##### **eFigure 17. Funnel Plot displaying meta-analytical results obtained from fitting multilevel models to MMN/MMF component amplitudes and latencies.**

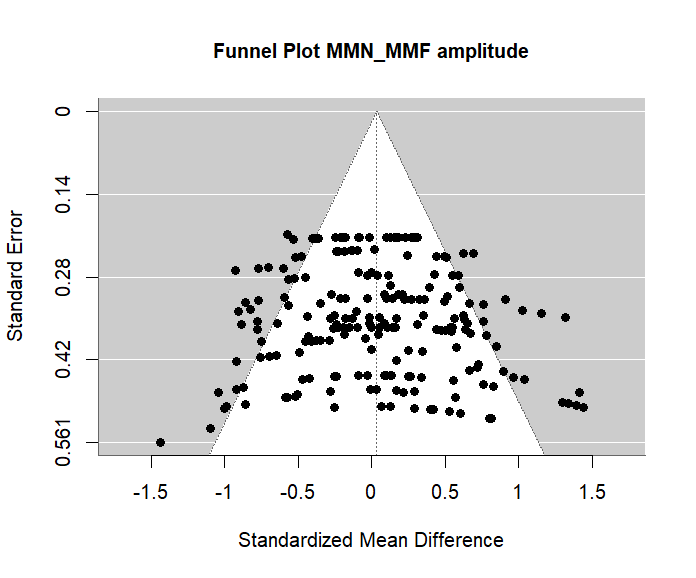

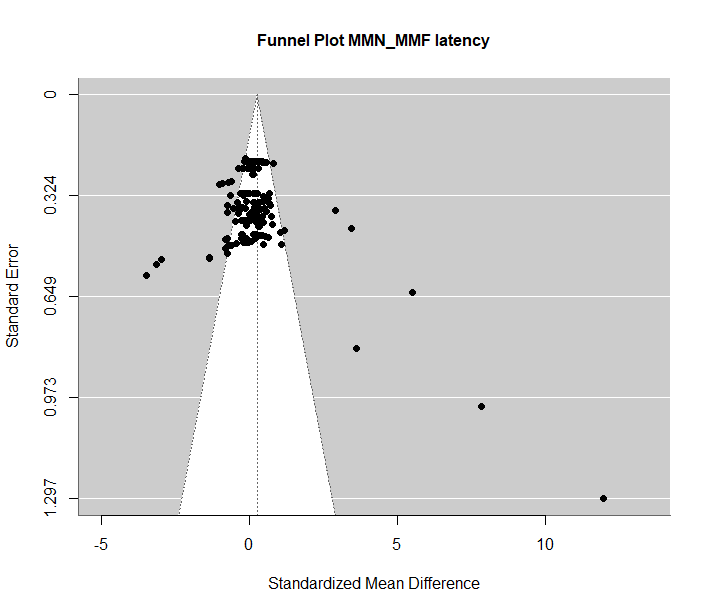

1. A study quality rating was implemented based on Newcastle-Ottawa quality assessment scale for case-control studies plus rating scale for EEG/MEG signal quality.
